# Noninvasive Detection of Fetal Genetic Variations through Polymorphic Sites Sequencing of Maternal Plasma DNA

**DOI:** 10.1101/2021.01.26.21250573

**Authors:** Song Gao

**Affiliations:** The State Key Laboratory Breeding Base of Basic Science of Stomatology & Key Laboratory of Oral Biomedicine Ministry of Education, School & Hospital of Stomatology, Wuhan University, Wuhan, China, 430079

**Keywords:** noninvasive prenatal testing, amplicon sequencing, goodness of fit, polymorphic site, fetal fraction, robust linear regression

## Abstract

Non-invasive prenatal testing (NIPT) for common fetal aneuploidies using circulating cell free DNA in maternal plasma has been widely adopted in clinical practice for its sensitivity and accuracy. However, the detection of subchromosomal abnormalities or monogenetic variations showed no cost-effectiveness or satisfactory accuracy. Here we developed an assay, the goodness-of-fit and graphical analysis of polymorphic sites based non-invasive prenatal testing (GGAP-NIPT), to simultaneously detect fetal chromosomal/subchromosomal and nucleotide level abnormalities. In each sample, fetal fraction was estimated using allelic counts of reference polymorphic sites and a robust linear regression model. Then the genotype of each polymorphic site was estimated using allelic goodness of fit test. Finally, monogenic mutations were detected using allelic wildtype and mutant counts of each target site, and chromosomal/subchromosomal abnormalities were identified by collective analysis of all target polymorphic sites. Such an analytic approach was highly accurate for detecting aneuploidies, microdeletions or microduplications and monogenic mutations for simulated samples with different fetal fractions and sequencing depths. Moreover, more than 93% of fetal monogenic mutations were correctly identified for target hotspot sites amplified using circulating or barcode-enabled single-molecule assays. With the aid of sample replicates, higher detection accuracy was observed. Through target polymorphic sites sequencing, all chromosomal/subchromosomal and monogenic abnormalities could be detected simultaneously, facilitating the extension of NIPT to an expanded panel of genetic disorders in a cost-effective manner.

## Introduction

Noninvasive prenatal testing (NIPT) is now widely used for the detection of fetal chromosomal aneuploidies and certain copy number variations, where cell-free DNA (cfDNA) in maternal plasma^1^ was analyzed by whole-genome sequencing (WGS) ^2^, target analysis of nonpolymorphic regions^3^, single-nucleotide polymorphism (SNP) sequencing^4^ or microarray^5^. NIPT showed high test sensitivity and specificity for common fetal aneuploidies, such as trisomies 21, 18 and 13, but low in detecting subchromosomal deletions and duplications^6^, especially when the genomic aberrations were small^7^. For monogenic disorders, different noninvasive approaches have been developed^8^, but the application of such methods in clinical practice has lagged behind aneuploidy testing due to high costs and technical challenges. Due to different test philosophies, NIPT approaches currently in practice could not be extended to detect monogenetic disorders in a cost-effective manner.

In cfDNA, a certain number of polymorphic sites showed allelic imbalance due to the presence of fetal DNA and characteristic relative allelic ratios were observed when there were fetal aneuploidies. For example, when the fetus inherits a paternal allele different from the mother’s (Fig. S1), fetal aneuploidies can be detected using relative allelic counts. Here we proposed an assay, goodness-of-fit and graphical analysis of polymorphic sites based non-invasive prenatal testing (GGAP-NIPT), to detect fetal abnormalities at the chromosomal/subchromosomal and monogenic sequence levels simultaneously, which was shown sensitive and accurate for all test samples.

## Materials and Methods

### Dataset

The insertion/deletion polymorphism^9^ dataset (BioProject ID: PRJNA387652) and the replication^10^ dataset (BioProject ID: PRJNA517742) were retrieved from the NCBI Sequence Read Archive (SRA). The wilson validation dataset was retrieved from Supplementary Table 4^11^ and the wilson disease dataset from Table 2^11^. The hbb dataset was retrieved from Table S1^12^, the arnshl dataset from Supplementary Table S2^13^ and the cfbest dataset was retrieved from Table S8^14^. The simulated datasets were generated using ART^15^ simulator. Details of all datasets were described in Supplementary Methods.

### Reads Processing and Mapping

Reads retrieved from SRA or simulated were filtered out using custom scripts, and alleles for each site were identified using unique sequences. Whole genome sequencing reads were mapped by bowtie2^16^. Details of reads processing were described in Supplementary Methods.

### Fetal Fraction Estimation by Allelic Read Counts

For each polymorphic site, read counts for all alleles were sorted in descending order and labeled as R1, R2, R3, etc. Then the possible maternal-fetal genotype was estimated using allelic read counts (Fig. S2) followed by the estimation of fetal and total read counts (Table S1). Finally, fetal fraction was estimated using fetal and total read counts and a robust linear regression model (Supplementary Methods).

### Fetal Fraction Estimation by Whole Genome Sequencing

Fetal fraction was calculated as described using the formula^17^ 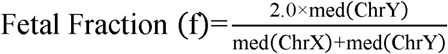, where med(ChrX) and med(ChrY) represent the median read counts of the 50-kb bins on the X and Y chromosomes, respectively. Briefly, the 50-kb bins from the X and Y chromosomes were extracted and bins having too low or too high read counts were filtered out. Fetal fractions for the 61 samples from PRJNA387652^18^ were calculated using the median count values of X bins and Y bins.

### Maternal-Fetal Genotype Estimation

For each polymorphic site, reads for each allele were counted, followed by the calculations of Akaike information criterion (AIC) scores for all possible genotype models using goodness-of-fit test^19^. Then, the genotype was estimated to be the model with the minimal AIC, and ΔAIC was calculated as the absolute difference between the AICs of different models (Supplementary Methods).

### Statistical Analysis

Statistical analysis was performed in R (version 3.5.1)^20^. AICs were calculated using custom scripts.

## Results

### Fetal Fraction Estimation

For each polymorphic site on the reference chromosome, one of five maternal-fetal genotypes is possible, and the genotype as well as read counts amplified from either the maternal or the fetal genomic materials can be estimated roughly by analyzing relative counts of different alleles (Fig. 1A-B; Fig. S2; Table S1). Obviously, fetal read count should be in proportional to total read count for each polymorphic site in a sample, and fetal fraction was estimated as the overall slope of a fitted robust linear regression line model without an intercept (Fig. 1C-D). A high degree of correlation was observed for fetal fractions estimated this way and that estimated using the WGS method^9^ when low quality WGS samples were excluded in the analysis (Fig. 1E; Fig. S3). As robust linear regression was not sensitive to outliers, nearly identical fetal fraction estimations were observed (Fig. 1F) for replicate samples prepared independently^10^, indicating that the method for fetal fraction estimation was accurate and reliable. For our simulated samples, great estimation accuracy was observed for samples with fetal fraction ≥5% and sequencing coverage ≥ 2000 (Fig. S4).

**Fig. 1.**
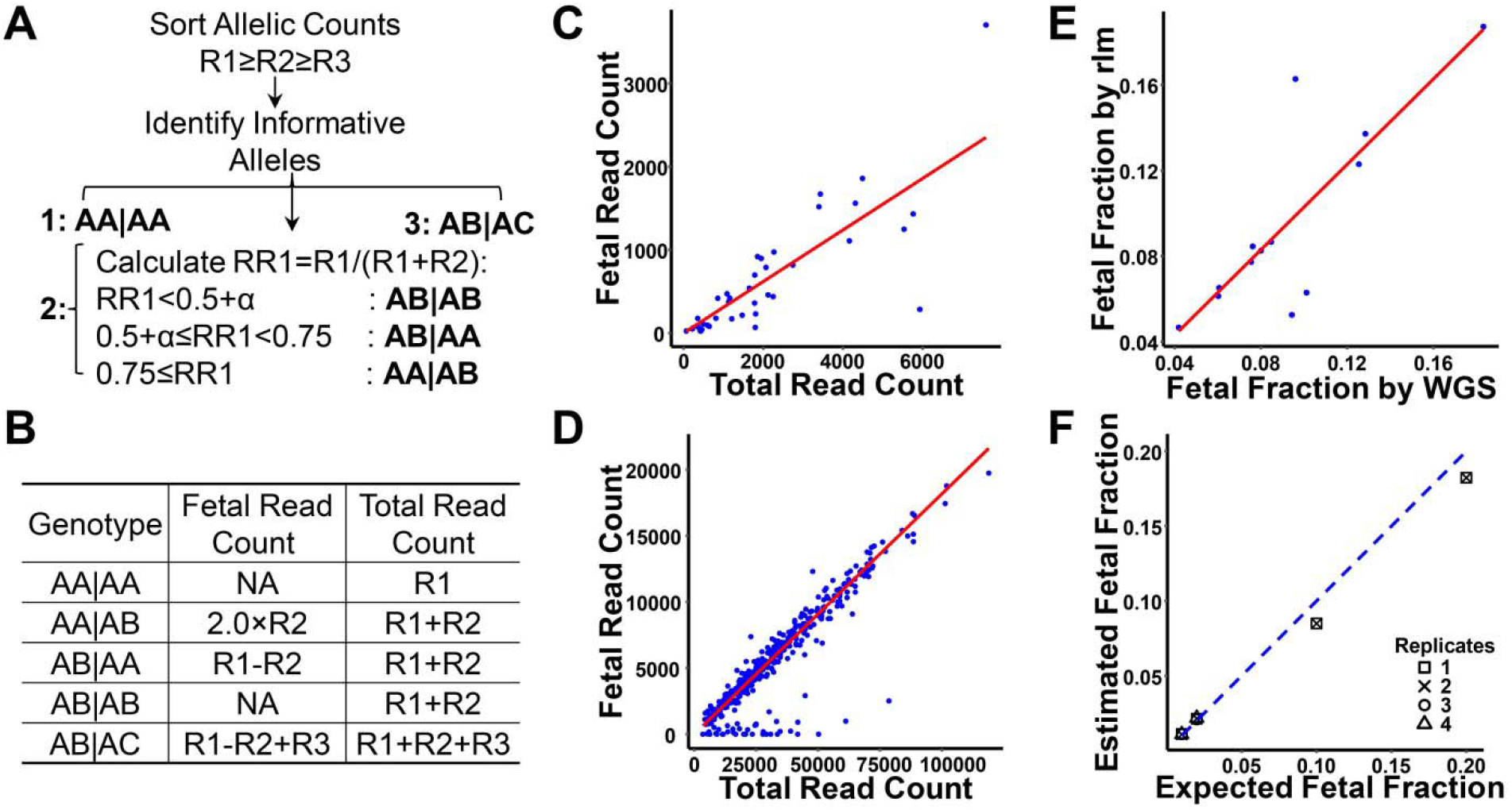
Fetal fraction estimation. (A) Genotype estimation for each polymorphic site using allelic counts. (B) Expected fetal read count and total read count for each genotype. (C, D) Representative plots from the insertion/deletion polymorphism dataset (C) and replication dataset (D). A robust linear regression line was fitted (red line, model y=βx+0), and fetal fraction was estimated to be the model coefficient (β). (E) Fetal fractions were estimated for each insertion/deletion polymorphism sample by both allelic read counts method (rlm) and WGS method (red line was the fitted regression line y∼x), excluding low quality WGS samples. (F) Expected and estimated fetal fractions for replication samples (blue line: y=x). α: background noise threshold.

### Discrete Nature of Relative Allelic Counts

As there are only a limited number of maternal-fetal genotypes (Table S2) and there are a great number of polymorphic sites in a sample, relative allelic counts of polymorphic sites were clustered in different groups and each group corresponded to a distinct genotype (Fig. 2A), whereas the genotype of each polymorphic site could be determined using either graphical analysis of relative allelic counts (Fig. 2B; Table S2; Fig. S5) or allelic goodness-of-fit test (Fig. 2C-E). Here, Akaike information criterion (AIC) was used for comparing non-nested genotype models for each site (Fig. 2E) and ΔAIC for estimating the genotype fitness between different models. In general, the genotype of a target was estimated to be the one having the minimal AIC (Fig. 2E) and ΔAIC was calculated as the absolute difference between the AICs of the two best fitted models. However, such ΔAIC values were highly affected by both fetal fractions and total allelic counts (Fig. S6; Fig. 2F), and nearly similar magnitude of values were observed when ΔAIC values were adjusted by fetal fraction and total read counts (Fig. S6; Fig. 2G), indicating that the adjusted ΔAIC could be a good measure for comparing fitnesses of different genotypes. As expected, allelic goodness-of-fit test was highly accurate in estimating maternal-fetal genotypes of each polymorphic site for simulated samples with different fetal fractions and different sequencing depths (Fig. S7).

**Fig. 2.**
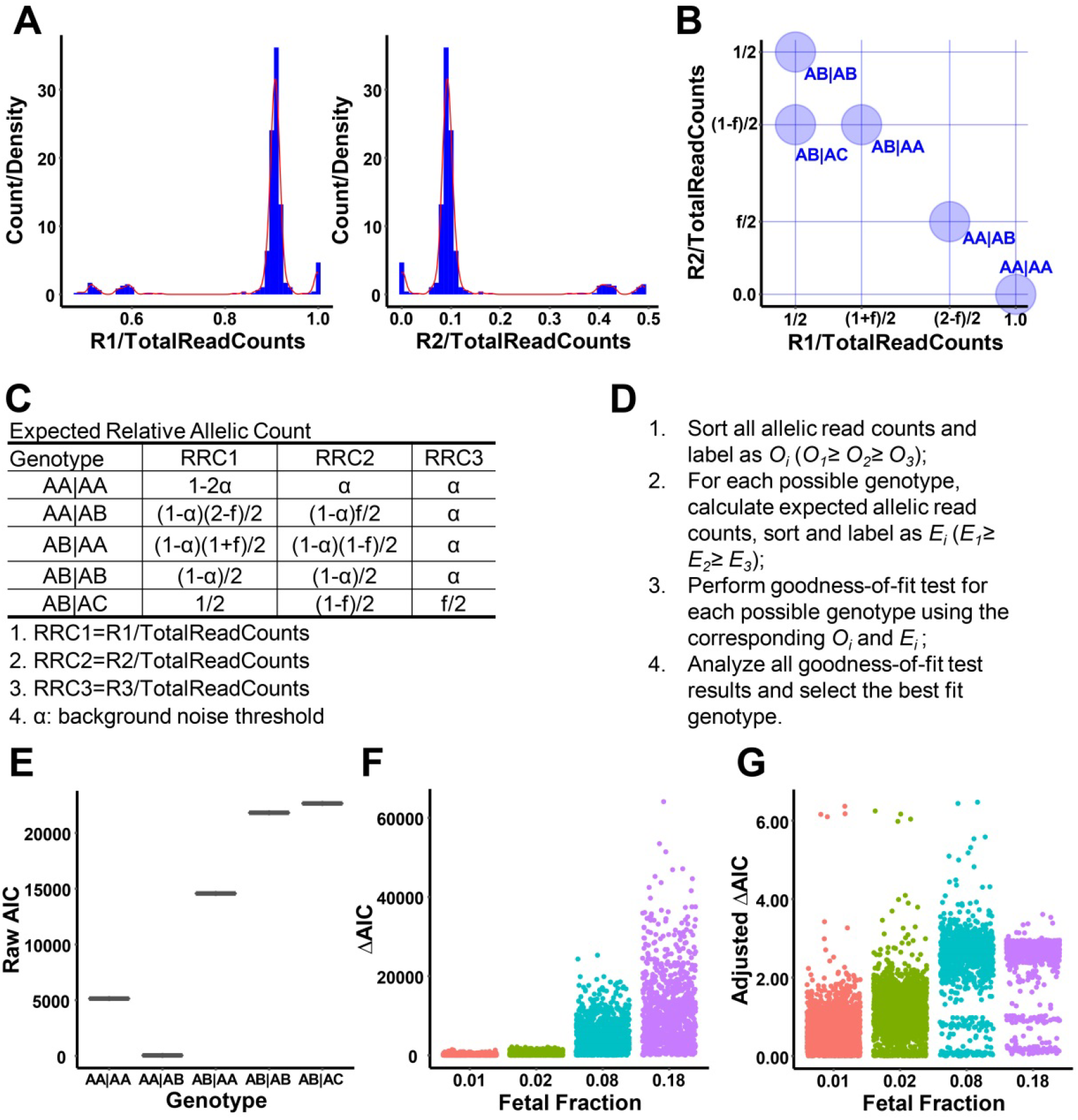
Maternal-fetal genotype estimation. (A) Discrete clusters of relative allelic counts for a representative sample from the replication dataset (blue: histogram count, red: density). (B) Expected relative allelic clusters for polymorphic sites on normal reference chromosomes. (C) Expected relative allelic counts for each normal maternal-fetal genotype. (D) Steps to estimate maternal-fetal genotype for each polymorphic site (allelic goodness-of-fit test). (E) A representative plot for genotype estimation using allelic goodness-of-fit test. As AA|AB model had the minimal raw AIC, the target genotype was estimated to be AA|AB. (F,G) ΔAIC (F) and adjusted ΔAIC (G) were calculated for each polymorphic site of the replicate samples grouped by estimated fetal fraction. ΔAIC= absolute AIC differences between two best fitted models. Adjusted ΔAIC= ΔAIC/TotalCount/FetalFraction.

### Detection of Chromosomal Aneuploidies

When there is a fetal aneuploidy, all polymorphic sites on the target chromosome are affected, and the relative allelic counts for each polymorphic site are changed due to the absence of one chromosome or the presence of one extra chromosome (Table S2-S4). To detect fetal aneuploidy, two models were checked, whereas one model (normal model) assumed that both the maternal and the fetal chromosomes were normal disomy and the other model (aneuploidy model) assumed that the maternal chromosome was disomy and the fetal chromosome was aneuploidy. Then each polymorphic site on the target chromosome was fitted with both a genotype from the normal model and a genotype from the aneuploidy model, and the overall fitness of all polymorphic sites for the normal model was compared with the overall fitness for the aneuploidy model (Fig. 3A). Such an approach might seem unsound mathematically, but were sensitive and reliable to detect chromosomal aneuploidies for our simulated samples (Fig. 3K,L), possibly due to its similarity to repeated tests of goodness-of-fit^19^ whereas each polymorphic site was considered as an experimental repetition. As nearly all polymorphic sites on the target chromosome showed some positive contributions to the correct model fittings (Fig. S8A,B), nearly all chromosomal aneuploidies were detected with high accuracy for our simulated samples (Fig. 3B,C,K,L). In addition, distinct genotype clusters were observed when the relative allelic counts of target polymorphic sites were plotted, and such characteristic cluster distributions were informative enough to identify fetal aneuploidies as well (Fig. 3G,H; Fig. S8C,D; Fig. S9,S10).

**Fig. 3.**
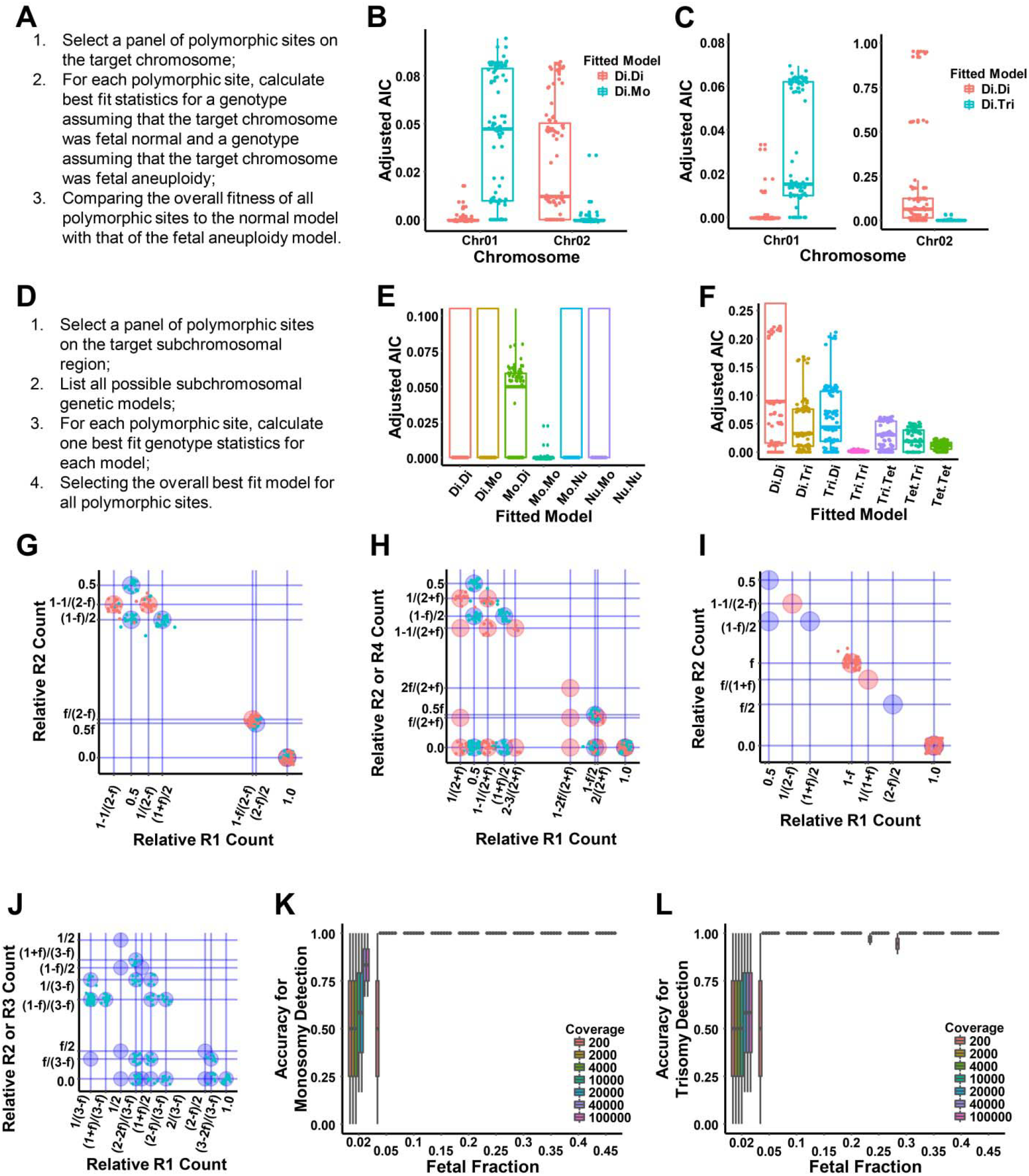
Chromosomal or subchromosomal abnormality detection. (A) Steps to detect fetal aneuploidy using overall goodness-of-fit test for all polymorphic sites. (B) Detecting fetal monosomy. Chr01: simulated Di.Di chromosome; Chr02: simulated Di.Mo. (C) Detecting fetal trisomy. Chr01: simulated Di.Di; Chr02: simulated Di.Tri chromosome. (D) Steps to detect fetal subchromosomal abnormality using overall goodness-of-fit test for all polymorphic sites. (E) Detecting fetal subchromosomal deletions. As Mo.Mo model was the best fit for all target polymorphic sites, both the mother and the fetus were heterozygous for the microdeletion. (F) Detecting fetal subchromosomal duplications. As Tri.Tri model was the best fit for all target polymorphic sites, both the mother and the fetus were heterozygous for the microduplication. (G-J) Detecting fetal abnormalities using relative allelic counts plots. (G) Detecting fetal monosomy. Blue: reference normal chromosome. Red: target chromosome. From the characteristic cluster positions, the target chromosome was estimated to be normal for the mother but monosomy for the fetus. (H) Detecting fetal trisomy. Blue: reference normal chromosome. Red: target chromosome. From the characteristic cluster positions, the target chromosome was estimated to be normal for the mother but trisomy for the fetus. (I) Detecting subchromosomal microdeletion. From characteristic clusters, both the mother and the fetus were heterozygous for the microdeletion. (J) Detecting subchromosomal microduplication. From characteristic clusters, the mother was heterozygous and the fetus was homozygous for the microduplication. (K) Detecting accuracy for simulated normal and monosomy samples. (J) Detecting accuracy for simulated normal and trisomy samples.

### Detection of Subchromosomal Abnormalities

In most cases, subchromosomal abnormalities could be tested in a way similar to detecting chromosomal aneuploidies, whereas a model of a healthy mother with a healthy fetus was compared with a model of a healthy mother with an affected fetus (Fig. 3A). For some microdeletions^21^ and microduplications^22^, the heterozygotes could be phenotypically normal and only the homozygotes showed clinical symptoms. In such cases, all possible models should be tested and the overall best fitted model for all polymorphic sites on the target region was selected (Fig. 3D). As expected, subchromosomal microdeletion or microduplication could be detected with accuracy using either collective analysis of allelic goodness of fit test for each polymorphic site in the target region (Fig. 3E,F; Fig. S11,S12) or relative allelic counts plots of all target polymorphic sites (Fig. S13,S14; Fig. 3I,J; Table S5,S6). For subchromosomal microduplications, too many distinct clusters were expected when all possible models were considered (Fig. S14A). In such a case, a simpler plot containing only models with a healthy fetus was generated, and when allelic clusters of target polymorphic sites were not in the expected positions (Fig. S14C), abnormal fetus was expected and further analysis should be followed.

### Detection of Short Genetic Variations

Single-base-pair substitutions, small (≤20bp) deletions, small (≤20bp) insertions and small (≤20bp) indels are the major types of mutations associated with human inherited diseases reported in the Human Gene Mutation Database (HGMD)^23^. To detect such genetic variations, the genotype of each target site was estimated using allelic goodness-of-fit test followed by sequence analysis of different alleles (Fig. 4A,C; Fig. S15A). Alternatively, fetal genetic mutation of each target site could be detected using a relative allelic plot whereas the relative count of the most abundant mutant allele was plotted against that of the wildtype allele (Fig. 4B; Fig. S15B-D; Table S7,S8).

**Fig. 4.**
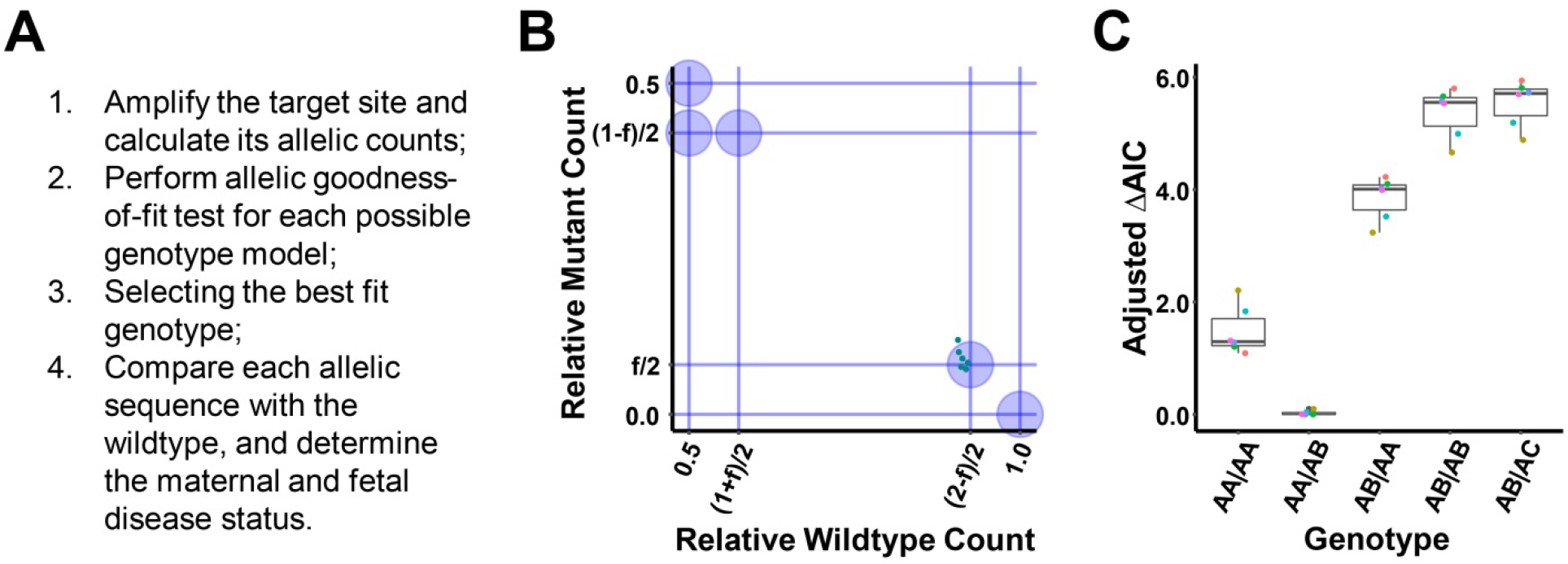
Detection of fetal short genetic variations. (A) Steps to detect sequence-level variation using goodness-of-fit test and wildtype sequence comparison. (B, C) Detecting target mutations by relative allelic counts plot (B) or allelic goodness-of-fit test (C). Maternal and paternal plasma samples were mixed, mimicking homozygous-heterozygous maternal-fetal genotypes with a fetal fraction of 15% (6 replicates). Each color represented a replicate sample. From the allelic plot (B), the mother was homozygous wildtype and the fetus was heterozygous mutant for the target site. From the goodness-of-fit test (C), the maternal-fetal genotype was estimated to be AA|AB. As wildtype counts were the major components, A was a wildtype allele and B was a mutant allele, hence the mother was wildtype and the fetus was heterozygous mutant.

### Testing Performance for Detecting Monogenic Mutations

To detect fetal monogenic mutations noninvasively, target hotspot sites were amplified from maternal plasma cfDNA using circulating single-molecule amplification and resequencing technology (cSMART)^11-13^ or barcode-enabled single-molecule test (cfBEST)^14^, whereas fetal fraction for each sample was estimated and allelic read counts of each target site were reported as mutant and wildtype counts. In the meanwhile, genotypes of the mothers, the fathers, the fetuses and/or the probands were determined using conventional molecular diagnostic techniques^11-14^. Here, we estimated the genotype of each target site using the sample’s fetal fraction and read counts of mutant and wildtype alleles which were the only information required by our allelic goodness of fit method, and then compared the estimated genotype with the true genotype reported. Using allelic counts of a single target site, 100% estimation accuracy was observed for all maternal genotypes, and 95.7%, 93.4% and 96.8% for fetal genotypes of the hbb dataset^12^, the arnshl dataset^13^ and the cfbest dataset^14^, respectively (Table 1; Table S9-S12). Moreover, when both the mother and the fetus were homozygous wildtype, 100% accuracy was observed (Table S9-S12). For all genotypes estimated incorrectly, mutant alleles well above the noise threshold were detected (Table S9-S12). Such an observation supports the notion that a small number of replicates (such as 1-2 replicates) are used for each target site to screen monogenic mutations, which should be accurate when both the mother and the fetus are wildtype, and if a mutant allele is detected, further analysis is followed. Such a two-tier screening strategy should be accurate and cost-effective, especially when the disease prevalence is low. In addition, when replicate samples were analyzed together, improved estimation accuracy was observed for the library or sequencing level repetition dataset (Fig. S16). Similarly, in the wilson validation dataset (Fig. S17-S20; Fig 4B,C)^11^, two sites out of eight from the 5% fetal fraction replicate mixtures (Table S13) were incorrectly estimated when each sample was analyzed individually, however, unambiguous genotype estimation was observed when all eight replicates were analyzed in a group as a single sample (Table S13; Fig. S17). Moreover, relative small adjusted ΔAIC values were observed for sites estimated incorrectly (Table S10-S12, Fig. S21), suggesting that the differences between two best fitted models were small for wrong estimated sites and test with repetitions would be more appropriate for such cases.

**Table 1.** Performance for detecting monogenic mutations

| Dataset | Amplification Method | Number of plasma samples | Number of sites | Correct Estimated Genotype |  | Accuracy for Genotype Estimation |  |
| --- | --- | --- | --- | --- | --- | --- | --- |
|  |  |  |  | Maternal | Fetal | Maternal | Fetal |
| hbb<sup>12</sup> | cSMART | 102 | 188 | 188 | 180 | 100% | 95.70% |
| arnshl<sup>13</sup> | cSMART | 80 | 137 | 137 | 128 | 100% | 93.43% |
| cfbest<sup>14</sup> | cfBEST | 143 | 189 | 189 | 183 | 100% | 96.83% |

## Discussion

Through amplicon sequencing of polymorphic and target sites of maternal plasma DNA, here we described an approach to detect chromosomal, subchromosomal and sequence-level abnormalities simultaneously, whereas a panel of polymorphic sites from normal reference chromosomes, a panel of polymorphic sites on the target chromosomal/subchromosomal regions, and a panel of specific target sites were amplified, sequenced and analyzed. As the same underlying principle was applied to amplify targets, it would be cost-effective for detecting abnormalities involving both large and small genomic fragments and easy to extend for detecting new abnormalities, which was unattainable by other reported NIPT methods so far.

To estimate fetal fraction, allelic read counts of each polymorphic sites were linear transformed, which not only preserved linear relationships between variables, but also improved the interpretability of the data whereas the correlation between variables was easy to understand. Moreover, allelic read counts for different underlying genotypes could be analyzed together after such linear transformations, and a robust linear regression could be fitted, which was insensitive to outliers. On the contrary, methods current available for estimating fetal fractions^17,24,25^ were based on either medians or statistical distributions, whereas only median values were used or models sensitive to outliers were applied. Hence the method for estimating fetal fraction using a robust linear regression model should be accurate inherently as all data points were included and the effects of outliers were minimized, and fetal fractions even at the level of 1% were consistently and accurately measured for repetitive samples (Fig. 1F).

For monogenic mutations, more than 90% of the target mutations were correctly identified for three public datasets (Table 1). Obviously, some of the mutation sites were not correctly detected by relative allelic ratios^11^ or maximal likelihood estimations^14^. When samples were tested using replicates, it is difficult to analyze a group of relative values or probabilities collectively to give a conclusive estimate using the current available methods. Here we identified a measure, the adjusted ΔAIC value, to detect target mutation using allelic counts of a single site, which was a good measure for the fitness of allelic counts to the estimated genotype. Moreover, the adjusted ΔAIC values of multiple replicate samples could be analyzed collectively, and the more the replicates used, the higher the estimation accuracies observed (Fig. S17). Therefore, when low adjusted ΔAIC value was observed for a site, which indicated non-optimal fitness of the estimated genotype as the two best fitted models had similar adjusted AIC values, analysis with repeated samples were desired, and all independent repeat data could be analyzed together. Such a strategy could increase the detection accuracy for monogenic mutations.

For aneuploidies, microdeletions and microduplications, a large number of polymorphic sites in the target regions were expected. Each target site was independent and chromosomal or subchromosomal abnormalities were detected using the trends of most sites by our method, which was less sensitive to outliers, while z-score^2^ or maximal likelihood estimation^4^ based methods were highly affected by outliers. In addition, when the adjusted ΔAIC values between the normal and abnormal models were too low, analysis of sample repeats were desired, and all repeated data could be analyzed together. For z-score^2^ or maximal likelihood estimation^4^ based methods, it was difficult to analyze the repeated data collectively and it was difficult to judge which results were reliable when inconsistent repetitive results were observed. As indicated in detecting monogenic mutations, when there were not enough polymorphic sites in the target region when detecting microdeletions or microduplications, each target polymorphic site could be amplified separately in different tubes as different sites, and then all sites were analyzed collectively. Such a strategy could be used to detect abnormal short genomic fragments with accuracy. In addition, when doing sample repeats, a different panel of target polymorphic sites could be used, especially when the results of the original set showed limited genotype diversity. As all maternal genotypes were accurately estimated for both real mutation samples and all our simulated samples even when there were aneuploidies or copy number variations for some targets, maternal genotype estimation for each polymorphic site was expected to be accurate.

Therefore, the exact same routines for SNP based NIPT could be used to detect chromosomal/subchromosomal abnormalities as well whereas the maternal genotype for each target could be estimated by our method. As no separate maternal genotypes determination was performed in this case, it should be more cost-effective than SNP based NIPT approaches. In our method, each target site was independent, and no cross-sample or cross-site comparisons were performed. Therefore, each target could be amplified individually or in a multiplex manner, especially when detecting multiple abnormalities simultaneously whereas it was challenging to amplify all targets in a single tube. As different alleles of each amplicon had nearly identical sequences with similar amplification properties, relative allelic counts of each target were expected to reserve in each amplicon product. For real plasma cfDNA samples, which was inherently noisy, it may be necessary to optimize amplification conditions to improve detection accuracy, such as using unique molecular identifiers^26^ or cSMART techniques^11^. For abnormality detection, nearly all amplification products were used for our method and therefore it should be cost-effective, while the WGS-based approaches used only a fraction of reads mapped to the target chromosomal regions.

In principle, target amplicon sequencing could be applied to detect other genetic variations as well. For examples, chromosomal inversion or translocation with known break point could be detected by amplicons covering the specific breakpoint. Genomic abnormalities for preimplantation embryos or non-pregnant samples could also be detected using assays similar to the reported approach, as distinct allelic distributions for all target polymorphic sites were informative enough to identify different abnormalities (Fig. S22). For cfDNA sample from a surrogate mother, fetal fraction was first estimated and updated iteratively using relative allelic counts of polymorphic sites and allelic goodness-of-fit test (Fig. S23), then genetic variations could be detected by checking all possible genotype models. For samples from a mother with multiple pregnancies, fetal fraction for each fetus could be estimated using a similar approach (Fig. S23), whereas each fetal fraction estimate was updated iteratively until converge, and genetic abnormalities could be detected using allelic goodness-of-fit test, as expected allelic counts for each polymorphic site could be calculated when fetal fractions for all fetuses were available.

Collectively, we developed a new assay, GGAP-NIPT, for detecting fetal genetic abnormalities noninvasively at both chromosomal/subchromosomal and nucleotide levels with demonstrated accuracy for simulated samples in a cost-effective manner. Such an assay showed the potential to facilitate the expansion of NIPT to detect both genetic conditions that were common to all pregnancies and disorders that had high prevalence in particular groups, which would have great socioeconomic benefits.

## Data Availability

All data are provided upon request to.

## Acknowledgements

The author would like to thank Yongxin Ke for technical and administrative assistance, discussions and comments on the project.

## Data Availability Statement

All data in the manuscript was available upon request.

## Author Contributions

S. G. designed the experiments, performed the experiments, analyzed the data and wrote the manuscript.

