## Supplementary Information for "Noninvasive Detection of Fetal Genetic Variations through Polymorphic Sites Sequencing of Maternal Plasma DNA"

**Supplementary Figures**

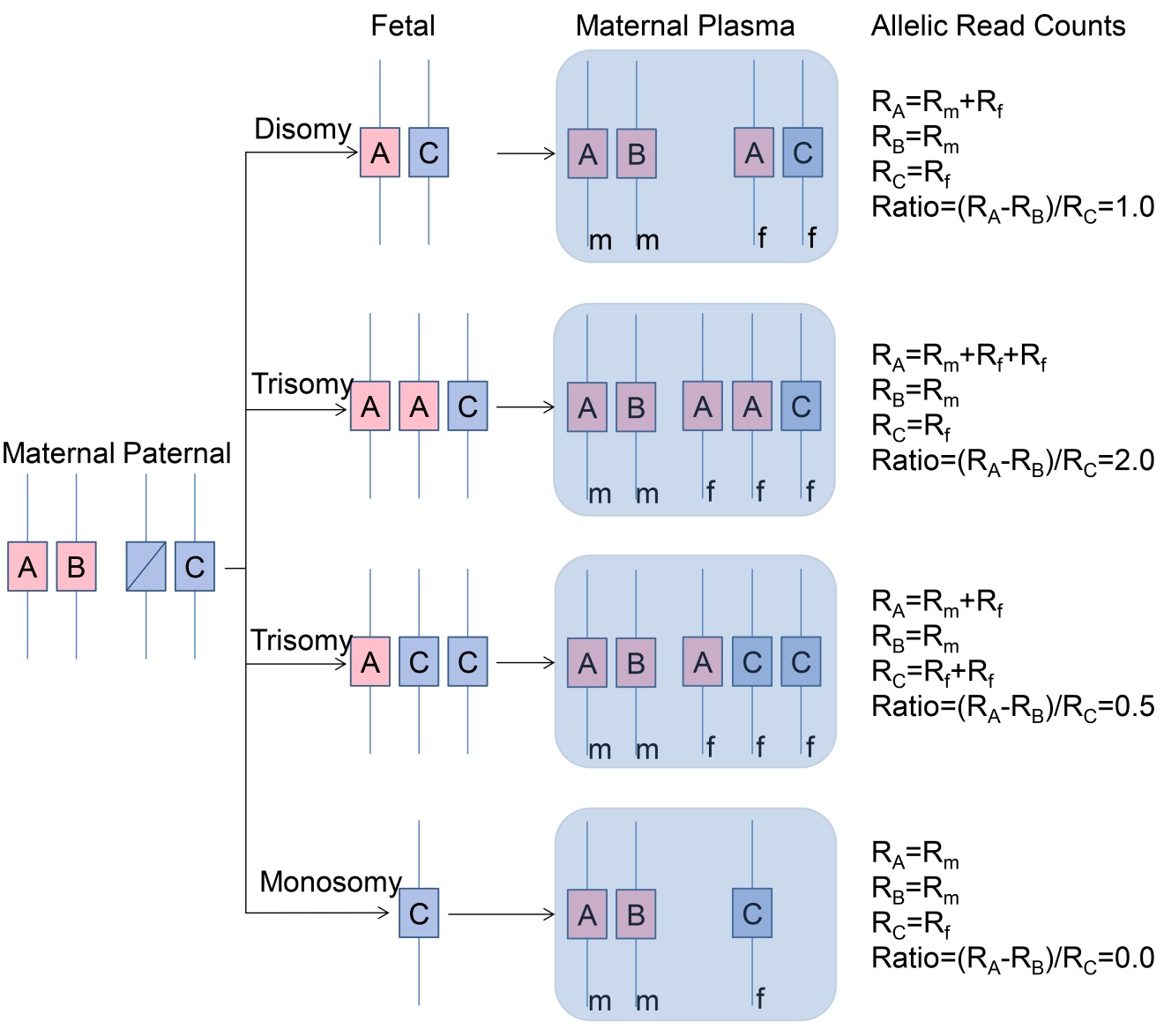

**Fig. S1. Imbalance of allelic read counts for a polymorphic site from a heterozygous mother with a fetus inheriting a distinct allele from the father.** A, B and C: distinct alleles for a polymorphic site; m and f: maternal and fetal genomic material; R_A_, R_B_ and R_C_: allelic read counts for alleles A, B and C, respectively.

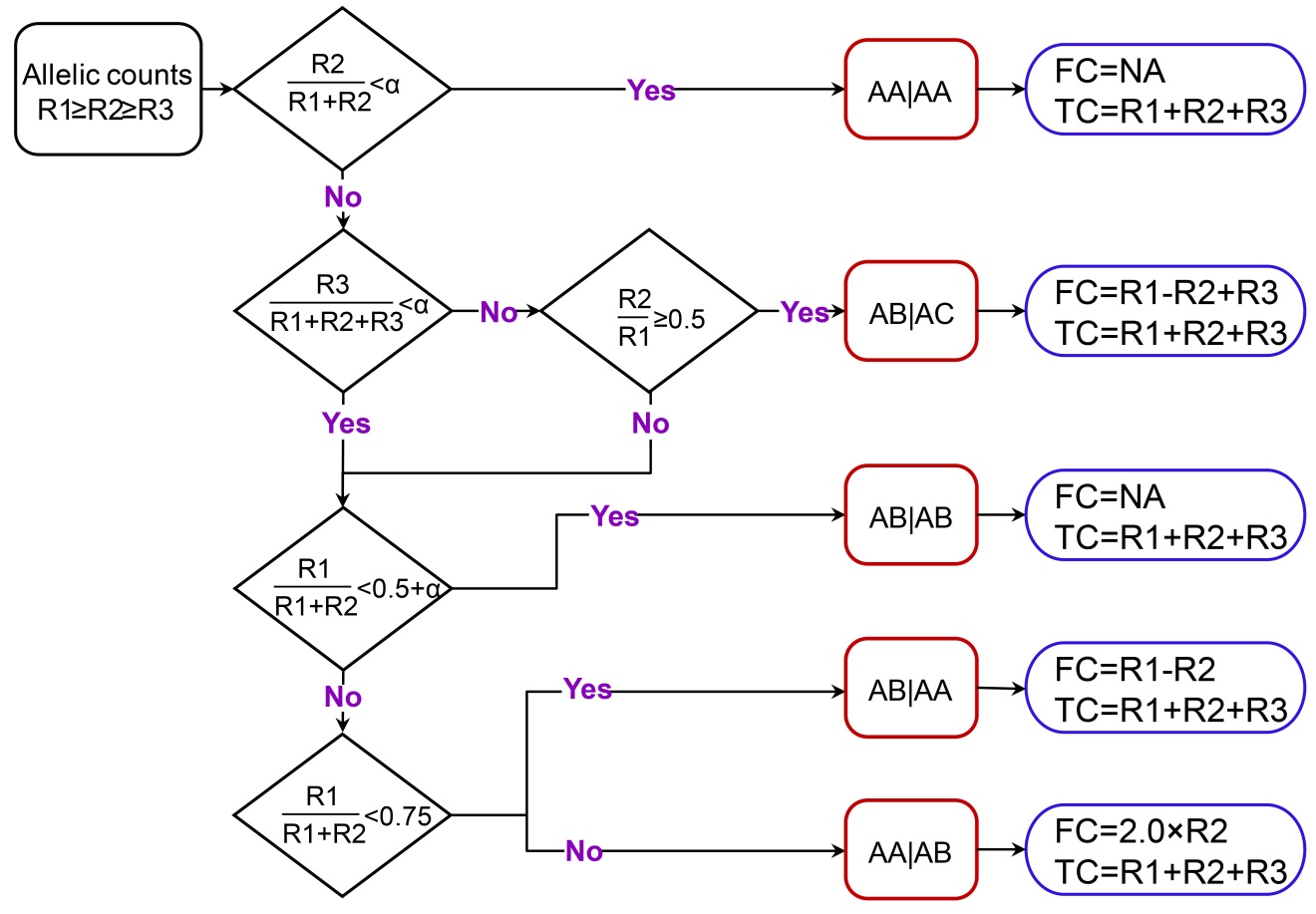

**Fig. S2. Estimating the maternal-fetal genotype of a polymorphic site using its allelic read counts.** R1, R2 and R3: allelic read counts in descending order; α: background noise threshold. A, B and C are distinct alleles for each polymorphic site, and the portion before the vertical bar denotes the maternal genotype and the part after the vertical bar denotes the fetal genotype. FC: estimated reads count amplified from fetal genetic materials (Fetal Reads); TC: total reads count amplified from both maternal and fetal genetic materials (Total Reads).

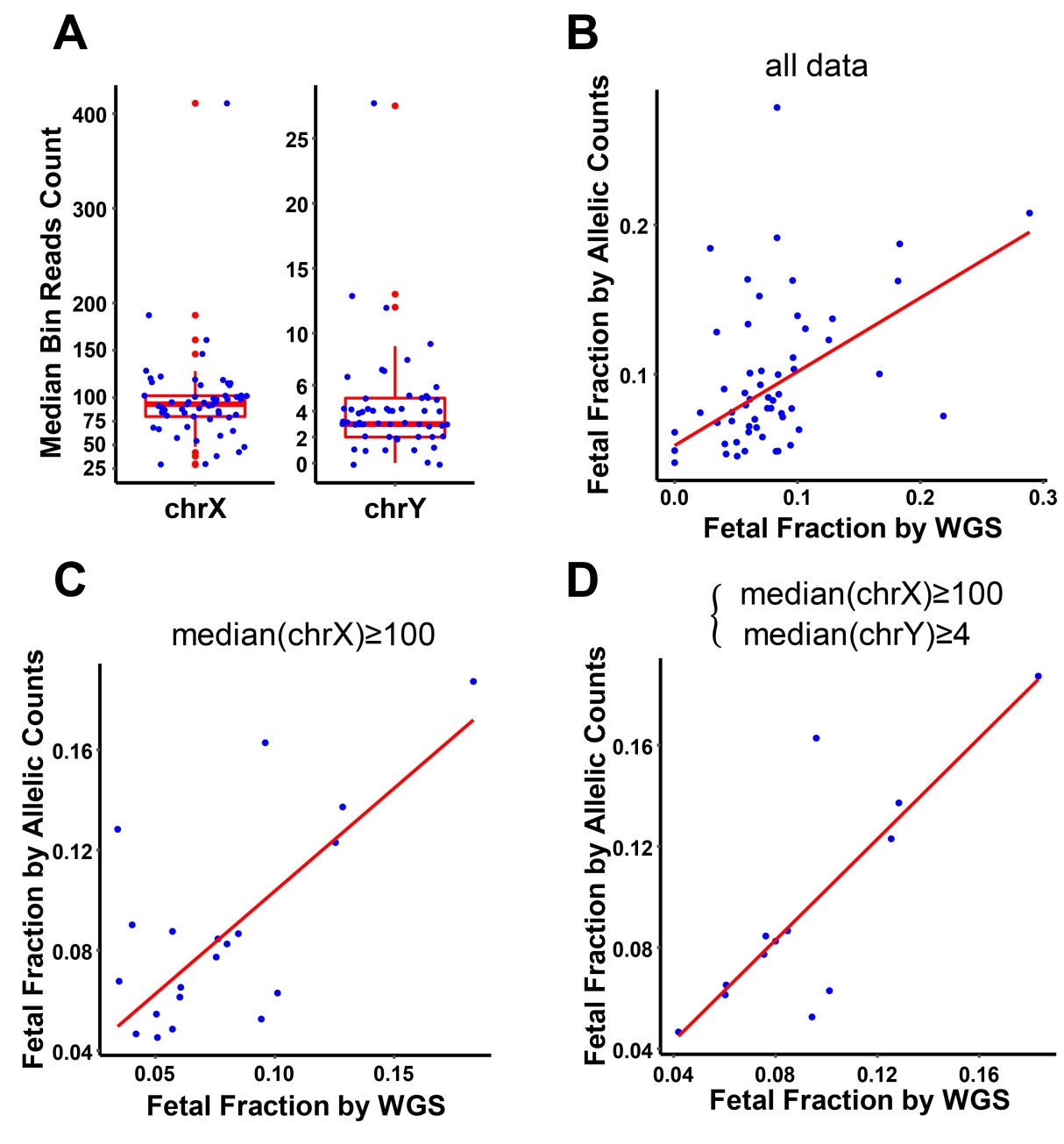

**Fig. S3. Fetal fraction estimation using allelic read counts or whole genome sequencing.** (A) Median bin read counts for WGS dataset of the insertion/deletion polymorphism samples. (B-D) Fetal fractions were estimated for each sample by both allelic read counts and WGS methods, and their relationship was plotted (red line was the fitted regression line y~x; B: all samples; C: excluding WGS samples with median bin read counts of chromosome X<100; D: same as Fig. 1E, included here for comparison purposes, whereas WGS samples with median bin counts of chrX<100 and chrY<4 were excluded). WGS: whole genome sequencing.

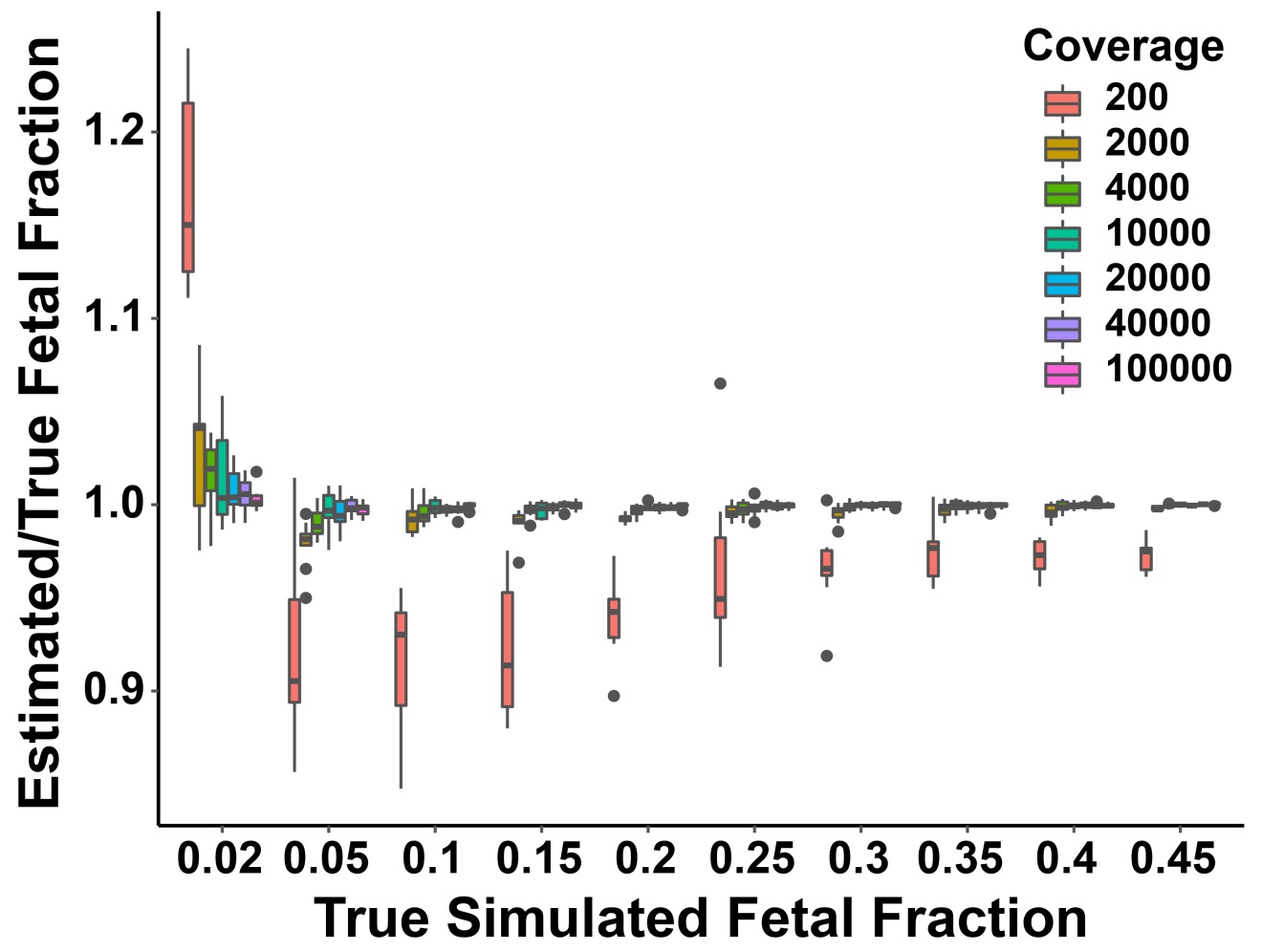

**Fig. S4. Estimation accuracy for fetal fractions of simulated samples.** One hundred samples were simulated for each sequencing coverage, and 100 polymorphic sites were simulated for each sample. At each polymorphic site, allelic sequences for one of the five disomic-disomic genotypes were randomly generated with different fetal fractions. Fetal fraction for each sample was estimated using allelic reads counts. The ratio of the estimated fetal fraction to the true fetal fraction (the simulated value) was plotted and grouped by sequencing coverage.

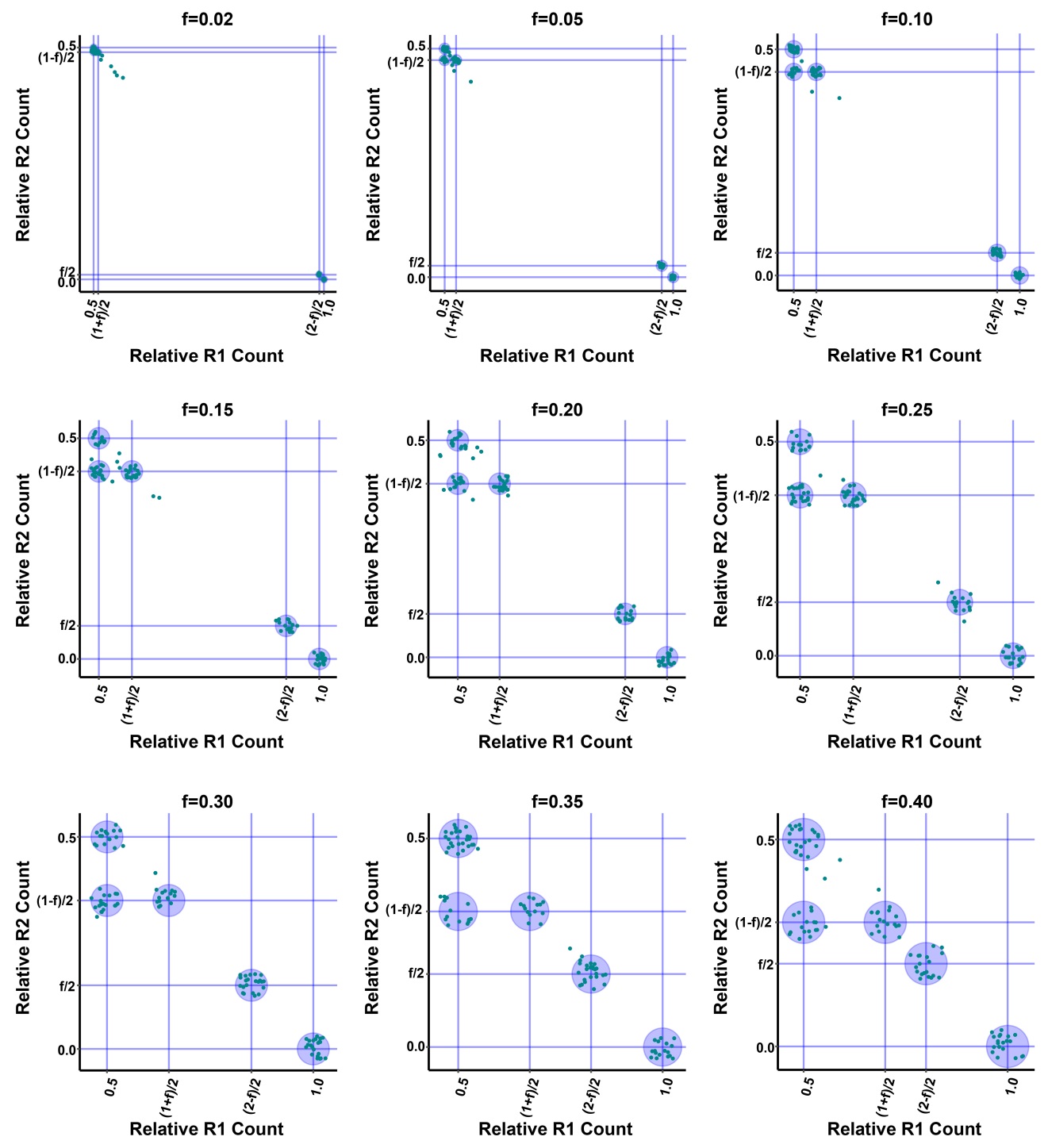

**Fig. S5. Distribution plots of relative allelic counts.** One hundred polymorphic sites on a disomy-disomy chromosome were simulated for each sample. For each polymorphic site in a sample, relative allelic read counts were calculated, and then the relative R2 count was plotted against the relative R1 count. One representative plot was shown for each fetal fraction. f: fetal fraction. Relative R1 Count=R1/(R1+R2+R3) and Relative R2 Count=R2/(R1+R2+R3).

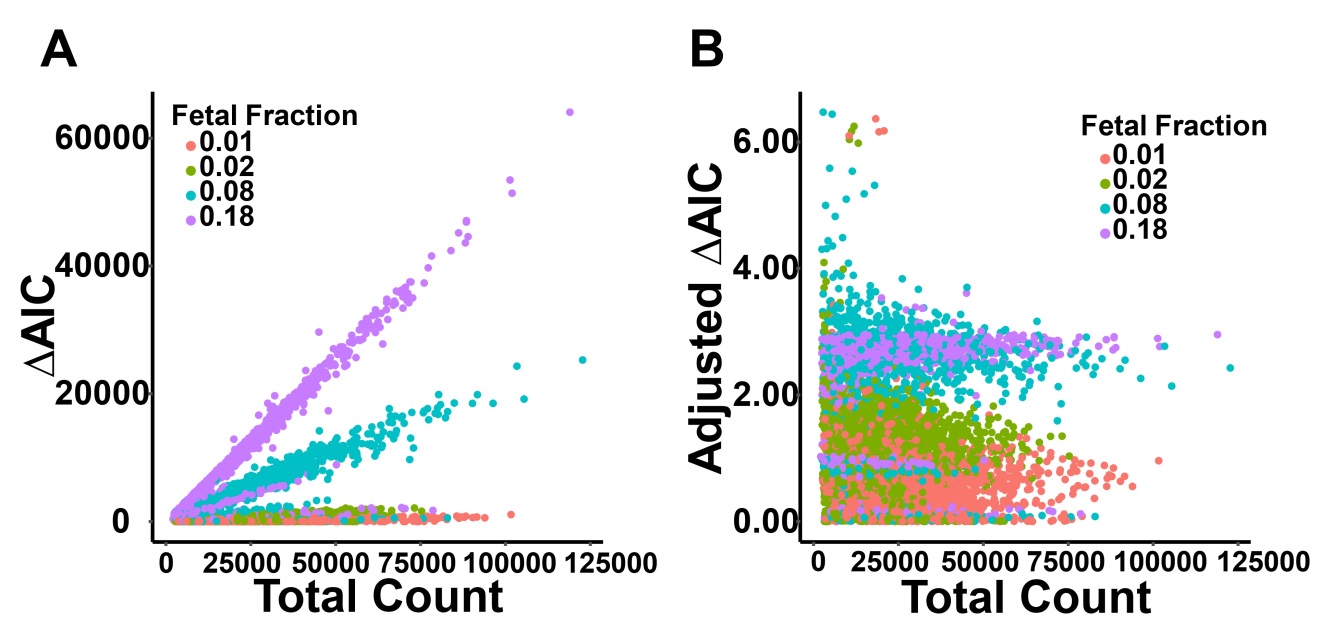

**Fig. S6. Influence of fetal fraction and total allelic read count on ΔAIC.** Fetal fraction was estimated for each sample in the replicates dataset and rounded to the second decimal place. (A) ΔAIC was calculated for each polymorphic site of each replicate sample using the normal disomy-disomy model, plotting against total allelic read count grouped by the estimated fetal fraction. (B) Adjusted ΔAICs were calculated for the replicate samples and plotted against total allelic read counts.

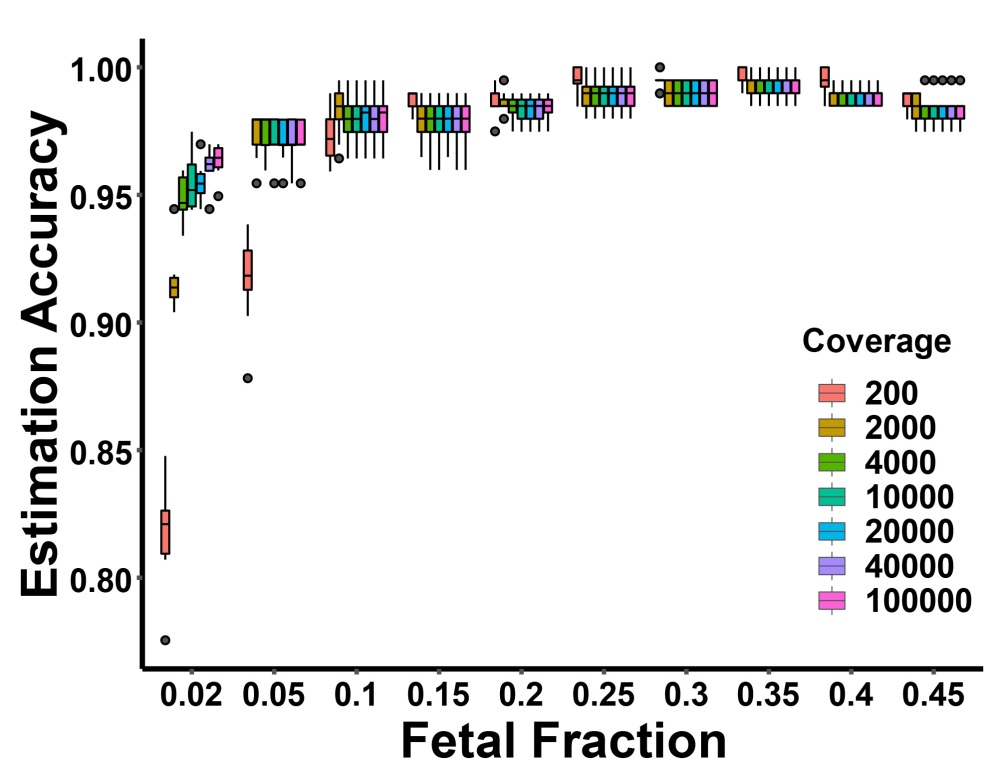

**Fig. S7. Influence of fetal fraction on maternal-fetal genotype estimation.** Sequencing reads were simulated for samples with different fetal fractions and different sequencing coverage, and fetal fraction was estimated for each sample followed with genotype estimation for each polymorphic site. Estimation accuracy was calculated as the ratio of the number of correctly estimated genotypes to the total number of all polymorphic sites, plotting against fetal fractions grouped by sequencing coverage.

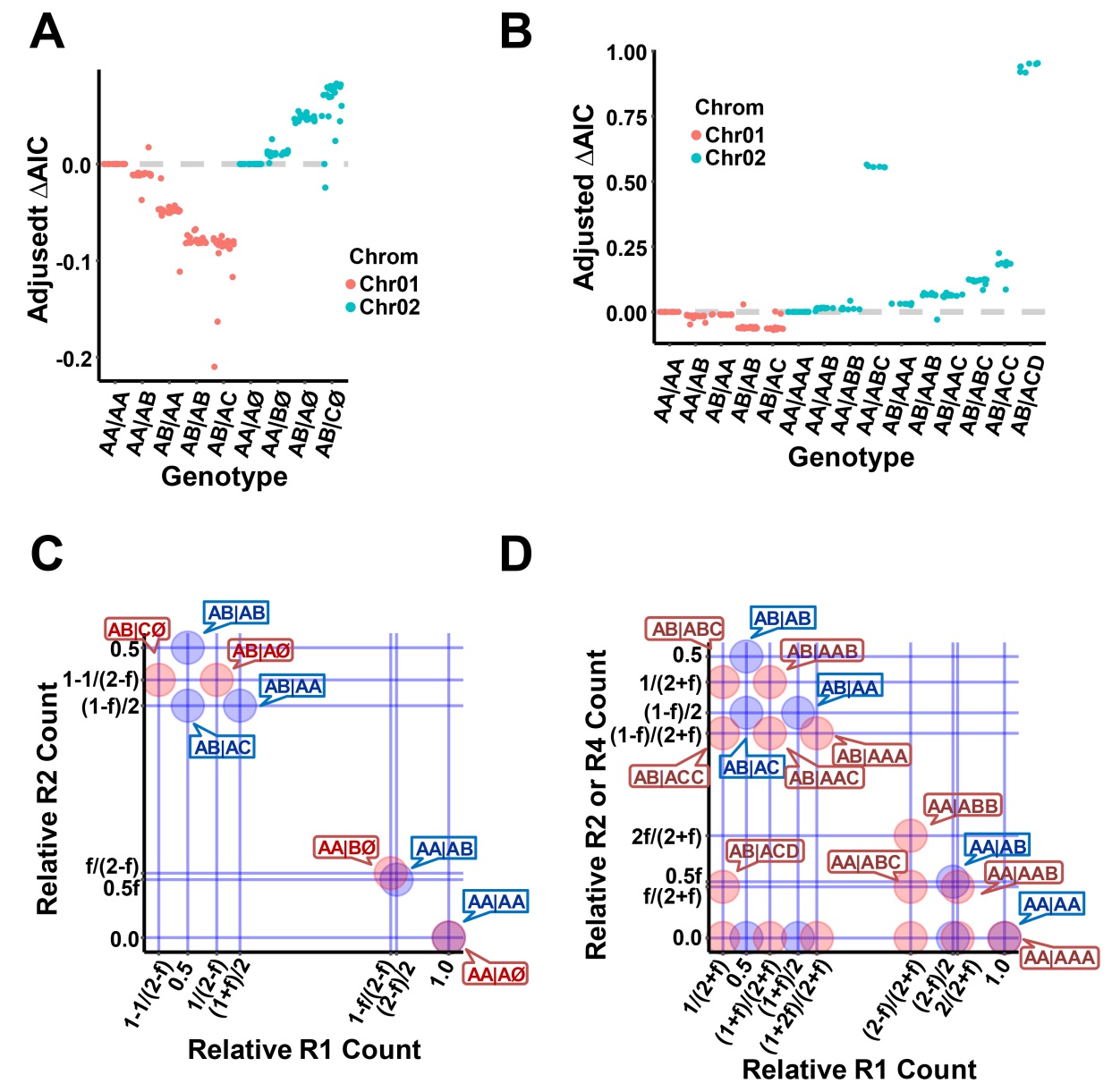

**Fig. S8. Detection of chromosomal aneuploidies.** (A) Contributions of different genotypes to the detection of fetal monosomy for two simulated chromosomes (Chr01, Di.Di; Chr02, Di.Mo). Adjusted ΔAIC= the minimal adjusted AIC for Di.Di model - the minimal adjusted AIC for Di.Mo model. (B) Contributions of different genotypes to the detection of fetal trisomy for two simulated chromosomes (Chr01, Di.Di; Chr02, Di.Tri). Adjusted ΔAIC= the minimal adjusted AIC for Di.Di model - the minimal adjusted AIC for Di.Tri model. (C) Expected relative allelic clusters of polymorphic sites for fetal monosomy detection. (D) Expected relative allelic clusters of polymorphic sites for fetal trisomy detection.

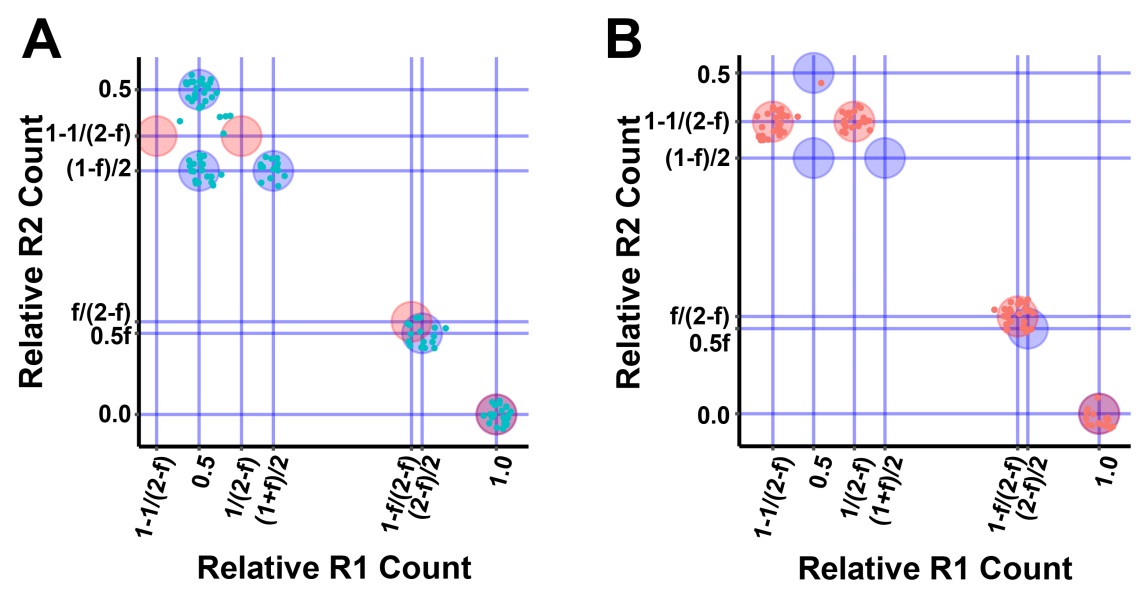

**Fig. S9. Detection of chromosomal monosomy.** (A) Relative allelic count plot for simulated polymorphic sites on a representative target chromosome. From the characteristic cluster positions, the target chromosome was estimated to be disomy-disomy. (B) Relative allelic count plot for simulated polymorphic sites on a representative target chromosome. From the characteristic cluster positions, the target chromosome was estimated to be disomy-monosomy. f: fetal fraction.

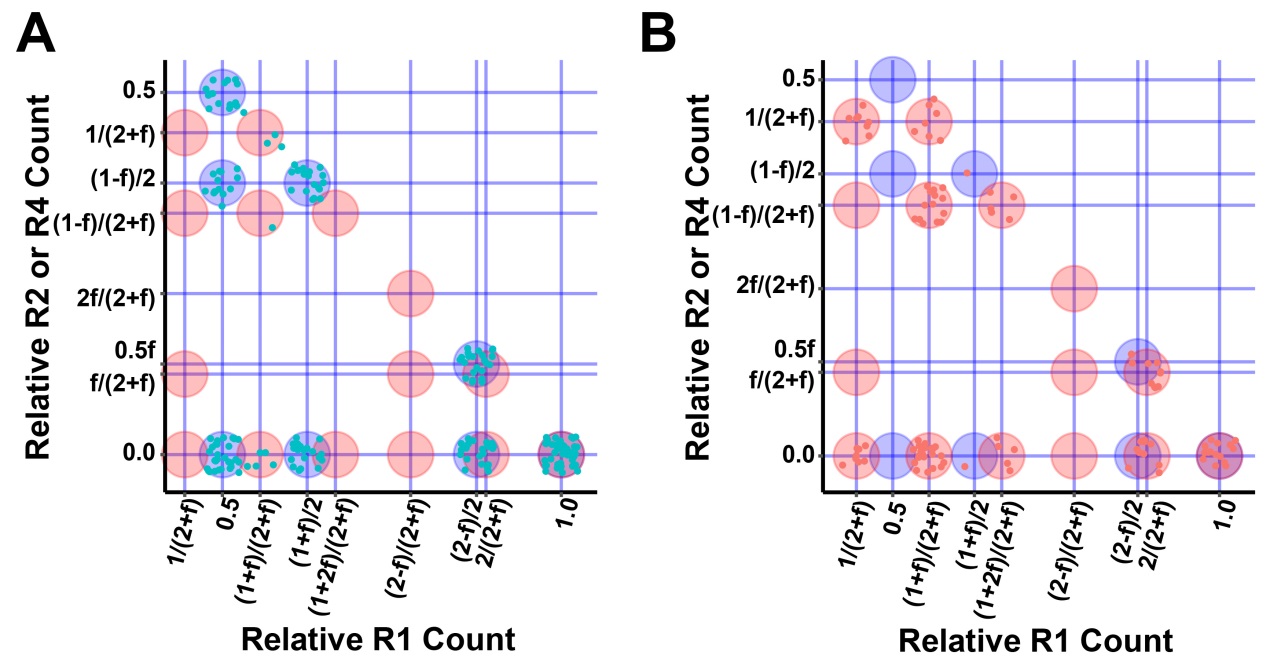

**Fig. S10. Detection of chromosomal trisomy.** (A) Relative allelic count plot for simulated polymorphic sites on a representative target chromosome. From the characteristic cluster positions, the target chromosome was estimated to be disomy-disomy. (B) Relative allelic count plot for simulated polymorphic sites on a representative target chromosome. From the characteristic cluster positions, the target chromosome was estimated to be disomy-trisomy.

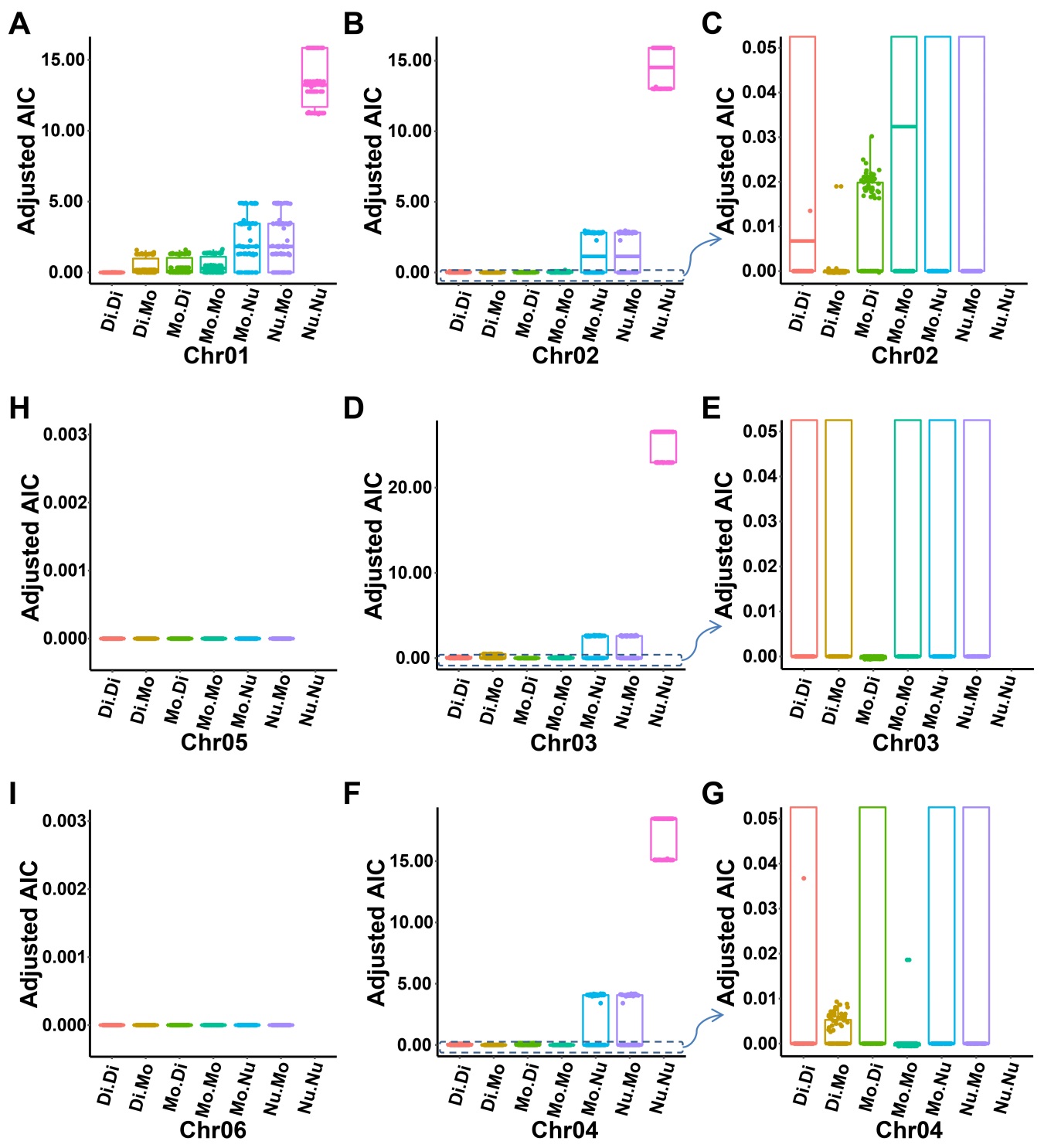

**Fig. S11. Detection of subchromosomal microdeletion.** Maternal-fetal subchromosomes (labeled as Chr01-Chr06) were simulated, and each having 100 polymorphic sites. Chr01-Chr06 subchromosomes were simulated as Di.Di, Di.Mo, Mo.Di, Mo.Mo, Mo.Nu and Nu.Mo, respectively. Allelic read counts of each polymorphic site was tested against all possible genotypes assuming each one of the seven maternal-fetal subchromosomal microdeletion models, and the overall best fitted model for all target polymorphic sites was selected for each subchromosome. (A,B,D,F,H,I) Overall fitted results for each subchromosome. (C,E,G) Partial enlarged drawings of B, D and F, respectively. Di: homozygous normal; Mo: heterozygous microdeletion; Nu: homozygous microdeletion.

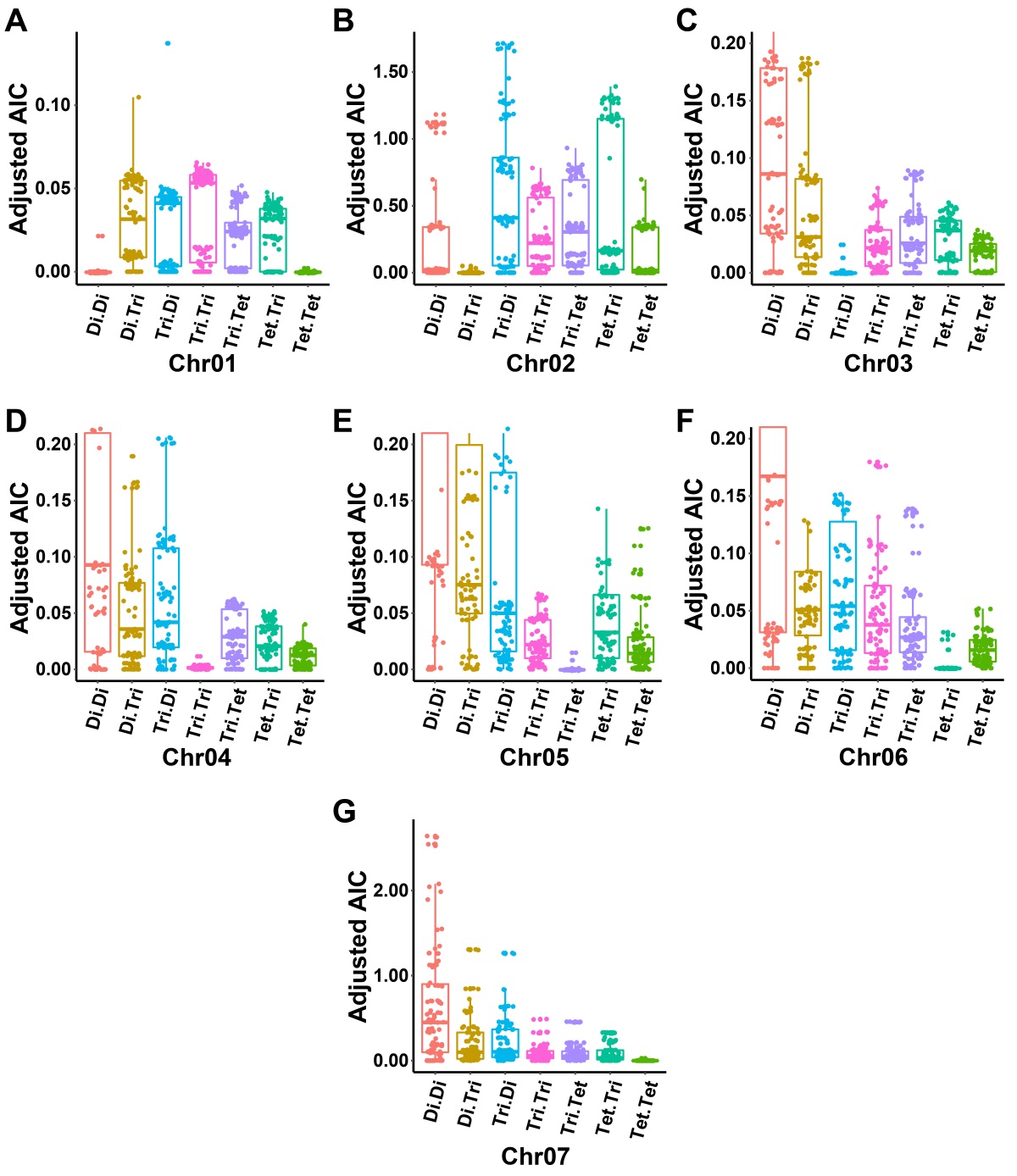

**Fig. S12. Detection of subchromosomal microduplication.** Maternal-fetal subchromosomes (labeled as Chr01-Chr07) were simulated, and each having 100 polymorphic sites. Chr01-Chr07 subchromosomes were simulated as Di.Di, Di.Tri, Tri.Di, Tri.Tri, Tri.Tet, Tet.Tri and Tet.Tet, respectively. Allelic read counts of each polymorphic site was tested against all possible genotypes assuming each one of the seven maternal-fetal subchromosomal microduplication models and the overall best fitted model for all target polymorphic sites was selected for each target. (A,B,G) Overall fitted results for subchromosomes Chr01, Chr02 and Chr07, respectively. (C,D,E,F) Partial enlarged drawings of overall fitted results for subchromosomes Chr03, Chr04, Chr05 and Chr06, respectively. Di: homozygous normal; Tri: heterozygous microduplication; Tet: homozygous microduplication.

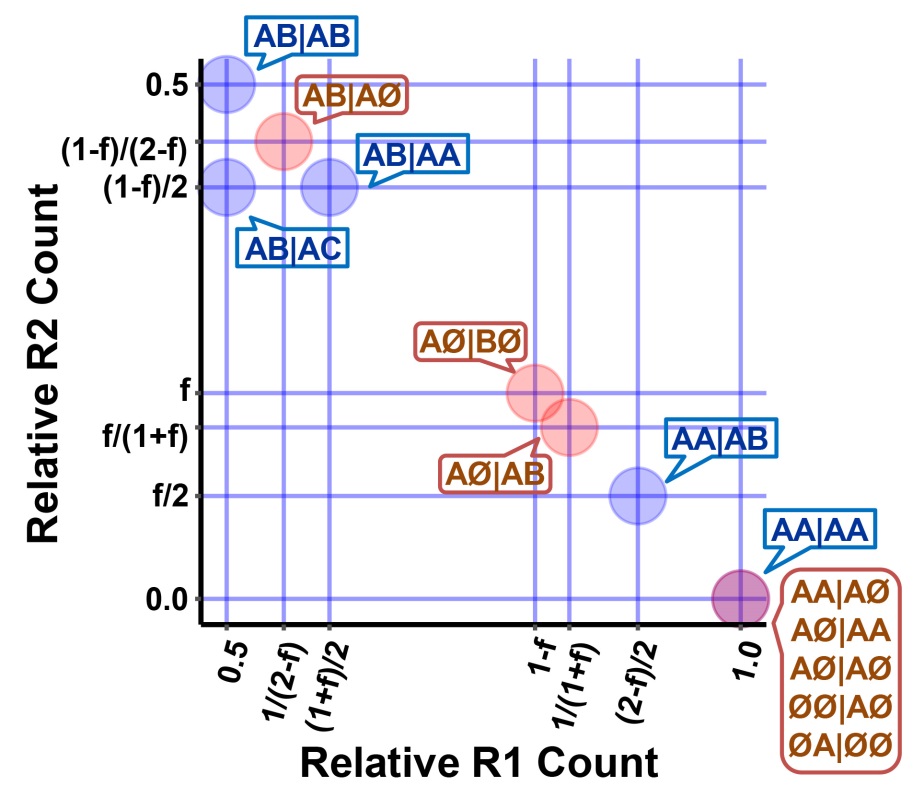

**Fig. S13. Expected relative allelic clusters of polymorphic sites for all possible subchromosomal microdeletion genotypes.** A-C: different alleles; Ø: null.

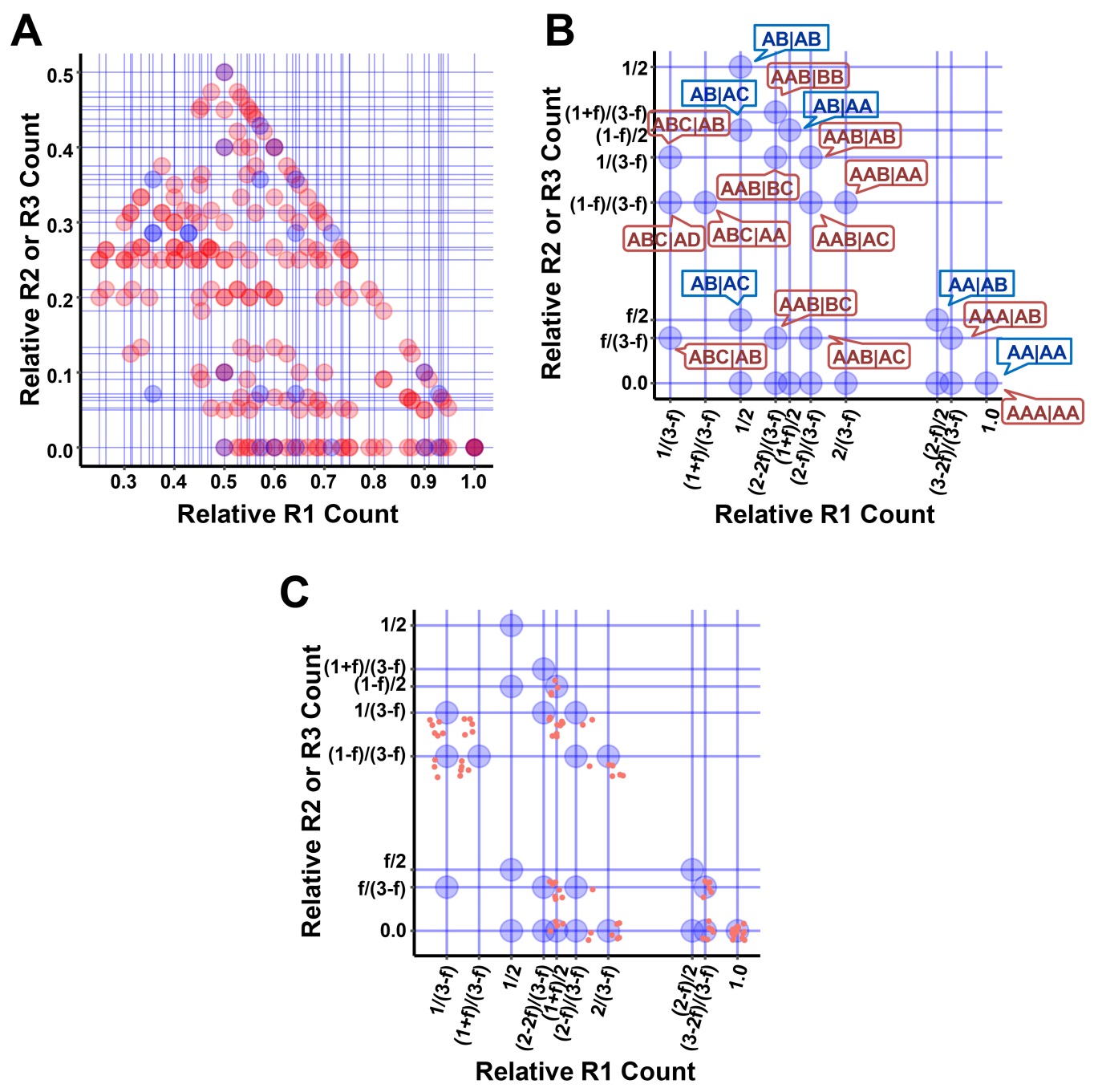

**Fig. S14. Expected relative allelic clusters of polymorphic sites for microduplication genotypes.** (A) Clusters for all possible microduplication genotypes. (B) Clusters for microduplication genotypes that are normal for the fetus. (C) Detecting maternal-fetal subchromosomal duplication by relative allelic cluster distributions of all target polymorphic sites. A maternal fetal trisomy-trisomy chromosome was simulated, and relative allelic counts of each simulated target polymorphic sites were plotted. Blue circle: all possible fetal disomy clusters. As there were allelic clusters not in the expected positions for a normal disomy fetus in the plot, either the fetus was abnormal for microduplications or the true and correct model was not included in the analysis.

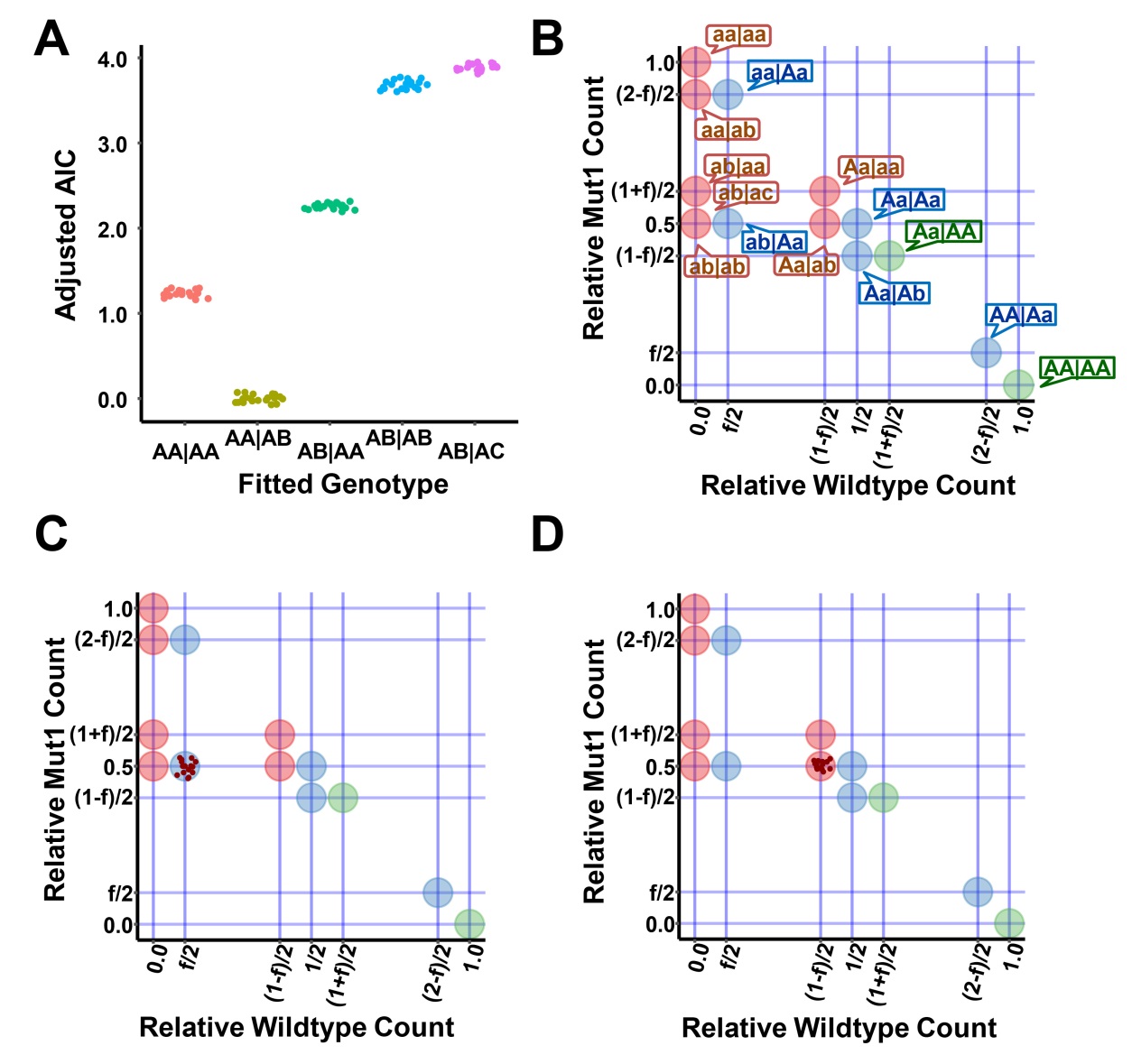

**Fig. S15. Detection of fetal short genetic variations.** (A) Detection of short genetic variation by allelic goodness-of-fit analysis for a simulated site with library-level replicates. According to the plot, AA|AB genotype was the best fit. As allele A was mutant and allele B was wildtype by sequence analysis, the target was estimated to be a homozygous mutant-mutant for the mother and a heterozygous wildtype-mutant for the fetus. (B) Expected all possible relative allelic clusters for polymorphic sites on normal disomy-disomy chromosomes (A, wildtype allele; a-c, different mutant alleles). (C,D) Detecting maternal-fetal short variation by relative allelic cluster distribution of the target site against theoretical clusters of all possible wildtype/mutant genotype models. According to the characteristic allelic positions, the simulated target site was estimated to be a heterozygous mutant-mutant for the mother and a heterozygous wildtype-mutant for the fetus (C) or a heterozygous wildtype-mutant for the mother and a heterozygous mutant-mutant for the fetus carrying two different mutant alleles (D).

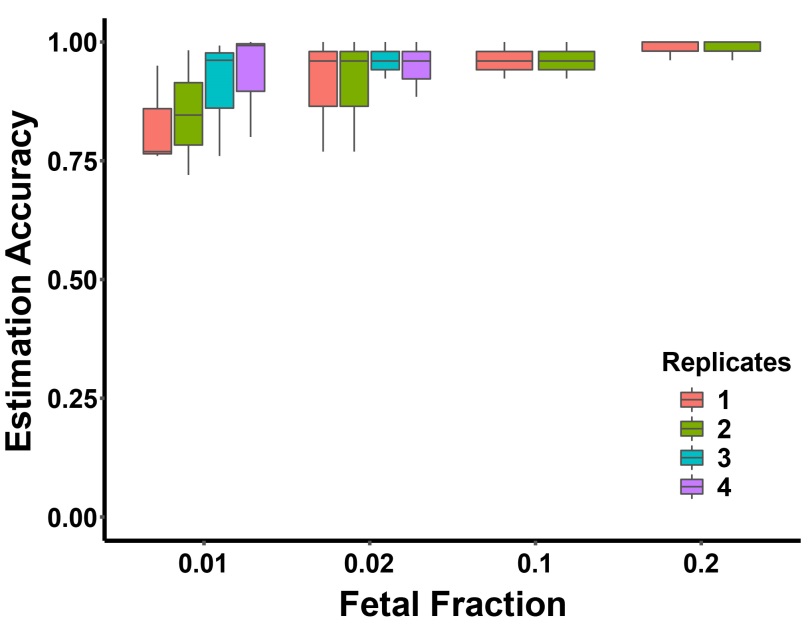

**Fig. S16. Estimation accuracy for genotypes of the replication dataset.** Genotype was estimated for each polymorphic site using its allelic read counts for each sample in the replication dataset. Estimation accuracy was calculated as the ratios of the number of correctly estimated genotypes to the total number of polymorphic sites grouped by different replicates and different fetal fractions. Replicates were labeled as 1 to 4, and ratios for replicates 1 to 4 means that 1 to 4 related samples were selected to calculate the estimation accuracy, respectively.

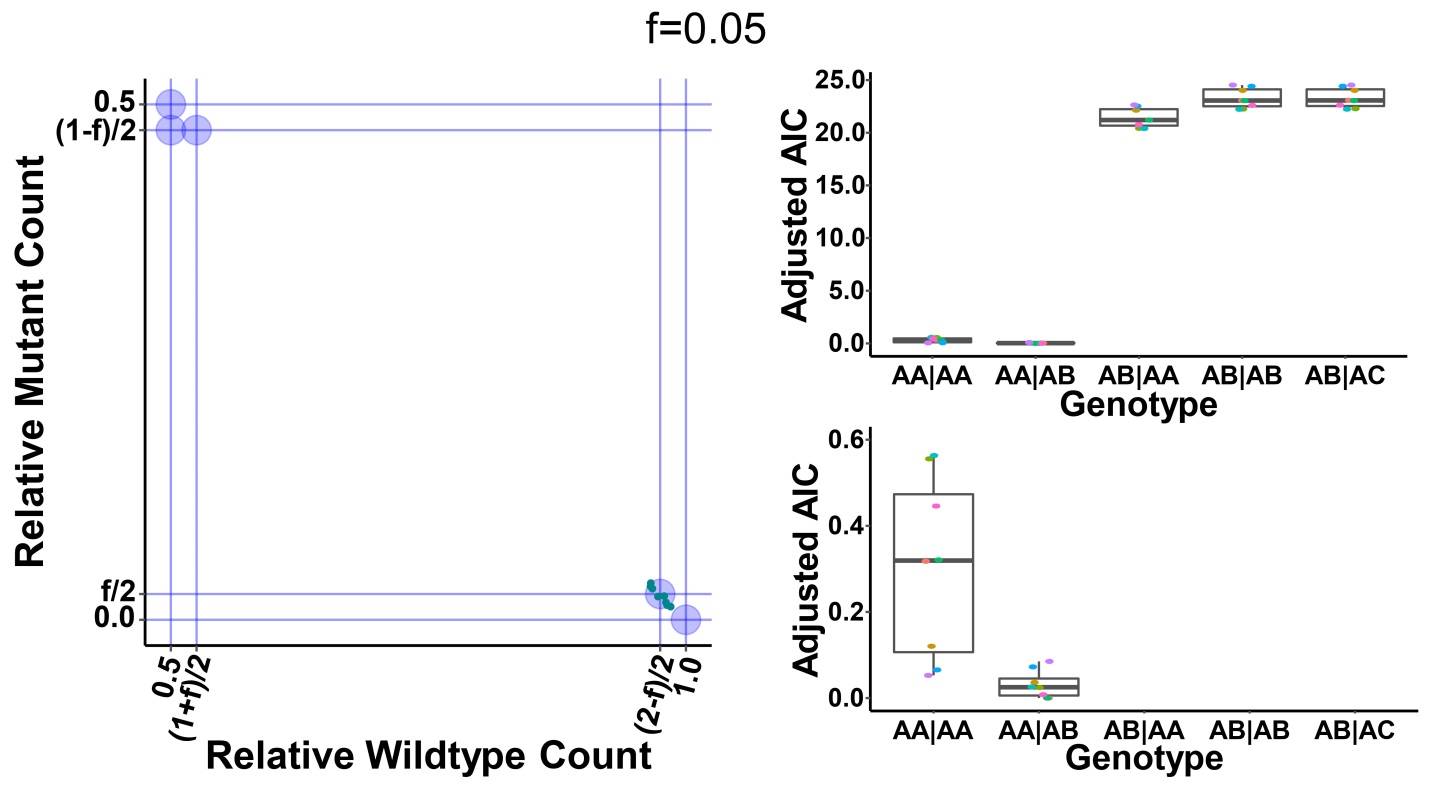

**Fig. S17. Detecting fetal mutations for the wilson validation dataset.** Maternal and paternal plasma samples were mixed, mimicking homozygous-heterozygous maternal-fetal genotypes with a fetal fraction of 5% (8 replicates). Maternal-fetal genotype was estimated using relative allelic counts plot (left) or goodness-of fit test (right). From the left plot, the maternal-fetal genotype was estimated as homozygous wildtype-heterozygous mutant. From the right plot, the maternal-fetal genotype was estimated to be AA|AB. As wildtype counts were the major components, A was a wildtype allele and B was a mutant allele, hence the mother was wildtype and the fetus was heterozygous mutant.

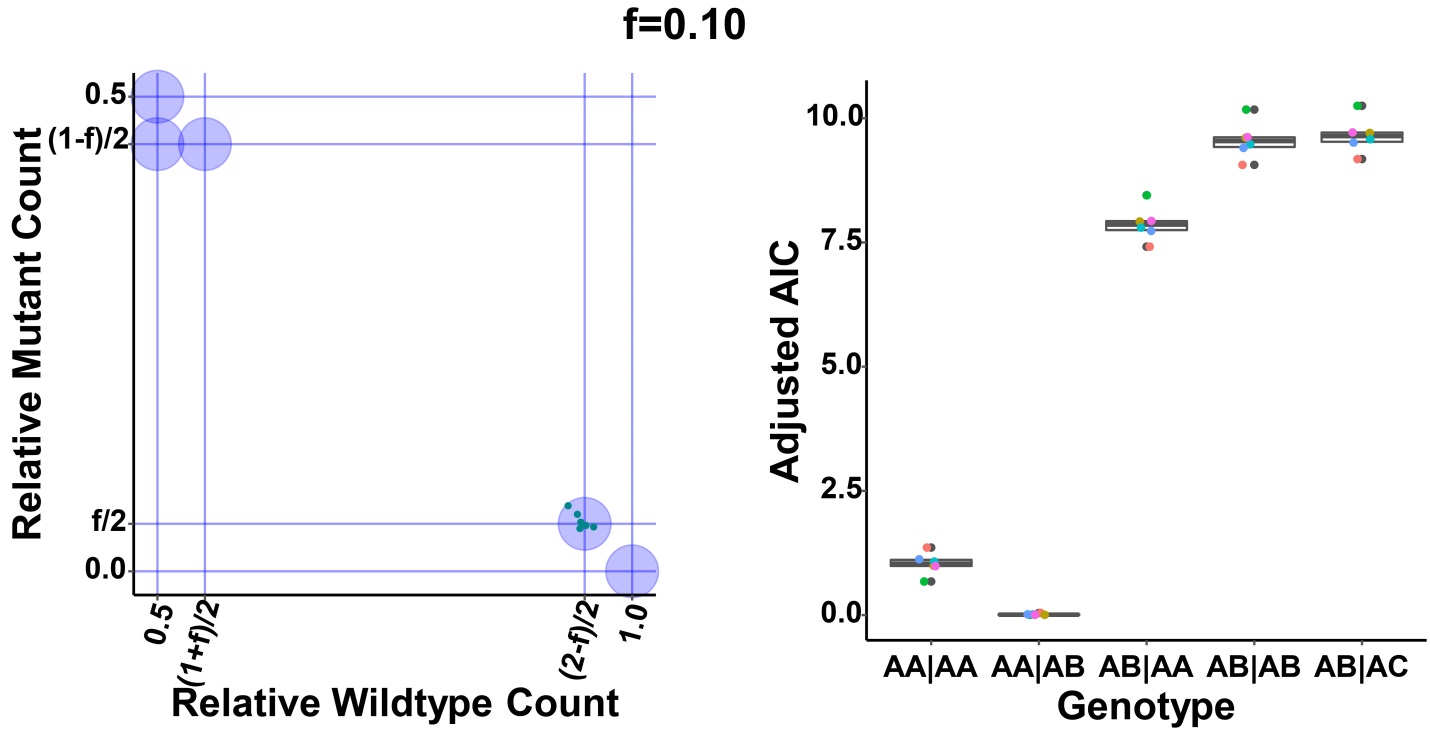

**Fig. S18. Detecting fetal mutations for the wilson validation dataset.** Maternal and paternal plasma samples were mixed, mimicking homozygous-heterozygous maternal-fetal genotypes with a fetal fraction of 10% (6 replicates). Maternal-fetal genotype was estimated using relative allelic counts plot (left) or goodness-of fit test (right). From the left plot, the maternal-fetal genotype was estimated as homozygous wildtype-heterozygous mutant. From the right plot, the maternal-fetal genotype was estimated to be AA|AB. As wildtype counts were the major components, A was a wildtype allele and B was a mutant allele, hence the mother was wildtype and the fetus was heterozygous mutant.

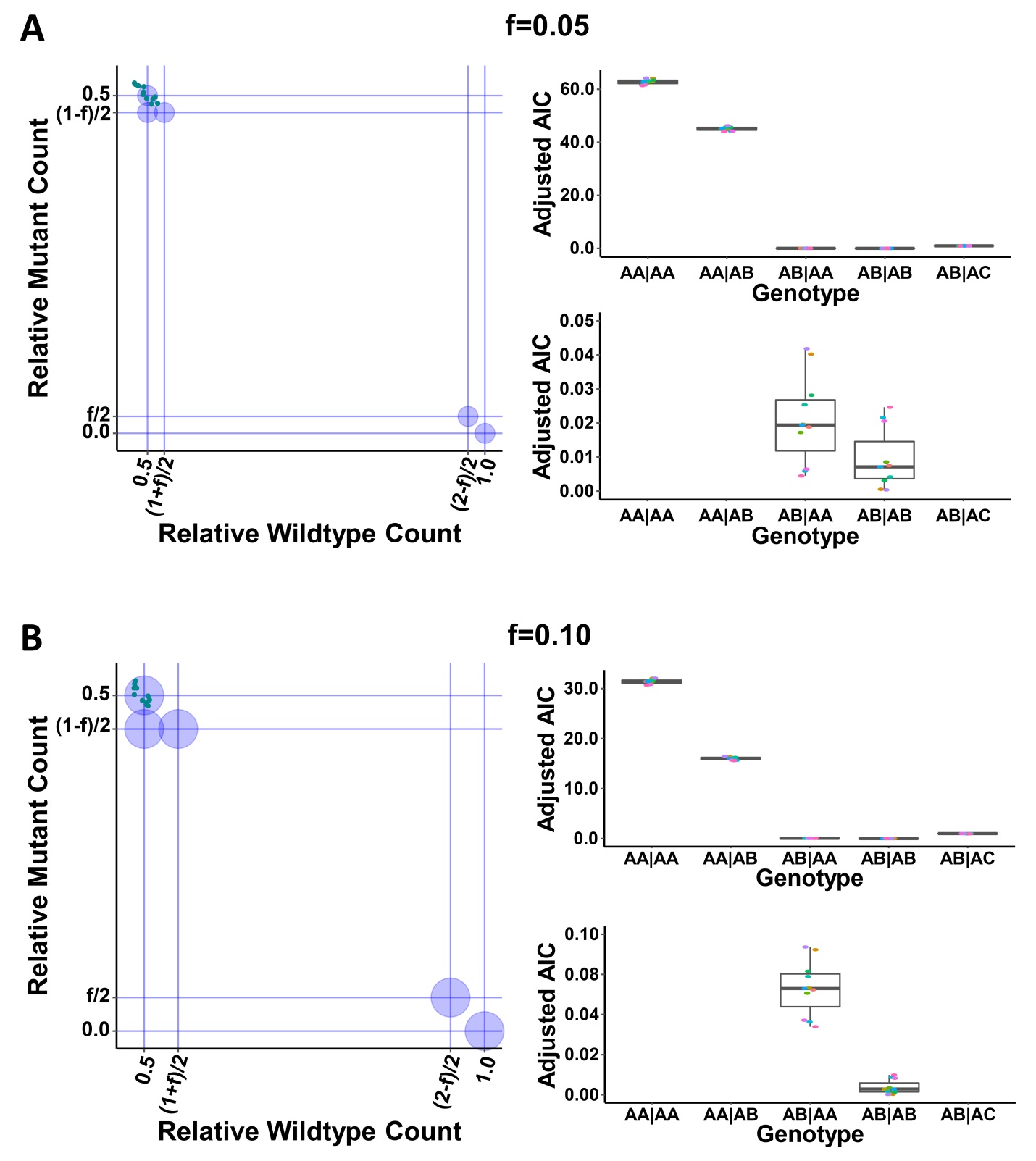

**Fig. S19. Detecting fetal mutations for the wilson validation dataset.** Plasma samples from 11 pregnant women were prepared, whereas both the mothers and the fetuses were heterozygous mutant. As fetal fractions were not reported for 6 of the 11 samples, a marginal (f=0.05, A) and moderate (f=0.10, B) fetal fraction values were assumed. Maternal-fetal genotype was estimated using relative allelic counts plot (left) or goodness-of fit test (right). From the left plots, the maternal-fetal genotype was estimated as heterozygous mutant-heterozygous mutant. From the right plots, the maternal-fetal genotypes were estimated to be AB|AB, hence both the mothers and the fetuses were heterozygous mutant.

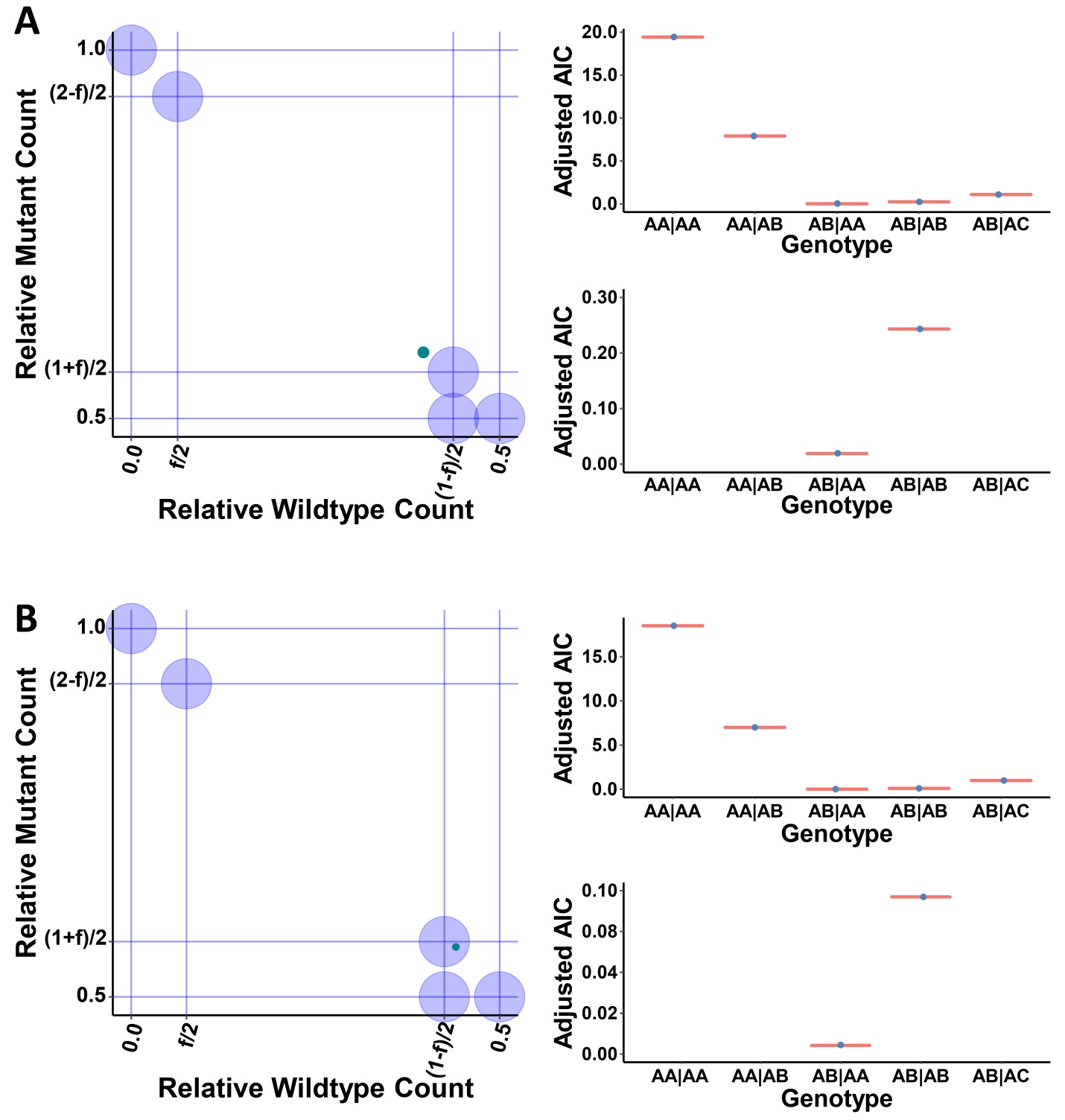

**Fig. S20. Detecting fetal mutations for the wilson validation dataset.** Plasma samples from 2 pregnant women were prepared, whereas the mothers were heterozygous mutant and the fetuses were homozygous mutant. Maternal-fetal genotype was estimated for sample 1 (A) and sample 2 (B) using relative allelic counts plot (left) or goodness-of fit test (right). From the left plots, the maternal-fetal genotype was estimated as heterozygous mutant-homozygous mutant. From the right plots, the maternal-fetal genotypes were estimated to be AB|AA. As mutant counts were the major components, A was a mutant allele and B was a wildtype allele, hence the mothers were heterozygous mutant and the fetuses were homozygous mutant.

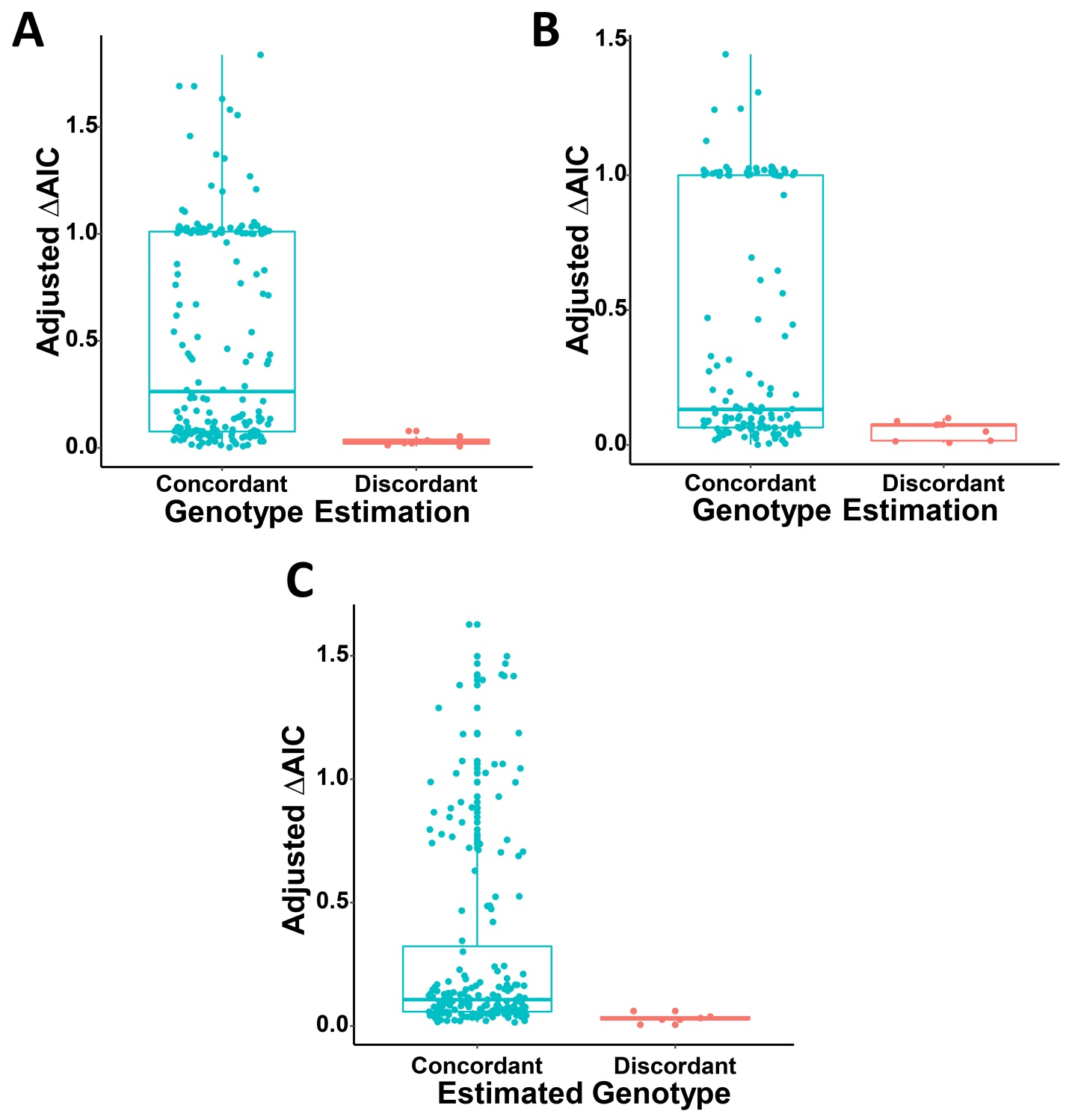

**Fig. S21. Comparisons of goodness of fit test results for target sites with correctly and incorrectly estimated genotypes.** Adjusted ∆AIC values were plotted for sites from the hbb dataset (A), arnshl dataset (B) and cfbest dataset (C), grouped by estimation concordances of maternal-fetal genotypes.

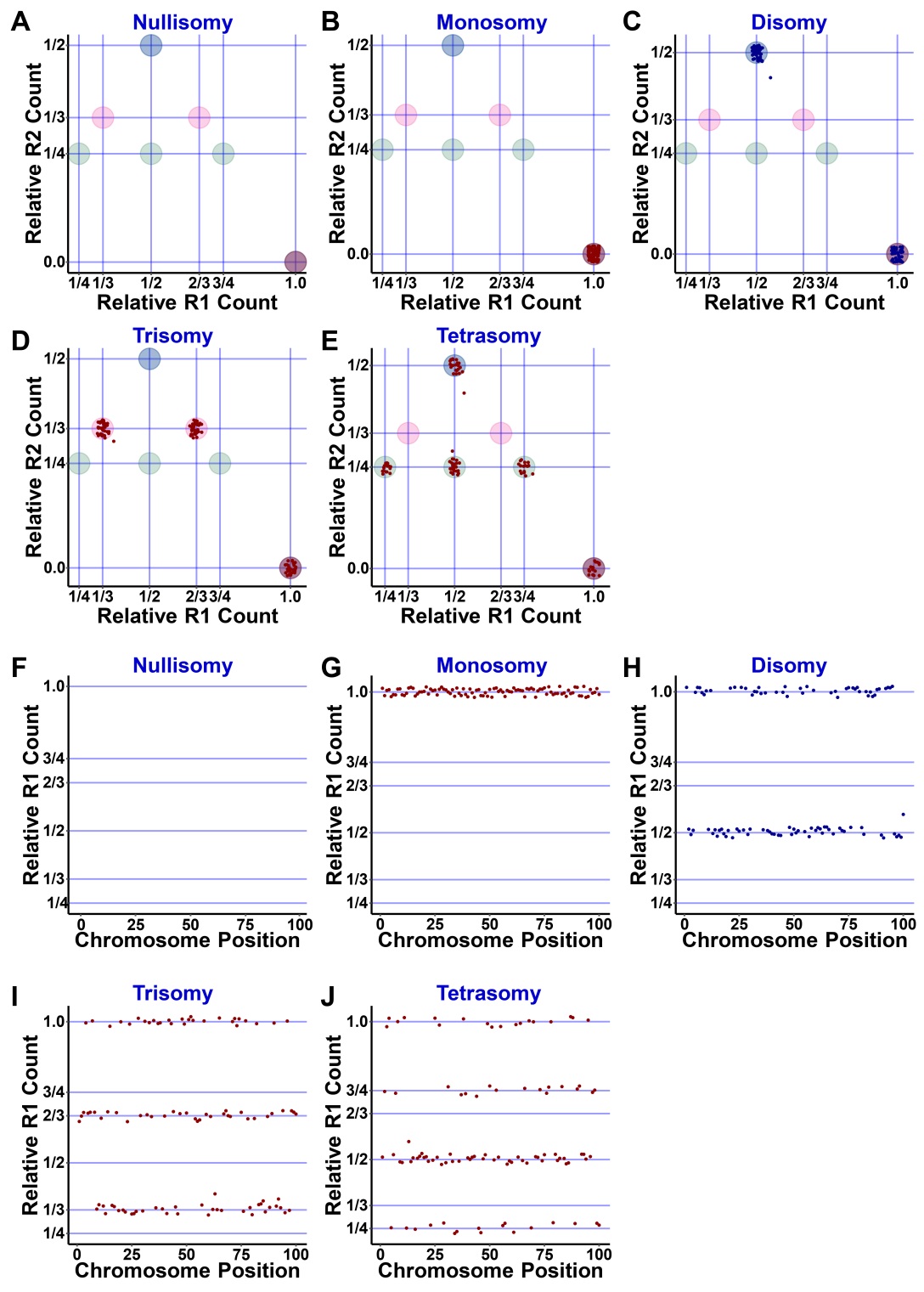

**Fig. S22. Detecting genetic aberrations for samples from non-pregnant individuals or preimplantation embryos.** A panel of polymorphic sites on the target chromosome was simulated for a normal non-pregnant individual, and relative allelic counts for each polymorphic site were calculated. Then the relative R2 count was plotted against the relative R1 count (A-E) or the relative R1 count was plotted against its relative chromosomal position (F-J) for each amplicon. (A,F) nullisomy (or homozygous microdeletion); (B,G) monosomy (or heterozygous microdeletion); (C,H) disomy (or normal); (D,I) trisomy (or heterozygous microduplication); (E,J) tetrasomy (or homozygous microduplication).

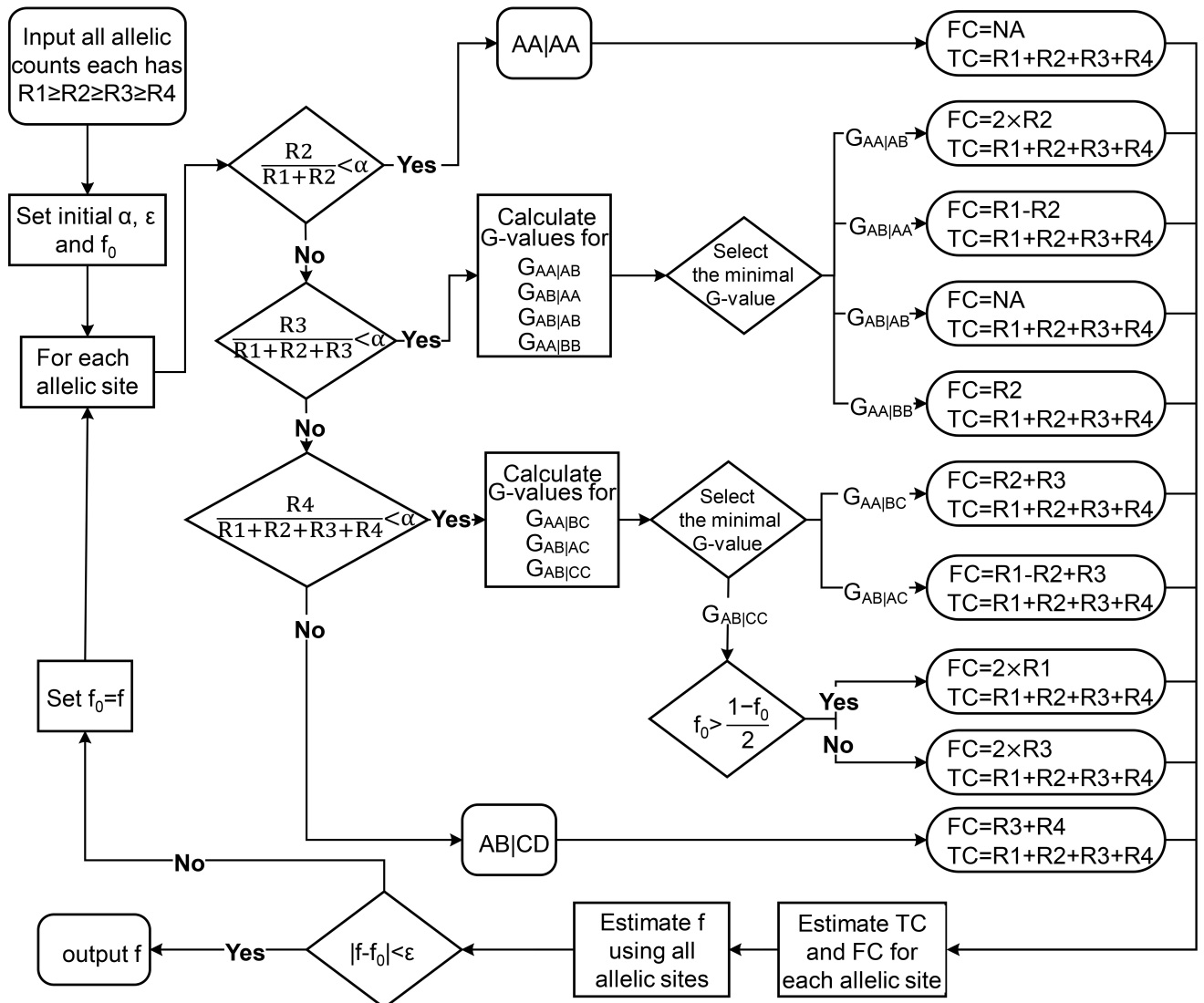

**Fig. S23. Estimating fetal fraction for a sample from a surrogate mother using allelic read counts.** R1, R2, R3 and R4: allelic read counts in descending order; α: background threshold; ε: estimation precision; f_0_: initial fetal fraction estimate; A-D: distinct alleles for each polymorphic site.

**Supplementary Tables**

| **Table S1.** Relative allelic counts for a polymorphic site on reference chromosomes | | | | | | |
| --- | --- | --- | --- | --- | --- | --- |
| Genotype | Allelic Read Counts | | | R1/(R1+R2) | Estimation | |
|  | R1 | R2 | R3 |  | Fetal Reads | Total Reads |
| AA\|AA | R_m_R_m_R_f_R_f_ | 0 | 0 | 1 | NA | R1 |
| AA\|AB | R_m_R_m_R_f_ | R_f_ | 0 | 1-0.5*f∈(0.75, 1) | 2.0*R2 | R1+R2 |
| AB\|AA | R_m_R_f_R_f_ | R_m_ | 0 | 0.5+0.5*f∈(0.5, 0.75] | R1-R2 | R1+R2 |
| AB\|AB | R_m_R_f_ | R_m_R_f_ | 0 | 0.5 | NA | R1+R2 |
| AB\|AC | R_m_R_f_ | R_m_ | R_f_ | 1/(2-f)∈[0.5, 2/3] | R1-R2+R3 | R1+R2+R3 |
| 1. f: fetal fraction | | | | | | |
| 2. R1, R2 and R3: read counts of each allele sorted in descending order | | | | | | |
| 3. R_m_ and R_f_: Reads mapped to maternal and fetal chromosomes, respectively. | | | | | | |

| **Table S2.** Relative allelic count for disomy-disomy model | | | |
| --- | --- | --- | --- |
| Genotype | RRC1 | RRC2 | RRC3 |
| AA\|AA | 1 | 0 | 0 |
| AA\|AB | (2-f)/2 | f/2 | 0 |
| AB\|AA | (1+f)/2 | (1-f)/2 | 0 |
| AB\|AB | 1/2 | 1/2 | 0 |
| AB\|AC | 1/2 | (1-f)/2 | f/2 |
| 1. RRC1 (relative R1 count)=R1/(R1+R2+R3) | | | |
| 2. RRC2 (relative R2 count)=R2/(R1+R2+R3) | | | |
| 2. RRC3 (relative R3 count)=R3/(R1+R2+R3) | | | |

| **Table S3.** Relative allelic count for disomy-monosomy model | | | |
| --- | --- | --- | --- |
| Genotype | RRC1 | RRC2 | RRC3 |
| AA\|AØ | 1 | 0 | 0 |
| AA\|BØ | 1-f/(2-f) | f/(2-f) | 0 |
| AB\|AØ | 1/(2-f) | 1-1/(2-f) | 0 |
| AB\|CØ | 1-1/(2-f) | 1-1/(2-f) | f/(2-f) |
| 1. RRC1 (relative R1 count)=R1/(R1+R2+R3) | | | |
| 2. RRC2 (relative R2 count)=R2/(R1+R2+R3) | | | |
| 2. RRC3 (relative R3 count)=R3/(R1+R2+R3) | | | |

| **Table S4.** Relative allelic count for disomy-trisomy model | | | | |
| --- | --- | --- | --- | --- |
| Genotype | RRC1 | RRC2 | RRC3 | RRC4 |
| AA\|AAA | 1 | 0 | 0 | 0 |
| AA\|AAB | 2/(2+f) | f/(2+f) | 0 | 0 |
| AA\|ABB | (2-f)/(2+f) | 2f/(2+f) | 0 | 0 |
| AB\|AAA | (1+2f)/(2+f) | (1-f)/(2+f) | 0 | 0 |
| AB\|AAB | (1+f)/(2+f) | 1/(2+f) | 0 | 0 |
| AA\|ABC | (2-f)/(2+f) | f/(2+f) | f/(2+f) | 0 |
| AB\|AAC | (1+f)/(2+f) | (1-f)/(2+f) | f/(2+f) | 0 |
| AB\|ABC | 1/(2+f) | 1/(2+f) | f/(2+f) | 0 |
| AB\|ACC | 1/(2+f) | (1-f)/(2+f) | 2f/(2+f) | 0 |
| AB\|ACD | 1/(2+f) | (1-f)/(2+f) | f/(2+f) | f/(2+f) |
| 1. RRC1 (relative R1 count)=R1/(R1+R2+R3+R4) | | | | |
| 2. RRC2 (relative R2 count)=R2/(R1+R2+R3+R4) | | | | |
| 3. RRC3 (relative R3 count)=R3/(R1+R2+R3+R4) | | | | |
| 4. RRC4 (relative R4 count)=R4/(R1+R2+R3+R4) | | | | |

| **Table S5.** Relative allelic count for subchromosomal microdeletion model | | | |
| --- | --- | --- | --- |
| Genotype | | RRC1 | RRC2 |
| Di.Mo | AA\|AØ | 1 | 0 |
|  | AB\|AØ | 1/(2-f) | (1-f)/(2-f) |
| Mo.Di | AØ\|AA | 1 | 0 |
|  | AØ\|AB | 1/(1+f) | f/(1+f) |
| Mo.Mo | AØ\|AØ | 1 | 0 |
|  | AØ\|BØ | 1-f | f |
| Nu.Mo | ØØ\|AØ | 1 | 0 |
| Mo.Nu | ØA\|ØØ | 1 | 0 |
| Nu.Nu | ØØ\|ØØ | 0 | 0 |
| 1. RRC1 (relative R1 count)=R1/(R1+R2+R3) | | | |
| 2. RRC2 (relative R2 count)=R2/(R1+R2+R3) | | | |
| 3. Only heritable genotypes were listed. | | | |

| **Table S6.** Relative allelic count for subchromosomal microduplication model | | | | | |
| --- | --- | --- | --- | --- | --- |
| Genotype | | RRC1 | RRC2 | RRC3 | RRC4 |
| Trisomy-Disomy Model | AAA\|AA | 1 | 0 | 0 | 0 |
|  | AAA\|AB | (3-2f)/(3-f) | f/(3-f) | 0 | 0 |
|  | AAB\|AA | 2/(3-f) | (1-f)/(3-f) | 0 | 0 |
|  | AAB\|AB | (2-f)/(3-f) | 1/(3-f) | 0 | 0 |
|  | AAB\|BB | (2-2f)/(3-f) | (1+f)/(3-f) | 0 | 0 |
|  | AAB\|AC | (2-f)/(3-f) | (1-f)/(3-f) | f/(3-f) | 0 |
|  | AAB\|BC | (2-2f)/(3-f) | 1/(3-f) | f/(3-f) | 0 |
|  | ABC\|AA | (1+f)/(3-f) | (1-f)/(3-f) | (1-f)/(3-f) | 0 |
|  | ABC\|AB | 1/(3-f) | 1/(3-f) | f/(3-f) | 0 |
|  | ABC\|AD | 1/(3-f) | (1-f)/(3-f) | (1-f)/(3-f) | f/(3-f) |
| Other Genotypes | | … | … | … | … |
| 1. Only the model having normal fetal genotypes is listed. | | | | | |

| **Table S7.** Genotype estimation for a two-allele site | | | | | |
| --- | --- | --- | --- | --- | --- |
| Group | R1’s Allele | R2’s Allele | Expected Genotype | Relative Read Count | |
|  |  |  |  | Wildtype | Mutant |
| I (AA\|AA) | A |  | AA\|AA | 1 | 0 |
|  | a |  | aa\|aa | 0 | 1 |
| II (AA\|AB) | A | a | AA\|Aa | (2-f)/2 | f/2 |
|  | a | A | aa\|Aa | f/2 | (2-f)/2 |
| III (AB\|AA) | A | a | Aa\|AA | (1+f)/2 | (1-f)/2 |
|  | a | A | Aa\|aa | (1-f)/2 | (1+f)/2 |
| IV (AB\|AB) | A | a | Aa\|Aa | 1/2 | 1/2 |
|  | a | A | Aa\|Aa | 1/2 | 1/2 |
| 1. Group: estimated genotype groups by allelic read counts. | | | | | |
| 2. A and a: wildtype and mutant alleles for a polymorphic site | | | | | |

| **Table S8.** Genotype estimation for a site with more than two alleles | | | | | | | |
| --- | --- | --- | --- | --- | --- | --- | --- |
| Group | R1’s Allele | R2’s Allele | R3’s Allele | Expected Genotype | Relative Read Count | | |
|  |  |  |  |  | Wildtype | Mutant 1 | Mutant 2 |
| I (AA\|AA) | A |  |  | AA\|AA | 1 | 0 | 0 |
|  | a |  |  | aa\|aa | 0 | 1 | 0 |
| II (AA\|AB) | A | a |  | AA\|Aa | (2-f)/2 | f/2 | 0 |
|  | a | A |  | aa\|Aa | f/2 | (2-f)/2 | 0 |
|  | a | b |  | aa\|ab | 0 | (2-f)/2 | f/2 |
| III (AB\|AA) | A | a |  | Aa\|AA | (1+f)/2 | (1-f)/2 | 0 |
|  | a | A |  | Aa\|aa | (1-f)/2 | (1+f)/2 | 0 |
|  | a | b |  | ab\|aa | 0 | (1+f)/2 | (1-f)/2 |
| IV (AB\|AB) | A | a |  | Aa\|Aa | 1/2 | 1/2 | 0 |
|  | a | A |  | Aa\|Aa | 1/2 | 1/2 | 0 |
|  | a | b |  | ab\|ab | 0 | 1/2 | 1/2 |
| V (AB\|AC) | A | a | b | Aa\|Ab | 1/2 | (1-f)/2 | f/2 |
|  | a | A | b | Aa\|ab | (1-f)/2 | 1/2 | f/2 |
|  | a | b | A | ab\|Aa | f/2 | 1/2 | (1-f)/2 |
|  | a | b | c | ab\|ac | 0 | 1/2 | (1-f)/2 |
| 1. Group: estimated genotype groups by allelic read counts. A-C: different alleles. | | | | | | | |
| 2. A and a-c: wildtype and mutant alleles for a polymorphic site. | | | | | | | |

| **Table S9.** Genotype estimation for fetal Wilson diseases using plasma samples | | | | | | | |
| --- | --- | --- | --- | --- | --- | --- | --- |
| SampleID | Position | Wildtype Reads | Mutant Reads | Fetal Fraction | ∆AIC | Estimated Genotype | Estimation Concordant |
| G2256 | p.P992L | 222 | 11 | 0.0944 | 17.13 | AA\|AB | Yes |
|  | p.E854G | 123 | 100 | 0.1032 | 2.38 | AB\|AA | Yes |
| G2671 | p.L795F | 193 | 10 | 9.86 | 16.27 | AA\|AB | Yes |
|  | p.G1335fs | 97 | 96 | 0.05* | 0.38 | AB\|AB | Yes |
| G2672 | p.A874P | 399 | 0 | 0.05* | 19.28 | AA\|AA | Yes |
|  | p.M769fs | 268 | 199 | 0.1478 | 10.23 | AB\|AA | Yes |
| G3203 | p.S1067N | 382 | 24 | 0.1182 | 46.42 | AA\|AB | Yes |
|  | p.R778L | 238 | 160 | 0.196 | 15.39 | AB\|AA | Yes |
| *As fetal fraction not listed in the original paper, 0.05 was assumed. | | | | | | | |

| **Table S10**. Genotype estimation for the hbb dataset using maternal plasma samples | | | | | | | |
| --- | --- | --- | --- | --- | --- | --- | --- |
| SampleID | Family | Mutant Reads | Wildtype Reads | Fetal Fraction | ∆AIC | Estimated Genotype | Discordant Genotype |
| ID001 | 13535 | 873 | 1093 | 0.0997 | 24.37423 | AB\|AA |  |
| ID002 | 13535 | 59 | 983 | 0.0997 | 108.7859 | AA\|AB |  |
| ID003 | 13551 | 474 | 471 | 0.0606 | 3.112722 | AB\|AB |  |
| ID004 | 13551 | 0 | 1464 | 0.0606 | 88.87129 | AA\|AA |  |
| ID005 | 13552 | 645 | 754 | 0.0702 | 8.417442 | AB\|AA |  |
| ID006 | 13552 | 0 | 925 | 0.0702 | 64.94543 | AA\|AA |  |
| ID007 | 13757 | 411 | 371 | 0.0715 | 1.721744 | AB\|AA | AB\|AB |
| ID008 | 14137 | 859 | 814 | 0.0771 | 3.021875 | AB\|AB |  |
| ID009 | 14137 | 43 | 1140 | 0.0771 | 49.32768 | AA\|AB |  |
| ID010 | 5001 | 764 | 850 | 0.0944 | 1.837886 | AB\|AA | AB\|AB |
| ID011 | 5001 | 0 | 738 | 0.0944 | 70.19435 | AA\|AA |  |
| ID012 | 5003 | 571 | 624 | 0.1283 | 6.159294 | AB\|AB |  |
| ID013 | 5003 | 68 | 910 | 0.1283 | 150.4004 | AA\|AB |  |
| ID014 | 5004 | 436 | 547 | 0.0694 | 10.68569 | AB\|AA |  |
| ID015 | 5004 | 41 | 1639 | 0.0694 | 19.1987 | AA\|AB |  |
| ID016 | 5005 | 760 | 809 | 0.093 | 4.488893 | AB\|AB |  |
| ID017 | 5005 | 43 | 1087 | 0.093 | 50.50314 | AA\|AB |  |
| ID018 | 5009 | 1538 | 1928 | 0.103 | 43.6587 | AB\|AA |  |
| ID019 | 5009 | 166 | 2491 | 0.103 | 330.802 | AA\|AB |  |
| ID020 | 5010 | 269 | 283 | 0.0865 | 1.717663 | AB\|AB |  |
| ID021 | 5010 | 0 | 513 | 0.0865 | 44.28228 | AA\|AA |  |
| ID022 | 5013 | 586 | 507 | 0.1566 | 2.190303 | AB\|AB |  |
| ID023 | 5013 | 44 | 564 | 0.1566 | 100.4654 | AA\|AB |  |
| ID024 | 5014 | 273 | 370 | 0.0764 | 11.08638 | AB\|AA |  |
| ID025 | 5014 | 0 | 1301 | 0.0764 | 100.1029 | AA\|AA |  |
| ID026 | 5015 | 286 | 547 | 0.1671 | 64.46166 | AB\|AA |  |
| ID027 | 5015 | 0 | 858 | 0.1671 | 148.4022 | AA\|AA |  |
| ID028 | 5018 | 784 | 773 | 0.1194 | 19.71751 | AB\|AB |  |
| ID029 | 5018 | 0 | 2332 | 0.1194 | 285.6509 | AA\|AA |  |
| ID030 | 5020 | 432 | 425 | 0.0781 | 4.147742 | AB\|AB |  |
| ID031 | 5028 | 343 | 406 | 0.081 | 5.298028 | AB\|AA |  |
| ID032 | 5028 | 0 | 861 | 0.081 | 70.02174 | AA\|AA |  |
| ID033 | 5048 | 451 | 414 | 0.0782 | 0.492717 | AB\|AA | AB\|AB |
| ID034 | 5048 | 33 | 985 | 0.0782 | 31.21931 | AA\|AB |  |
| ID035 | 5092 | 884 | 884 | 0.1094 | 21.28771 | AB\|AB |  |
| ID036 | 5092 | 0 | 1191 | 0.1094 | 132.6987 | AA\|AA |  |
| ID037 | 5101 | 914 | 751 | 0.077 | 15.25062 | AB\|AA |  |
| ID038 | 5114 | 581 | 588 | 0.0906 | 8.363284 | AB\|AB |  |
| ID039 | 5114 | 77 | 1597 | 0.0906 | 116.6841 | AA\|AB |  |
| ID040 | 5131 | 880 | 951 | 0.1527 | 21.34526 | AB\|AB |  |
| ID041 | 5131 | 0 | 2089 | 0.1527 | 330.3505 | AA\|AA |  |
| ID042 | 5137 | 292 | 293 | 0.1007 | 5.760384 | AB\|AB |  |
| ID043 | 5137 | 48 | 917 | 0.1007 | 78.8603 | AA\|AB |  |
| ID044 | 5151 | 423 | 427 | 0.1533 | 18.97808 | AB\|AB |  |
| ID045 | 5151 | 56 | 536 | 0.1533 | 153.3917 | AA\|AB |  |
| ID046 | 5161 | 497 | 516 | 0.0928 | 5.224999 | AB\|AB |  |
| ID047 | 5161 | 66 | 829 | 0.0928 | 140.4725 | AA\|AB |  |
| ID048 | 5165 | 364 | 533 | 0.0716 | 19.63193 | AB\|AA |  |
| ID049 | 5165 | 0 | 907 | 0.0716 | 64.97557 | AA\|AA |  |
| ID050 | 5175 | 376 | 390 | 0.1331 | 9.942753 | AB\|AB |  |
| ID051 | 5175 | 0 | 931 | 0.1331 | 126.9462 | AA\|AA |  |
| ID052 | 5177 | 846 | 868 | 0.1068 | 14.94546 | AB\|AB |  |
| ID053 | 5177 | 87 | 1355 | 0.1068 | 170.0045 | AA\|AB |  |
| ID054 | 5219 | 728 | 775 | 0.1066 | 7.118597 | AB\|AB |  |
| ID055 | 5219 | 0 | 1284 | 0.1066 | 139.3514 | AA\|AA |  |
| ID056 | 5236 | 712 | 699 | 0.0936 | 9.975439 | AB\|AB |  |
| ID057 | 5236 | 85 | 1538 | 0.0936 | 145.8412 | AA\|AB |  |
| ID058 | 5270 | 352 | 415 | 0.109 | 4.621488 | AB\|AA |  |
| ID059 | 5270 | 60 | 1066 | 0.109 | 105.4209 | AA\|AB |  |
| ID060 | 5295 | 268 | 396 | 0.2165 | 24.43894 | AB\|AA |  |
| ID061 | 5295 | 121 | 1026 | 0.2165 | 361.9365 | AA\|AB |  |
| ID062 | 5327 | 230 | 346 | 0.1336 | 20.80775 | AB\|AA |  |
| ID063 | 5327 | 56 | 912 | 0.1336 | 104.9294 | AA\|AB |  |
| ID064 | 5362 | 417 | 480 | 0.268 | 32.24373 | AB\|AB |  |
| ID065 | 5362 | 116 | 551 | 0.268 | 110.5311 | AA\|AB |  |
| ID066 | 5425 | 313 | 298 | 0.1902 | 16.73694 | AB\|AB |  |
| ID067 | 5425 | 136 | 1068 | 0.1902 | 420.6528 | AA\|AB |  |
| ID068 | 5487 | 1829 | 1943 | 0.1116 | 21.72237 | AB\|AB |  |
| ID069 | 5487 | 129 | 1726 | 0.1116 | 280.04 | AA\|AB |  |
| ID070 | 5536 | 320 | 339 | 0.1058 | 3.382702 | AB\|AB |  |
| ID071 | 5536 | 0 | 633 | 0.1058 | 67.64511 | AA\|AA |  |
| ID072 | 5002 | 711 | 880 | 0.077 | 16.61651 | AB\|AA |  |
| ID073 | 5002 | 0 | 2690 | 0.077 | 209.8341 | AA\|AA |  |
| ID074 | 5316 | 432 | 401 | 0.1034 | 2.520227 | AB\|AB |  |
| ID075 | 5316 | 0 | 947 | 0.1034 | 99.30308 | AA\|AA |  |
| ID076 | 5323 | 615 | 622 | 0.1078 | 12.94407 | AB\|AB |  |
| ID077 | 5323 | 0 | 1464 | 0.1078 | 160.8975 | AA\|AA |  |
| ID078 | 5021 | 1783 | 2020 | 0.0884 | 12.17576 | AB\|AA |  |
| ID079 | 5021 | 200 | 3931 | 0.0884 | 318.0574 | AA\|AB |  |
| ID080 | 5023 | 937 | 1107 | 0.0883 | 14.1011 | AB\|AA |  |
| ID081 | 5023 | 0 | 663 | 0.0883 | 58.73874 | AA\|AA |  |
| ID082 | 5029 | 1360 | 1658 | 0.1008 | 29.45986 | AB\|AA |  |
| ID083 | 5042 | 435 | 451 | 0.0827 | 3.42797 | AB\|AB |  |
| ID084 | 5042 | 0 | 1282 | 0.0827 | 107.0211 | AA\|AA |  |
| ID085 | 5045 | 871 | 791 | 0.1725 | 22.32686 | AB\|AB |  |
| ID086 | 5045 | 0 | 1405 | 0.1725 | 252.0372 | AA\|AA |  |
| ID087 | 5040 | 796 | 976 | 0.1398 | 15.68483 | AB\|AA |  |
| ID088 | 5040 | 135 | 1798 | 0.1398 | 300.5732 | AA\|AB |  |
| ID089 | 5057 | 1413 | 1574 | 0.0838 | 5.997005 | AB\|AA |  |
| ID090 | 5057 | 147 | 3184 | 0.0838 | 212.6443 | AA\|AB |  |
| ID091 | 5066 | 255 | 261 | 0.1896 | 16.5878 | AB\|AB |  |
| ID092 | 5068 | 496 | 782 | 0.1933 | 63.30911 | AB\|AA |  |
| ID093 | 5488 | 749 | 723 | 0.1088 | 11.84858 | AB\|AB |  |
| ID094 | 5488 | 36 | 884 | 0.1088 | 40.82689 | AA\|AB |  |
| ID095 | 5548 | 776 | 720 | 0.0903 | 2.1073 | AB\|AB |  |
| ID096 | 5548 | 0 | 1172 | 0.0903 | 107.0409 | AA\|AA |  |
| ID097 | 5605 | 364 | 342 | 0.1791 | 15.05108 | AB\|AB |  |
| ID098 | 5605 | 59 | 543 | 0.1791 | 167.7112 | AA\|AB |  |
| ID099 | 5610 | 772 | 701 | 0.1267 | 5.749146 | AB\|AB |  |
| ID100 | 5646 | 311 | 373 | 0.127 | 4.711322 | AB\|AA |  |
| ID101 | 5646 | 76 | 1009 | 0.127 | 168.8543 | AA\|AB |  |
| ID102 | 5049 | 1804 | 2059 | 0.0919 | 14.23768 | AB\|AA |  |
| ID103 | 5055 | 1195 | 1228 | 0.1026 | 18.84603 | AB\|AB |  |
| ID104 | 5055 | 9 | 2704 | 0.1026 | 283.3526 | AA\|AA |  |
| ID105 | 5062 | 1050 | 1288 | 0.0816 | 23.30838 | AB\|AA |  |
| ID106 | 5062 | 0 | 955 | 0.0816 | 78.3693 | AA\|AA |  |
| ID107 | 5063 | 324 | 333 | 0.0732 | 2.209868 | AB\|AB |  |
| ID108 | 5063 | 0 | 2358 | 0.0732 | 174.4914 | AA\|AA |  |
| ID109 | 5064 | 539 | 695 | 0.08 | 17.09047 | AB\|AA |  |
| ID110 | 5064 | 0 | 2420 | 0.08 | 196.2034 | AA\|AA |  |
| ID111 | 5357 | 1246 | 1498 | 0.1013 | 22.92737 | AB\|AA |  |
| ID112 | 5357 | 0 | 2017 | 0.1013 | 208.2924 | AA\|AA |  |
| ID113 | 5374 | 1246 | 1206 | 0.0886 | 12.21741 | AB\|AB |  |
| ID114 | 5374 | 73 | 1802 | 0.0886 | 90.2261 | AA\|AB |  |
| ID115 | 5478 | 781 | 802 | 0.0821 | 7.250213 | AB\|AB |  |
| ID116 | 5072 | 502 | 749 | 0.1076 | 38.7927 | AB\|AA |  |
| ID117 | 5072 | 0 | 4033 | 0.1076 | 444.5239 | AA\|AA |  |
| ID118 | 5074 | 631 | 1014 | 0.0644 | 42.56217 | AB\|AA |  |
| ID119 | 5074 | 0 | 4703 | 0.0644 | 306.3922 | AA\|AA |  |
| ID120 | 5082 | 1065 | 1121 | 0.1163 | 16.68413 | AB\|AB |  |
| ID121 | 5082 | 0 | 2944 | 0.1163 | 351.2566 | AA\|AA |  |
| ID122 | 5083 | 1717 | 2109 | 0.1247 | 38.31426 | AB\|AA |  |
| ID123 | 5083 | 149 | 2498 | 0.1247 | 273.9695 | AA\|AB |  |
| ID124 | 5086 | 1137 | 1563 | 0.0977 | 57.61071 | AB\|AA |  |
| ID125 | 5086 | 33 | 845 | 0.0977 | 37.03113 | AA\|AB |  |
| ID126 | 5089 | 1372 | 1507 | 0.1012 | 2.219273 | AB\|AB |  |
| ID127 | 5089 | 43 | 1075 | 0.1012 | 49.40683 | AA\|AB |  |
| ID128 | 5519 | 730 | 858 | 0.196 | 11.37354 | AB\|AB |  |
| ID129 | 5519 | 113 | 1067 | 0.196 | 317.1635 | AA\|AB |  |
| ID130 | 5551 | 519 | 707 | 0.1213 | 27.66148 | AB\|AA |  |
| ID131 | 5551 | 88 | 990 | 0.1213 | 213.2628 | AA\|AB |  |
| ID132 | 5638 | 570 | 856 | 0.1708 | 56.44557 | AB\|AA |  |
| ID133 | 5638 | 89 | 1102 | 0.1708 | 207.1707 | AA\|AB |  |
| ID134 | 5144 | 985 | 947 | 0.1128 | 16.13075 | AB\|AB |  |
| ID135 | 5144 | 126 | 2487 | 0.1128 | 197.1659 | AA\|AB |  |
| ID136 | 5467 | 1242 | 1527 | 0.0835 | 28.3323 | AB\|AA |  |
| ID137 | 5467 | 0 | 2951 | 0.0835 | 250.2768 | AA\|AA |  |
| ID138 | 5474 | 1155 | 1202 | 0.0956 | 12.62659 | AB\|AB |  |
| ID139 | 5474 | 77 | 1614 | 0.0956 | 115.2583 | AA\|AB |  |
| ID140 | 5483 | 911 | 911 | 0.0816 | 12.17247 | AB\|AB |  |
| ID141 | 5483 | 64 | 1834 | 0.0816 | 64.05203 | AA\|AB |  |
| ID142 | 5061 | 599 | 696 | 0.1358 | 2.404085 | AB\|AA |  |
| ID143 | 5061 | 153 | 1911 | 0.1358 | 355.7958 | AA\|AB |  |
| ID144 | 5556 | 608 | 676 | 0.0761 | 2.91212 | AB\|AA |  |
| ID145 | 5556 | 0 | 1425 | 0.0761 | 109.3004 | AA\|AA |  |
| ID146 | 5400 | 509 | 496 | 0.0964 | 6.868883 | AB\|AB |  |
| ID147 | 5400 | 105 | 2181 | 0.0964 | 158.6378 | AA\|AB |  |
| ID148 | 5486 | 1140 | 1148 | 0.0989 | 20.90199 | AB\|AB |  |
| ID149 | 5486 | 17 | 8371 | 0.0989 | 847.3967 | AA\|AA |  |
| ID150 | 5524 | 214 | 315 | 0.1488 | 18.438 | AB\|AA |  |
| ID151 | 5524 | 0 | 1167 | 0.1488 | 179.0955 | AA\|AA |  |
| ID152 | 5623 | 348 | 355 | 0.1582 | 15.5845 | AB\|AB |  |
| ID153 | 5623 | 0 | 589 | 0.1582 | 95.84291 | AA\|AA |  |
| ID154 | 5054 | 619 | 637 | 0.15 | 23.14172 | AB\|AB |  |
| ID155 | 5058 | 488 | 763 | 0.1696 | 57.6785 | AB\|AA |  |
| ID156 | 5058 | 0 | 1265 | 0.1696 | 222.7944 | AA\|AA |  |
| ID157 | 5109 | 789 | 790 | 0.0772 | 9.284034 | AB\|AB |  |
| ID158 | 5432 | 1703 | 1920 | 0.1117 | 3.192799 | AB\|AA |  |
| ID159 | 5432 | 99 | 2330 | 0.1117 | 119.6 | AA\|AB |  |
| ID160 | 1294 | 795 | 865 | 0.1018 | 2.991198 | AB\|AB |  |
| ID161 | 1294 | 0 | 875 | 0.1018 | 90.20216 | AA\|AA |  |
| ID162 | 2406 | 768 | 781 | 0.0679 | 5.389918 | AB\|AB |  |
| ID163 | 2406 | 0 | 1385 | 0.0679 | 94.4443 | AA\|AA |  |
| ID164 | 2466 | 1837 | 1861 | 0.1568 | 84.46726 | AB\|AB |  |
| ID165 | 2466 | 0 | 3057 | 0.1568 | 497.615 | AA\|AA |  |
| ID166 | 2543 | 985 | 1108 | 0.1194 | 0.540112 | AB\|AB |  |
| ID167 | 2543 | 140 | 1624 | 0.1194 | 332.9977 | AA\|AB |  |
| ID168 | 2691 | 1189 | 1211 | 0.0668 | 7.789758 | AB\|AB |  |
| ID169 | 2691 | 22 | 994 | 0.0668 | 5.50939 | AA\|AB |  |
| ID170 | 671 | 288 | 424 | 0.1566 | 25.27017 | AB\|AA |  |
| ID171 | 671 | 35 | 566 | 0.1566 | 63.13618 | AA\|AB |  |
| ID172 | 2851 | 627 | 452 | 0.0925 | 23.1959 | AB\|AA |  |
| ID173 | 2962 | 510 | 469 | 0.1433 | 8.480902 | AB\|AB |  |
| ID174 | 2962 | 0 | 991 | 0.1433 | 146.0416 | AA\|AA |  |
| ID175 | 3005 | 1425 | 1429 | 0.0746 | 15.32942 | AB\|AB |  |
| ID176 | 3006 | 912 | 727 | 0.09 | 20.06039 | AB\|AA |  |
| ID177 | 3044 | 1407 | 1358 | 0.0755 | 8.393177 | AB\|AB |  |
| ID178 | 3044 | 0 | 947 | 0.0755 | 71.70717 | AA\|AA |  |
| ID179 | 3066 | 1372 | 1402 | 0.0897 | 17.01363 | AB\|AB |  |
| ID180 | 3066 | 118 | 2951 | 0.0897 | 142.667 | AA\|AB |  |
| ID181 | 5051 | 902 | 805 | 0.091 | 3.508454 | AB\|AA | AB\|AB |
| ID182 | 5123 | 513 | 590 | 0.118 | 2.790955 | AB\|AA | AB\|AB |
| ID183 | 5123 | 0 | 1929 | 0.118 | 233.2058 | AA\|AA |  |
| ID184 | 5195 | 883 | 726 | 0.16 | 8.948611 | AB\|AA | AB\|AB |
| ID185 | 5195 | 0 | 1040 | 0.16 | 172.089 | AA\|AA |  |
| ID186 | 5450 | 1135 | 1119 | 0.068 | 8.267304 | AB\|AB | AB\|AA |
| ID187 | 5450 | 52 | 1612 | 0.068 | 48.15211 | AA\|AB |  |
| ID188 | 5084 | 2836 | 2493 | 0.05 | 20.98945 | AB\|AA | AB\|AB |

| **Table S11**. Genotype estimation for the arnshl dataset using maternal plasma samples | | | | | | | |
| --- | --- | --- | --- | --- | --- | --- | --- |
| SampleID | Sample Number | Mutant Reads | Wildtype Reads | Fetal Fraction | ∆AIC | Estimated Genotype | Discordant Genotype |
| Id001 | K002 | 883 | 855 | 0.088569 | 8.714489 | AB\|AB |  |
| Id002 | K004 | 422 | 432 | 0.139957 | 14.07655 | AB\|AB |  |
| Id003 | K006 | 620 | 586 | 0.153628 | 18.27459 | AB\|AB |  |
| Id004 | K010 | 509 | 493 | 0.101678 | 7.14801 | AB\|AB |  |
| Id005 | K012 | 642 | 653 | 0.114721 | 14.62164 | AB\|AB |  |
| Id006 | K013 | 670 | 842 | 0.088232 | 18.61411 | AB\|AA |  |
| Id007 | K015 | 914 | 1059 | 0.080111 | 10.579 | AB\|AA |  |
| Id008 | K019 | 953 | 746 | 0.110906 | 25.0772 | AB\|AA |  |
| Id009 | K037 | 895 | 1112 | 0.05298 | 17.37351 | AB\|AA |  |
| Id010 | K041 | 557 | 464 | 0.118662 | 7.697089 | AB\|AA |  |
| Id011 | K055 | 552 | 531 | 0.13483 | 14.17161 | AB\|AB |  |
| Id012 | K056 | 178 | 186 | 0.114432 | 2.958994 | AB\|AB |  |
| Id013 | K060 | 586 | 553 | 0.102044 | 5.164084 | AB\|AB |  |
| Id014 | K064 | 514 | 504 | 0.077751 | 4.614538 | AB\|AB |  |
| Id015 | K065 | 1409 | 1342 | 0.138335 | 34.49852 | AB\|AB |  |
| Id016 | K067 | 300 | 304 | 0.089518 | 4.141548 | AB\|AB |  |
| Id017 | K069 | 945 | 730 | 0.126744 | 27.66886 | AB\|AA |  |
| Id018 | K071 | 532 | 556 | 0.11 | 7.943643 | AB\|AB |  |
| Id019 | K073 | 1221 | 1171 | 0.081621 | 7.80836 | AB\|AB |  |
| Id020 | K079 | 1273 | 979 | 0.1269 | 38.46115 | AB\|AA |  |
| Id021 | K081 | 402 | 288 | 0.10386 | 16.2824 | AB\|AA |  |
| Id022 | K092 | 330 | 337 | 0.081133 | 3.266681 | AB\|AB |  |
| Id023 | K096 | 1025 | 988 | 0.060782 | 2.947215 | AB\|AB |  |
| Id024 | K100 | 550 | 556 | 0.1 | 9.911647 | AB\|AB |  |
| Id025 | K101 | 649 | 653 | 0.072629 | 6.304113 | AB\|AB |  |
| Id026 | K003 | 607 | 730 | 0.085689 | 11.27803 | AB\|AA |  |
| Id027 | K003 | 0 | 1166 | 0.085689 | 100.8737 | AA\|AA |  |
| Id028 | K005 | 580 | 489 | 0.082089 | 7.745945 | AB\|AA | AB\|AB |
| Id029 | K005 | 0 | 992 | 0.082089 | 81.94882 | AA\|AA |  |
| Id030 | K008 | 705 | 790 | 0.06562 | 4.720119 | AB\|AA |  |
| Id031 | K008 | 53 | 1639 | 0.06562 | 49.53363 | AA\|AB |  |
| Id032 | K009 | 573 | 538 | 0.126946 | 9.11539 | AB\|AB |  |
| Id033 | K009 | 99 | 1179 | 0.126946 | 234.978 | AA\|AB |  |
| Id034 | K011 | 443 | 533 | 0.154789 | 4.418441 | AB\|AA |  |
| Id035 | K011 | 0 | 958 | 0.154789 | 153.0193 | AA\|AA |  |
| Id036 | K018 | 497 | 593 | 0.1388 | 5.618498 | AB\|AA |  |
| Id037 | K018 | 0 | 860 | 0.1388 | 122.4339 | AA\|AA |  |
| Id038 | K020 | 482 | 513 | 0.061629 | 0.03953 | AB\|AA |  |
| Id039 | K020 | 0 | 1289 | 0.061629 | 79.49212 | AA\|AA |  |
| Id040 | K025 | 358 | 351 | 0.106044 | 6.527907 | AB\|AB |  |
| Id041 | K025 | 0 | 347 | 0.106044 | 36.76579 | AA\|AA |  |
| Id042 | K027 | 644 | 673 | 0.080927 | 3.949571 | AB\|AB |  |
| Id043 | K027 | 0 | 1910 | 0.080927 | 156.4546 | AA\|AA |  |
| Id044 | K029 | 1529 | 1519 | 0.063479 | 11.03569 | AB\|AB |  |
| Id045 | K029 | 60 | 2583 | 0.063479 | 23.8925 | AA\|AB |  |
| Id046 | K030 | 557 | 642 | 0.073692 | 6.021466 | AB\|AA |  |
| Id047 | K030 | 8 | 377 | 0.073692 | 0.137941 | AA\|AB |  |
| Id048 | K038 | 267 | 344 | 0.107184 | 9.509941 | AB\|AA |  |
| Id049 | K038 | 22 | 439 | 0.107184 | 34.33046 | AA\|AB |  |
| Id050 | K039 | 521 | 608 | 0.09 | 6.520351 | AB\|AA |  |
| Id051 | K039 | 0 | 1222 | 0.09 | 111.2694 | AA\|AA |  |
| Id052 | K040 | 161 | 194 | 0.123998 | 2.7255 | AB\|AA |  |
| Id053 | K040 | 0 | 330 | 0.123998 | 41.17863 | AA\|AA |  |
| Id054 | K042 | 690 | 715 | 0.054687 | 1.471063 | AB\|AB |  |
| Id055 | K042 | 0 | 1671 | 0.054687 | 91.42902 | AA\|AA |  |
| Id056 | K043 | 1612 | 1500 | 0.056139 | 2.765095 | AB\|AA | AB\|AB |
| Id057 | K043 | 79 | 3103 | 0.056139 | 48.7529 | AA\|AB |  |
| Id058 | K044 | 1186 | 1196 | 0.068676 | 9.885216 | AB\|AB |  |
| Id059 | K044 | 82 | 2431 | 0.068676 | 81.30116 | AA\|AB |  |
| Id060 | K045 | 219 | 266 | 0.110891 | 4.465874 | AB\|AA |  |
| Id061 | K045 | 0 | 335 | 0.110891 | 37.17313 | AA\|AA |  |
| Id062 | K046 | 398 | 372 | 0.085592 | 1.200082 | AB\|AB |  |
| Id063 | K046 | 0 | 709 | 0.085592 | 60.87856 | AA\|AA |  |
| Id064 | K047 | 614 | 525 | 0.056671 | 6.43437 | AB\|AA | AB\|AB |
| Id065 | K047 | 30 | 1295 | 0.056671 | 14.07334 | AA\|AB |  |
| Id066 | K048 | 169 | 229 | 0.091958 | 7.686317 | AB\|AA |  |
| Id067 | K048 | 1 | 1070 | 0.091958 | 99.48927 | AA\|AA |  |
| Id068 | K049 | 1060 | 1017 | 0.12 | 19.75626 | AB\|AB |  |
| Id069 | K049 | 104 | 1410 | 0.12 | 226.5393 | AA\|AB |  |
| Id070 | K049 | 0 | 909 | 0.12 | 111.2292 | AA\|AA |  |
| Id071 | K050 | 935 | 1064 | 0.054914 | 8.144875 | AB\|AA | AB\|AB |
| Id072 | K050 | 41 | 1975 | 0.054914 | 12.55012 | AA\|AB |  |
| Id073 | K051 | 692 | 694 | 0.088906 | 10.64236 | AB\|AB |  |
| Id074 | K051 | 0 | 1361 | 0.088906 | 122.4923 | AA\|AA |  |
| Id075 | K052 | 1161 | 1080 | 0.058712 | 1.783962 | AB\|AA | AB\|AB |
| Id076 | K052 | 0 | 1663 | 0.058712 | 97.86239 | AA\|AA |  |
| Id077 | K053 | 562 | 583 | 0.134787 | 15.29738 | AB\|AB |  |
| Id078 | K053 | 71 | 874 | 0.134787 | 166.5375 | AA\|AB |  |
| Id079 | K054 | 361 | 345 | 0.106443 | 4.625591 | AB\|AB |  |
| Id080 | K054 | 0 | 1196 | 0.106443 | 129.5275 | AA\|AA |  |
| Id081 | K058 | 312 | 455 | 0.17679 | 26.74383 | AB\|AA |  |
| Id082 | K058 | 70 | 753 | 0.17679 | 180.85 | AA\|AB |  |
| Id083 | K059 | 454 | 519 | 0.059428 | 4.292346 | AB\|AA |  |
| Id084 | K059 | 0 | 1228 | 0.059428 | 72.90435 | AA\|AA |  |
| Id085 | K061 | 289 | 280 | 0.100228 | 3.934714 | AB\|AB |  |
| Id086 | K061 | 0 | 745 | 0.100228 | 75.42144 | AA\|AA |  |
| Id087 | K062 | 876 | 874 | 0.108196 | 20.1725 | AB\|AB |  |
| Id088 | K062 | 0 | 1545 | 0.108196 | 170.5086 | AA\|AA |  |
| Id089 | K063 | 577 | 691 | 0.056086 | 8.806072 | AB\|AA |  |
| Id090 | K063 | 36 | 1070 | 0.056086 | 34.88123 | AA\|AB |  |
| Id091 | K066 | 540 | 505 | 0.1 | 3.479127 | AB\|AB |  |
| Id092 | K066 | 0 | 1109 | 0.1 | 112.5049 | AA\|AA |  |
| Id093 | K068 | 478 | 536 | 0.072667 | 3.075636 | AB\|AA |  |
| Id094 | K068 | 26 | 854 | 0.072667 | 21.04089 | AA\|AB |  |
| Id095 | K070 | 201 | 245 | 0.093695 | 4.336803 | AB\|AA |  |
| Id096 | K070 | 50 | 1110 | 0.093695 | 70.22661 | AA\|AB |  |
| Id097 | K072 | 353 | 473 | 0.10433 | 16.09054 | AB\|AA |  |
| Id098 | K072 | 0 | 578 | 0.10433 | 60.79058 | AA\|AA |  |
| Id099 | K074 | 442 | 501 | 0.05 | 3.544472 | AB\|AA | AB\|AB |
| Id100 | K074 | 36 | 1412 | 0.05 | 22.85759 | AA\|AB |  |
| Id101 | K075 | 781 | 953 | 0.120133 | 16.31895 | AB\|AA |  |
| Id102 | K075 | 33 | 654 | 0.120133 | 50.44945 | AA\|AB |  |
| Id103 | K077 | 211 | 262 | 0.09 | 5.358006 | AB\|AA |  |
| Id104 | K077 | 0 | 460 | 0.09 | 41.29388 | AA\|AA |  |
| Id105 | K078 | 847 | 888 | 0.055222 | 0.766105 | AB\|AB | AB\|AA |
| Id106 | K078 | 1 | 1630 | 0.055222 | 90.05612 | AA\|AA |  |
| Id107 | K078 | 730 | 740 | 0.055222 | 3.384014 | AB\|AB |  |
| Id108 | K080 | 891 | 1051 | 0.100708 | 12.53968 | AB\|AA |  |
| Id109 | K080 | 0 | 972 | 0.100708 | 99.19967 | AA\|AA |  |
| Id110 | K082 | 879 | 1343 | 0.124789 | 81.53692 | AB\|AA |  |
| Id111 | K082 | 0 | 2254 | 0.124789 | 288.9815 | AA\|AA |  |
| Id112 | K085 | 592 | 716 | 0.143862 | 8.572184 | AB\|AA |  |
| Id113 | K085 | 0 | 1296 | 0.143862 | 192.1236 | AA\|AA |  |
| Id114 | K086 | 833 | 949 | 0.077148 | 7.296172 | AB\|AA |  |
| Id115 | K086 | 25 | 850 | 0.077148 | 17.71129 | AA\|AB |  |
| Id116 | K087 | 998 | 1081 | 0.056109 | 2.768442 | AB\|AA |  |
| Id117 | K087 | 20 | 974 | 0.056109 | 5.409334 | AA\|AB |  |
| Id118 | K088 | 680 | 785 | 0.117379 | 4.438942 | AB\|AA |  |
| Id119 | K088 | 82 | 1342 | 0.117379 | 154.8676 | AA\|AB |  |
| Id120 | K089 | 558 | 547 | 0.129051 | 15.70284 | AB\|AB |  |
| Id121 | K089 | 0 | 897 | 0.129051 | 118.3904 | AA\|AA |  |
| Id122 | K090 | 716 | 692 | 0.070949 | 3.694151 | AB\|AB |  |
| Id123 | K090 | 0 | 799 | 0.070949 | 56.58878 | AA\|AA |  |
| Id124 | K091 | 1293 | 1252 | 0.076923 | 8.783704 | AB\|AB |  |
| Id125 | K091 | 79 | 2354 | 0.076923 | 75.50353 | AA\|AB |  |
| Id126 | K093 | 532 | 568 | 0.129871 | 9.307459 | AB\|AB |  |
| Id127 | K093 | 54 | 745 | 0.129871 | 116.9841 | AA\|AB |  |
| Id128 | K094 | 287 | 409 | 0.14714 | 20.93065 | AB\|AA |  |
| Id129 | K094 | 0 | 724 | 0.14714 | 109.3962 | AA\|AA |  |
| Id130 | K095 | 598 | 503 | 0.097471 | 8.068312 | AB\|AA | AB\|AB |
| Id131 | K095 | 0 | 877 | 0.097471 | 86.42381 | AA\|AA |  |
| Id132 | K097 | 808 | 895 | 0.052389 | 4.443549 | AB\|AA | AB\|AB |
| Id133 | K097 | 39 | 1869 | 0.052389 | 13.45921 | AA\|AB |  |
| Id134 | K099 | 624 | 673 | 0.0809 | 0.570911 | AB\|AB |  |
| Id135 | K099 | 41 | 1129 | 0.0809 | 44.05246 | AA\|AB |  |
| Id136 | K102 | 1062 | 1226 | 0.102238 | 9.610192 | AB\|AA |  |
| Id137 | K102 | 0 | 715 | 0.102238 | 73.85461 | AA\|AA |  |

| **Table S12**. Genotype estimation for the cfbest dataset using maternal plasma samples | | | | | | | |
| --- | --- | --- | --- | --- | --- | --- | --- |
| SampleID | Sample Number | Mutant Reads | Wildtype Reads | Fetal Fraction | ∆AIC | Estimated Genotype | Discordant Genotype |
| ID001 | FS213 | 765 | 1184 | 0.130402 | 76.47537 | AB\|AA |  |
| ID002 | FS213 | 149 | 1773 | 0.130402 | 355.2708 | AA\|AB |  |
| ID003 | FS227 | 1169 | 1367 | 0.055147 | 14.13616 | AB\|AA |  |
| ID004 | FS260 | 1285 | 1191 | 0.118115 | 12.47671 | AB\|AB |  |
| ID005 | FS295 | 72 | 1742 | 0.102643 | 86.97527 | AA\|AB |  |
| ID006 | GX005 | 517 | 667 | 0.106066 | 18.54447 | AB\|AA |  |
| ID007 | GX017 | 501 | 598 | 0.142019 | 5.346442 | AB\|AA |  |
| ID008 | GX033 | 642 | 771 | 0.106703 | 11.45451 | AB\|AA |  |
| ID009 | GX035 | 647 | 596 | 0.131164 | 8.11429 | AB\|AB |  |
| ID010 | GX045 | 854 | 988 | 0.088386 | 9.303052 | AB\|AA |  |
| ID011 | GX045 | 83 | 1620 | 0.088386 | 132.7904 | AA\|AB |  |
| ID012 | GX057 | 1091 | 1093 | 0.062127 | 8.197146 | AB\|AB | AB\|AA |
| ID013 | GX069 | 787 | 1034 | 0.091611 | 30.03573 | AB\|AA |  |
| ID014 | GX093 | 489 | 631 | 0.126338 | 18.05189 | AB\|AA |  |
| ID015 | GX100 | 1343 | 1301 | 0.096496 | 16.60388 | AB\|AB |  |
| ID016 | GX100 | 141 | 2353 | 0.096496 | 258.3844 | AA\|AB |  |
| ID017 | GX108 | 155 | 3268 | 0.078713 | 227.9796 | AA\|AB |  |
| ID018 | GX108 | 1847 | 1790 | 0.078713 | 13.61203 | AB\|AB |  |
| ID019 | GX222 | 1703 | 2136 | 0.0926 | 47.36152 | AB\|AA |  |
| ID020 | GX222 | 155 | 3319 | 0.0926 | 227.1249 | AA\|AB |  |
| ID021 | GX341 | 669 | 997 | 0.154505 | 61.92076 | AB\|AA |  |
| ID022 | GX358 | 473 | 587 | 0.084908 | 11.73615 | AB\|AA |  |
| ID023 | GX358 | 48 | 953 | 0.084908 | 75.27105 | AA\|AB |  |
| ID024 | GX361 | 625 | 627 | 0.054714 | 3.53459 | AB\|AB |  |
| ID025 | GX372 | 790 | 870 | 0.053451 | 3.810892 | AB\|AA |  |
| ID026 | GX373 | 896 | 865 | 0.081243 | 6.613627 | AB\|AB |  |
| ID027 | GX387 | 47 | 1001 | 0.087691 | 69.29973 | AA\|AB |  |
| ID028 | GX387 | 502 | 648 | 0.087691 | 16.79433 | AB\|AA |  |
| ID029 | GX397 | 892 | 1139 | 0.086537 | 27.58972 | AB\|AA |  |
| ID030 | GX400 | 50 | 1181 | 0.075906 | 65.72484 | AA\|AB |  |
| ID031 | GX400 | 537 | 641 | 0.075906 | 9.011955 | AB\|AA |  |
| ID032 | GX407 | 173 | 1790 | 0.140905 | 449.9492 | AA\|AB |  |
| ID033 | GX407 | 906 | 920 | 0.140905 | 32.64681 | AB\|AB |  |
| ID034 | GX413 | 605 | 722 | 0.08578 | 10.32153 | AB\|AA |  |
| ID035 | GX425 | 64 | 1028 | 0.109422 | 122.5169 | AA\|AB |  |
| ID036 | GX425 | 595 | 571 | 0.109422 | 8.771529 | AB\|AB |  |
| ID037 | GX426 | 451 | 558 | 0.064776 | 9.638896 | AB\|AA |  |
| ID038 | GX427 | 715 | 725 | 0.080261 | 7.697619 | AB\|AB |  |
| ID039 | GX430 | 647 | 576 | 0.054093 | 4.104905 | AB\|AA |  |
| ID040 | GX435 | 218 | 3292 | 0.104047 | 433.4297 | AA\|AB |  |
| ID041 | GX435 | 1571 | 2012 | 0.104047 | 53.10253 | AB\|AA |  |
| ID042 | GX439 | 615 | 605 | 0.066852 | 4.125618 | AB\|AB |  |
| ID043 | GX441 | 1361 | 1621 | 0.066763 | 21.44708 | AB\|AA |  |
| ID044 | GX441 | 107 | 2634 | 0.066763 | 132.0484 | AA\|AB |  |
| ID045 | GX445 | 1914 | 1874 | 0.054577 | 6.929566 | AB\|AB |  |
| ID046 | GX449 | 1200 | 1425 | 0.068751 | 18.54981 | AB\|AA |  |
| ID047 | GX456 | 620 | 864 | 0.136849 | 39.14862 | AB\|AA |  |
| ID048 | GX456 | 110 | 1243 | 0.136849 | 271.8745 | AA\|AB |  |
| ID049 | GX458 | 1502 | 1620 | 0.054821 | 3.553919 | AB\|AA |  |
| ID050 | GX458 | 79 | 2688 | 0.054821 | 63.93239 | AA\|AB |  |
| ID051 | GX459 | 953 | 1257 | 0.108775 | 40.0932 | AB\|AA |  |
| ID052 | GX462 | 711 | 831 | 0.07924 | 9.344844 | AB\|AA |  |
| ID053 | GX463 | 136 | 3156 | 0.064392 | 174.9113 | AA\|AB |  |
| ID054 | GX463 | 1469 | 1731 | 0.064392 | 20.49235 | AB\|AA |  |
| ID055 | GX465 | 655 | 656 | 0.103177 | 13.82406 | AB\|AB |  |
| ID056 | GX466 | 1297 | 1351 | 0.102337 | 16.78715 | AB\|AB |  |
| ID057 | GX468 | 1021 | 1300 | 0.089373 | 31.39007 | AB\|AA |  |
| ID058 | GX468 | 118 | 2469 | 0.089373 | 177.1967 | AA\|AB |  |
| ID059 | GX469 | 1173 | 1369 | 0.062855 | 14.609 | AB\|AA |  |
| ID060 | GX475 | 1549 | 1456 | 0.098172 | 10.78303 | AB\|AB |  |
| ID061 | GX476 | 79 | 2718 | 0.063891 | 61.65195 | AA\|AB |  |
| ID062 | GX476 | 1249 | 1226 | 0.063891 | 7.180905 | AB\|AB |  |
| ID063 | GX482 | 806 | 822 | 0.085216 | 9.131662 | AB\|AB |  |
| ID064 | GX483 | 96 | 2216 | 0.078655 | 129.6367 | AA\|AB |  |
| ID065 | GX493 | 1345 | 1223 | 0.151526 | 22.3898 | AB\|AB |  |
| ID066 | GX495 | 1987 | 2043 | 0.051921 | 5.058409 | AB\|AB |  |
| ID067 | GX509 | 1257 | 1273 | 0.064979 | 8.622615 | AB\|AB |  |
| ID068 | GX510 | 815 | 798 | 0.078997 | 7.405908 | AB\|AB |  |
| ID069 | GX532 | 853 | 834 | 0.145189 | 30.38551 | AB\|AB |  |
| ID070 | GX534 | 1188 | 1493 | 0.073527 | 30.39923 | AB\|AA |  |
| ID071 | GX536 | 34 | 990 | 0.064569 | 34.61789 | AA\|AB |  |
| ID072 | GX536 | 1301 | 1258 | 0.064569 | 5.130622 | AB\|AB |  |
| ID073 | GX547 | 1821 | 1686 | 0.093004 | 5.282721 | AB\|AB |  |
| ID074 | GX548 | 1322 | 1298 | 0.071713 | 10.06069 | AB\|AB |  |
| ID075 | GX549 | 1291 | 1278 | 0.079583 | 14.24886 | AB\|AB |  |
| ID076 | GX550 | 755 | 792 | 0.097066 | 7.439068 | AB\|AB |  |
| ID077 | GX551 | 1145 | 1344 | 0.059097 | 14.84009 | AB\|AA |  |
| ID078 | GX554 | 457 | 618 | 0.109834 | 22.46264 | AB\|AA |  |
| ID079 | GX559 | 907 | 734 | 0.118735 | 17.97758 | AB\|AA |  |
| ID080 | GX563 | 2285 | 1988 | 0.059372 | 20.21938 | AB\|AA |  |
| ID081 | GX567 | 1045 | 1156 | 0.077856 | 3.937056 | AB\|AA |  |
| ID082 | NF403 | 552 | 706 | 0.1382 | 18.58046 | AB\|AA |  |
| ID083 | NF409 | 1292 | 1213 | 0.083327 | 4.25764 | AB\|AB |  |
| ID084 | NF409 | 91 | 2351 | 0.083327 | 106.8782 | AA\|AB |  |
| ID085 | NF410 | 91 | 2328 | 0.064338 | 107.2055 | AA\|AB |  |
| ID086 | NF410 | 1105 | 1071 | 0.064338 | 4.644939 | AB\|AB |  |
| ID087 | NF413 | 44 | 1052 | 0.064993 | 55.34988 | AA\|AB |  |
| ID088 | NF413 | 965 | 1080 | 0.064993 | 6.312924 | AB\|AA |  |
| ID089 | NF415 | 680 | 855 | 0.110447 | 19.97489 | AB\|AA |  |
| ID090 | NF415 | 26 | 996 | 0.110447 | 4.281237 | AA\|AA | AA\|AB |
| ID091 | NF418 | 217 | 3011 | 0.097744 | 447.3741 | AA\|AB |  |
| ID092 | NF418 | 1319 | 1786 | 0.097744 | 61.77791 | AB\|AA |  |
| ID093 | NF421 | 799 | 772 | 0.071844 | 4.243565 | AB\|AB |  |
| ID094 | NF423 | 1085 | 1037 | 0.100136 | 11.7399 | AB\|AB |  |
| ID095 | NF426 | 44 | 1263 | 0.07152 | 45.5363 | AA\|AB |  |
| ID096 | NF426 | 747 | 909 | 0.07152 | 14.71972 | AB\|AA |  |
| ID097 | NF429 | 163 | 1711 | 0.151343 | 424.8869 | AA\|AB |  |
| ID098 | NF429 | 591 | 581 | 0.151343 | 24.10622 | AB\|AB |  |
| ID099 | NF432 | 1336 | 1543 | 0.050431 | 13.56468 | AB\|AA |  |
| ID100 | NF432 | 111 | 2951 | 0.050431 | 113.9083 | AA\|AB |  |
| ID101 | NF433 | 565 | 685 | 0.060886 | 9.988246 | AB\|AA |  |
| ID102 | NF435 | 122 | 2637 | 0.072106 | 172.2562 | AA\|AB |  |
| ID103 | NF435 | 567 | 658 | 0.072106 | 6.760378 | AB\|AA |  |
| ID104 | NF436 | 966 | 942 | 0.130266 | 26.36663 | AB\|AB |  |
| ID105 | NF440 | 661 | 669 | 0.089206 | 9.195065 | AB\|AB |  |
| ID106 | NF442 | 32 | 992 | 0.059919 | 29.81635 | AA\|AB |  |
| ID107 | NF442 | 898 | 1013 | 0.059919 | 6.924515 | AB\|AA |  |
| ID108 | NF443 | 1098 | 1150 | 0.094056 | 10.16467 | AB\|AB |  |
| ID109 | NF448 | 1205 | 1402 | 0.094117 | 13.9965 | AB\|AA |  |
| ID110 | NF452 | 112 | 2661 | 0.063794 | 140.7897 | AA\|AB |  |
| ID111 | NF452 | 2292 | 2889 | 0.063794 | 55.14584 | AB\|AA |  |
| ID112 | NF455 | 95 | 1922 | 0.076653 | 143.6848 | AA\|AB |  |
| ID113 | NF455 | 840 | 1032 | 0.076653 | 18.46092 | AB\|AA |  |
| ID114 | NF456 | 630 | 624 | 0.052152 | 2.78894 | AB\|AB |  |
| ID115 | NF460 | 1005 | 1014 | 0.131089 | 32.62338 | AB\|AB |  |
| ID116 | NF462 | 810 | 1068 | 0.112159 | 34.34418 | AB\|AA |  |
| ID117 | NF463 | 556 | 671 | 0.105206 | 10.63072 | AB\|AA |  |
| ID118 | NF463 | 159 | 2164 | 0.105206 | 337.5656 | AA\|AB |  |
| ID119 | NF466 | 1540 | 1614 | 0.05196 | 0.829777 | AB\|AB | AB\|AA |
| ID120 | NF468 | 627 | 633 | 0.051573 | 2.736388 | AB\|AB |  |
| ID121 | NF469 | 66 | 1105 | 0.097152 | 120.7951 | AA\|AB |  |
| ID122 | NF469 | 627 | 595 | 0.097152 | 5.351208 | AB\|AB |  |
| ID123 | NF470 | 505 | 726 | 0.132158 | 37.06724 | AB\|AA |  |
| ID124 | NF473 | 819 | 1011 | 0.128856 | 19.11715 | AB\|AA |  |
| ID125 | NF475 | 587 | 757 | 0.08542 | 19.27122 | AB\|AA |  |
| ID126 | NF479 | 53 | 974 | 0.079841 | 86.9543 | AA\|AB |  |
| ID127 | NF479 | 858 | 876 | 0.079841 | 8.208397 | AB\|AB |  |
| ID128 | NF485 | 882 | 904 | 0.108152 | 16.23624 | AB\|AB |  |
| ID129 | NF485 | 201 | 2981 | 0.108152 | 406.9719 | AA\|AB |  |
| ID130 | NF492 | 663 | 679 | 0.122459 | 16.33901 | AB\|AB |  |
| ID131 | NF497 | 585 | 688 | 0.061383 | 7.855274 | AB\|AA |  |
| ID132 | NF498 | 538 | 640 | 0.12897 | 6.69846 | AB\|AA |  |
| ID133 | NF500 | 1062 | 944 | 0.091639 | 4.770658 | AB\|AA | AB\|AB |
| ID134 | NF501 | 1483 | 1552 | 0.098058 | 15.74802 | AB\|AB |  |
| ID135 | NF503 | 1166 | 1396 | 0.062445 | 18.75245 | AB\|AA |  |
| ID136 | NF503 | 102 | 2763 | 0.062445 | 112.5169 | AA\|AB |  |
| ID137 | NF503 | 1281 | 1229 | 0.062445 | 3.303927 | AB\|AB |  |
| ID138 | NF505 | 920 | 900 | 0.086836 | 10.29345 | AB\|AB |  |
| ID139 | NF506 | 447 | 615 | 0.228475 | 21.21026 | AB\|AA |  |
| ID140 | NF509 | 1649 | 1601 | 0.139237 | 50.17194 | AB\|AB |  |
| ID141 | NF509 | 219 | 3133 | 0.139237 | 460.7324 | AA\|AB |  |
| ID142 | NF513 | 825 | 746 | 0.050599 | 3.974151 | AB\|AA |  |
| ID143 | NF514 | 186 | 3083 | 0.075818 | 319.406 | AA\|AB |  |
| ID144 | NF514 | 705 | 854 | 0.075818 | 13.64964 | AB\|AA |  |
| ID145 | NF518 | 1687 | 1669 | 0.06228 | 10.79771 | AB\|AB |  |
| ID146 | NF520 | 1575 | 1490 | 0.103283 | 15.25032 | AB\|AB |  |
| ID147 | NF523 | 635 | 807 | 0.080793 | 18.41 | AB\|AA |  |
| ID148 | NF523 | 130 | 2606 | 0.080793 | 200.4662 | AA\|AB |  |
| ID149 | NF525 | 1590 | 2101 | 0.114576 | 68.83813 | AB\|AA |  |
| ID150 | NF527 | 699 | 983 | 0.116085 | 43.41458 | AB\|AA |  |
| ID151 | NF528 | 142 | 2891 | 0.068954 | 206.7564 | AA\|AB |  |
| ID152 | NF528 | 661 | 798 | 0.068954 | 11.96984 | AB\|AA |  |
| ID153 | NF529 | 615 | 804 | 0.116499 | 24.84688 | AB\|AA |  |
| ID154 | NF530 | 1381 | 1695 | 0.091196 | 31.74151 | AB\|AA |  |
| ID155 | NF531 | 1085 | 1100 | 0.09073 | 15.33197 | AB\|AB |  |
| ID156 | NF533 | 1444 | 1160 | 0.076682 | 28.28394 | AB\|AA |  |
| ID157 | NF534 | 1707 | 1883 | 0.071954 | 6.736651 | AB\|AA | AB\|AB |
| ID158 | NF535 | 2469 | 2426 | 0.054087 | 9.684868 | AB\|AB |  |
| ID159 | NF539 | 709 | 715 | 0.126063 | 21.291 | AB\|AB |  |
| ID160 | NF539 | 123 | 1503 | 0.126063 | 287.4021 | AA\|AB |  |
| ID161 | NF541 | 2255 | 2310 | 0.057788 | 8.906468 | AB\|AB |  |
| ID162 | NF544 | 76 | 1184 | 0.074627 | 133.8991 | AA\|AB |  |
| ID163 | NF544 | 1214 | 1366 | 0.074627 | 8.320208 | AB\|AA |  |
| ID164 | NF545 | 1242 | 1457 | 0.083053 | 17.11357 | AB\|AA |  |
| ID165 | NF546 | 1591 | 2116 | 0.106865 | 70.06025 | AB\|AA |  |
| ID166 | NF547 | 1559 | 1424 | 0.058695 | 5.571395 | AB\|AA | AB\|AB |
| ID167 | NF549 | 1251 | 886 | 0.099367 | 51.57301 | AB\|AA |  |
| ID168 | NF550 | 2392 | 2372 | 0.070996 | 21.22842 | AB\|AB |  |
| ID169 | NF551 | 2324 | 2308 | 0.075385 | 23.98126 | AB\|AB |  |
| ID170 | NF556 | 953 | 956 | 0.069842 | 8.915021 | AB\|AB |  |
| ID171 | NF558 | 1997 | 1873 | 0.127598 | 31.70938 | AB\|AB |  |
| ID172 | NF559 | 1172 | 1180 | 0.125481 | 35.30998 | AB\|AB |  |
| ID173 | NF561 | 1070 | 1281 | 0.096409 | 18.85734 | AB\|AA |  |
| ID174 | NF561 | 164 | 2817 | 0.096409 | 294.1758 | AA\|AB |  |
| ID175 | NF562 | 798 | 781 | 0.126916 | 21.30263 | AB\|AB |  |
| ID176 | NF563 | 969 | 928 | 0.085609 | 6.916907 | AB\|AB |  |
| ID177 | NF565 | 1227 | 1368 | 0.094152 | 3.523582 | AB\|AA |  |
| ID178 | NF566 | 597 | 687 | 0.05667 | 6.081344 | AB\|AA |  |
| ID179 | NF567 | 2286 | 2495 | 0.051433 | 8.853804 | AB\|AA |  |
| ID180 | NF567 | 66 | 2183 | 0.051433 | 54.81282 | AA\|AB |  |
| ID181 | NF568 | 3131 | 3153 | 0.05686 | 17.84463 | AB\|AB |  |
| ID182 | NF570 | 1465 | 1678 | 0.079075 | 14.04196 | AB\|AA |  |
| ID183 | NF571 | 127 | 3293 | 0.055064 | 139.5646 | AA\|AB |  |
| ID184 | NF571 | 1712 | 1961 | 0.055064 | 16.296 | AB\|AA |  |
| ID185 | NF572 | 1105 | 1226 | 0.083081 | 4.006632 | AB\|AA |  |
| ID186 | NF575 | 1629 | 2268 | 0.118335 | 96.98797 | AB\|AA |  |
| ID187 | NF575 | 241 | 3670 | 0.118335 | 482.9952 | AA\|AB |  |
| ID188 | NF576 | 2623 | 2790 | 0.12063 | 38.85875 | AB\|AB |  |
| ID189 | NF577 | 1728 | 1501 | 0.058568 | 15.52515 | AB\|AA |  |

| Table S13. Genotype estimation results for the wilson validation dataset using plasma mixtures | | | | | | | | | | |
| --- | --- | --- | --- | --- | --- | --- | --- | --- | --- | --- |
| SampleID | Repetition ID | Mutation | Mutant Reads | Wildtype Reads | Fetal Fraction | ∆AIC | Estimated Genotype | True Genotype | Concordance | Group Concordance |
| ID001 | 1 | S106N (pat) | 15 | 587 | 0.050 | 9.56 | AA\|AB | AA\|AB | Yes | Yes |
|  | 2 | S106N (pat) | 11 | 579 | 0.050 | 2.48 | AA\|AB | AA\|AB | Yes |  |
|  | 3 | S106N (pat) | 19 | 601 | 0.050 | 16.47 | AA\|AB | AA\|AB | Yes |  |
|  | 4 | S106N (pat) | 10 | 390 | 0.050 | 6.42 | AA\|AB | AA\|AB | Yes |  |
|  | 5 | S106N (pat) | 11 | 346 | 0.050 | 9.59 | AA\|AB | AA\|AB | Yes |  |
|  | 6 | S106N (pat) | 11 | 668 | 0.050 | 0.24 | AA\|AA | AA\|AB | No |  |
|  | 7 | S106N (pat) | 9 | 571 | 0.050 | 0.94 | AA\|AA | AA\|AB | No |  |
|  | 8 | S106N (pat) | 17 | 587 | 0.050 | 13.23 | AA\|AB | AA\|AB | Yes |  |
| ID002 | 1 | S106N (pat) | 38 | 548 | 0.100 | 77.11 | AA\|AB | AA\|AB | Yes | Yes |
|  | 2 | S106N (pat) | 34 | 585 | 0.100 | 61.19 | AA\|AB | AA\|AB | Yes |  |
|  | 3 | S106N (pat) | 27 | 569 | 0.100 | 39.98 | AA\|AB | AA\|AB | Yes |  |
|  | 4 | S106N (pat) | 35 | 576 | 0.100 | 65.15 | AA\|AB | AA\|AB | Yes |  |
|  | 5 | S106N (pat) | 39 | 627 | 0.100 | 73.82 | AA\|AB | AA\|AB | Yes |  |
|  | 6 | S106N (pat) | 33 | 570 | 0.100 | 59.21 | AA\|AB | AA\|AB | Yes |  |
| ID003 | 1 | S106N (pat) | 42 | 543 | 0.150 | 95.50 | AA\|AB | AA\|AB | Yes | Yes |
|  | 2 | S106N (pat) | 62 | 509 | 0.150 | 180.71 | AA\|AB | AA\|AB | Yes |  |
|  | 3 | S106N (pat) | 42 | 511 | 0.150 | 99.85 | AA\|AB | AA\|AB | Yes |  |
|  | 4 | S106N (pat) | 64 | 595 | 0.150 | 177.09 | AA\|AB | AA\|AB | Yes |  |
|  | 5 | S106N (pat) | 49 | 574 | 0.150 | 119.50 | AA\|AB | AA\|AB | Yes |  |
|  | 6 | S106N (pat) | 51 | 589 | 0.150 | 125.52 | AA\|AB | AA\|AB | Yes |  |
| ID004 | 1 | rs1061472 (mat) | 79 | 76 | * | 0.96 | AB\|AB | AB\|AB | Yes | Yes |
| ID005 | 1 | rs1061472 (mat) | 96 | 97 | * | 1.74 | AB\|AB | AB\|AB | Yes |  |
| ID006 | 1 | rs2274084 (mat) | 105 | 109 | 0.095 | 1.19 | AB\|AB | AB\|AB | Yes |  |
| ID007 | 1 | rs2274084 (mat) | 142 | 148 | 0.154 | 5.07 | AB\|AB | AB\|AB | Yes |  |
| ID008 | 1 | rs2274084 (mat) | 158 | 162 | 0.210 | 12.71 | AB\|AB | AB\|AB | Yes |  |
| ID009 | 1 | rs2274084 (mat) | 176 | 171 | * | 2.48 | AB\|AB | AB\|AB | Yes |  |
| ID010 | 1 | rs2274084 (mat) | 220 | 206 | 0.180 | 8.99 | AB\|AB | AB\|AB | Yes |  |
| ID011 | 1 | rs72474224 (mat) | 78 | 81 | 0.170 | 3.61 | AB\|AB | AB\|AB | Yes |  |
| ID012 | 1 | rs72474224 (mat) | 117 | 116 | * | 2.14 | AB\|AB | AB\|AB | Yes |  |
| ID013 | 1 | rs72474224 (mat) | 177 | 166 | * | 1.24 | AB\|AB | AB\|AB | Yes |  |
| ID014 | 1 | rs111033313 (mat) | 59 | 55 | * | 0.34 | AB\|AB | AB\|AB | Yes |  |
| ID015 | 1 | rs72474224 (mat/pat) | 52 | 41 | 0.146 | 1.23 | AB\|AA | AB\|AA | Yes | Yes |
| ID016 | 1 | rs80338943 (mat/pat) | 131 | 92 | 0.126 | 6.32 | AB\|AA | AB\|AA | Yes | Yes |
| * fetal fraction for the specifc sample was missing, 0.10 was assumed. | | | | | | | | | | |

**Supplementary Methods**

**Dataset:**

The insertion/deletion polymorphism dataset (BioProject ID: PRJNA387652): A panel of 44 biallelic insertion/deletion polymorphic sites plus ZFX/ZFY was amplified using cfDNA or maternal genomic DNA as template. Some of the cfDNA samples were also sequenced using low coverage whole genome sequencing.

The replication dataset (BioProject ID: PRJNA517742): Genomic DNAs from two independent blood samples were mixed generating samples with minor allele frequencies of 0.5%, 1.0%, 5% and 10%, respectively. Mixed samples were PCR amplified using 564 primer pairs and sequenced.

The monogenic mutation samples: (1) the wilson validation dataset: mixtures of maternal wildtype and paternal heterozygous mutant plasma samples were prepared, mimicking homozygous-heterozygous maternal-fetal genotypes with fetal fractions of 5% (8 replicates), 10% (6 replicates) and 15% (6 replicates), respectively. To mimic heterozygous-heterozygous maternal-fetal genotypes, plasma samples from 11 pregnant women were prepared, whereas both the mothers and the fetuses were heterozygous mutant. For heterozygous-homozygous maternal-fetal genotypes, two plasma samples were selected whereas the mothers were heterozygous mutant and the fetuses were homozygous mutant. (2) the wilson disease dataset: 8 maternal plasma samples were selected. (3) the hbb dataset: 102 maternal plasma samples and 188 target sites were analyzed. (4) the arnshl dataset: 80 maternal plasma samples and 137 target sites were analyzed. (5) the cfbest dataset: 143 maternal plasma samples and 189 target sites were analyzed. All samples were processed using cSMART or cfBEST assay, and total reads, mutant reads and fetal fraction of each target site were reported. Wildtype reads were calculated as the differences between the total reads and the mutant reads.

The simulation dataset: Five hundred random 70-bp amplicon sequences were generated, and specific mutations were introduced into each amplicon to simulate polymorphic sites having four to six alleles each and could be identified by at least two unique 12-mer indexes. Fetal fraction in each sample was simulated as one of the following values: 0.02, 0.05, 0.10, 0.15, 0.20, 0.25, 0.30, 0.35, 0.40 and 0.45. In each simulated sample, a total of 400 polymorphic sites were selected and each had 200 genomic copies. Each polymorphic site was assigned to be one of the possible maternal-fetal genotypes randomly, and different number of allelic sequence amplicons was generated. For example, if the fetal fraction was 0.05, and the genotype was “AB|AC”, 100 copies of allele seqA, 95 copies of allele seqB and 5 copies of allele seqC were produced as amplicon templates for a polymorphic site having the genotype AB|AC. After generating amplicon templates for all of the polymorphic sites in a sample, sequencing reads were simulated using the ART simulator with the following command “art_illumina -ss HSXt -amp -i <inputfile> -na -l 65 -f <fold> -o <outputfile>”.

**Reads Processing and Mapping**

Reads retrieved from SRA or simulated were filtered out using custom scripts as follows. For each read, base positions with a quality score of 14 or less were identified, and then the longest subsequence was selected whereas each base in the subsequence had a quality score greater than 14. Subsequently, filtered reads were mapped first to unique polymorphic sites using 12-mer indexes, and then each read was mapped to a specific allele using unique allelic indexes. Finally, different alleles were counted for each polymorphic site in each sample.

**Fetal Fraction Estimation**

For each polymorphic site, read counts for all alleles were sorted in descending order and labeled as R1, R2, R3, etc., and the allelic read count Ri was considered as a noise background if its relative value ($RRi=\mathrm{Ri}/{\sum_{j=1}^{i} R_{j}}$) was less than the background threshold (α). All polymorphic sites used for fetal fraction estimation were assumed to be one of the normal disomy-disomy maternal-fetal genotypes (AA|AA, AA|AB, AB|AA, AB|AB or AB|AC, where the portion before the vertical bar denotes the maternal genotype and the portion after it denotes the fetal genotype), as the majority of the polymorphic sites were from chromosomes that were normal and only a small portion of them were abnormal if any at all. For each polymorphic site from a reference normal disomy-disomy chromosome, there are at most three informative alleles exist. In an ideal situation, if one or three alleles are detected for a target site, then its genotype can be determined unambiguously, while if two alleles are detected, a single measure of R1/(R1+R2) was informative enough to classify all three possible genotypes as follows. If the genotype is AA|AB, then R1/(R1+R2)=1-0.5f, where f denotes fetal fraction. As f is the minor component and f<0.5, then 1-0.5f ≥ 0.75. If the genotype is AB|AA, then R1/(R1+R2)=0.5+0.5f. As f<0.5, then must 0.5+0.5f ≤ 0.75. If the genotype is AB|AB, then R1/(R1+R2)=0.5 irrespective of f values. Therefore, for each polymorphic site (Fig. S2, RR2=R2/(R1+R2) and RR3=R3/(R1+R2+R3)), if only one informative allele was detected (RR2<α), then the genotype would be AA|AA. If two informative alleles were present (RR2≥α and RR3<α), then the site should be one of the genotypes having two different alleles (AA|AB, AB|AA or AB|AB), which could be identified using the ratio ${R1}/{(R1+R2)}$ as follows: when the ratio ≥0.75, the genotype was estimated to be AA|AB, between 0.5+α and 0.75 to be AB|AA and between 0.5 and 0.5+α to be AB|AB. If three alleles were informative (RR3≥α), then the genotype was AB|AC if R2/R1≥0.5, and R3 was considered as a background noise outlier otherwise. Clearly, relative allelic read counts of three genotypes were affected by the sample’s fetal fraction, and the estimated read count derived from fetal genetic materials (FC, FetalReads) was calculated along with the total read count (TC, TotalReads) for each polymorphic site (Fig. S2, Table S1). Finally, the fetal read counts (FetalReads) were regressed against the total read counts (TotalReads) for a panel of polymorphic sites using the R’s rlm function in MASS package with the fitting model$y=\beta x+0$, and fetal fraction was estimated as the model coefficient$\left( \beta\right)$.

For example, the following are imaginary representative allelic read counts for five polymorphic sites from a sample (R1-R3: allelic read counts in descending order). Background α is set to 0.01.

| MarkerID | R1 | R2 | R3 |
| --- | --- | --- | --- |
| ID-01 | 14127 | 35 | 0 |
| ID-02 | 4105 | 577 | 13 |
| ID-03 | 3148 | 3101 | 54 |
| ID-04 | 5809 | 3552 | 27 |
| ID-05 | 4007 | 3028 | 1011 |

For ID-01, RR2=R2/(R1+R2)=35/(14127+35)=0.002<0.01, genotype is AA|AA. FetalReads=NA. TotalReads=R1 =14127.

For ID-02, RR2=R2/(R1+R2)=577/(4105+577)=0.123≥0.01 and RR3=R3/(R1+R2+R3)=13/(4105+577 +13) =0.003<0.01, two alleles are informative. Ratio=R1/(R1+R2)=0.877. As Ratio≥0.75, genotype is AA|AB. FetalReads=$2\times R2$=2$\times$577=1154. TotalReads=R1+R2=4682.

For ID-03, RR2=0.496≥0.01 and RR3=0.009<0.01, two alleles are informative. Ratio=0.504. As 0.5≤Ratio<0.51, genotype is AB|AB. FetalReads=NA. TotalReads=R1+R2=6249.

For ID-04, RR2=0.379≥0.01, and RR3=0.003<0.01, two alleles are informative. Ratio=0.621. As 0.51≤Ratio<0.75, genotype is AB|AA. FetalReads=R1-R2 =2257, TotalReads=R1+R2=9361.

For ID-05, RR2=0.430≥0.01, and RR3=0.126>0.01, three alleles are informative. As R2/R1=0.756≥0.5, genotype is AB|AC. FetalReads=R1-R2+R3=1990, TotalReads=R1+R2+R3=8046.

Three polymorphic sites are considered informative for fetal fraction estimation in the above sample (ID-02, ID-04 and ID-05). Hence a robust linear regression model is fitted using the three informative sites and the fetal fraction (f) is estimated by the following R commands:

FetalReads=c(NA,1154,NA,2257,1990)

TotalReads=c(14127,4682,6249,9361,8046)

rlmfit=rlm(FetalReads~TotalReads+0,maxit=1000)

f=rlmfit$coefficients["TotalReads"]

Therefore, the estimated fetal fraction (f) for the sample is 0.244.

**Maternal-Fetal Genotype Estimation for Polymorphic Sites**

For each sample, fetal fraction was estimated using a panel of allelic read counts. Then the genotype for each polymorphic site was estimated using the minimal AIC value as detailed below. First, observed allelic read counts ($O_{i}$), total read count (TotalReads) and expected allelic read counts ($E_{i}$) for each possible genotype model were calculated for each polymorphic site ($O_{i}$ is set to 0.1 if $O_{i}$ = 0 and $E_{i}$ is set to TotalReads$\times$α if the expected $E_{i}$= 0), and then AIC was calculated for each genotype model using the following formula:

$$AIC=2\times\sum\left[ O_{i}\times ln\left( \frac{O_{i}}{E_{i}} \right) \right]-2\times df$$

Where df is the residual degrees of freedom. Finally, the genotype for the polymorphic site was estimated to be the one with the minimal AIC, and AIC difference (ΔAIC) was the absolute difference between the minimal AIC and the second minimal AIC. The adjusted AIC = AIC/f/TotalReads, and the adjusted ΔAIC = ΔAIC/f/TotalReads. AIC could also be calculated using a modified formula as described below and identical genotype estimations were observed for our simulated samples. If only one allele was observed informative, then for model AA|AA, only O_1_ and E_1_ was used for AIC calculation; for models AB|AA, AB|AB and AA|AB, O_1_-O_2_ and E_1_-E_2_ were used; and for model AB|AC, both O_1_-O_3_ and E_1_-E_3_ were used. Similarly, if two or three alleles were observed informative, then two or three O_i_ and E_i_ were used for AIC calculation depending on the specific fitted genotype model.

For example, the estimated fetal fraction for the imaginary sample is f=0.244 and if the expected model fitting background (α) is set to 0.005, then the observed and the expected allelic read counts were calculated as follows:

For ID-01, observed allelic read counts are [14127, 35, 0], then $O_{1}$=14127, $O_{2}$=35 and $O_{3}$=0.1, TotalReads=R1+R2+R3=14127+35+0=14162, df=3-1=2, f=0.244.

Fitting AA|AA model:

$E_{1}$=$TotalReads\times\left( 1-2\times\alpha\right)$=14162$\times(1-2\times0.005)$=14020.38

$E_{2}$=$TotalReads\times\alpha$=70.81

$E_{3}$=$TotalReads\times\alpha$=70.81

$\mathrm{AIC}_{AA|AA}$=$2\times\left( O_{1}\times ln\frac{O_{1}}{E_{1}}+O_{2}\times ln\frac{O_{2}}{E_{2}}{+O}_{3}\times ln\frac{O_{3}}{E_{3}} \right)-2\times df$=159.41

${Adjusted AIC}_{AA|AA}$=${{AIC}_{AA|AA}}/{f/{TotalReads}}$=0.046

Fitting AA|AB model:

$E_{1}$=$TotalReads\times\left( 1-\alpha\right)\times{(2-f)}/2$=12372.06

$E_{2}$=$TotalReads\times\left( 1-\alpha\right)\times f/2$=1719.13

$E_{3}$=$TotalReads\times\alpha$=70.81

$\mathrm{AIC}_{AA|AB}$=3469.89

${Adjusted AIC}_{AA|AB}$=1.004

Fitting AB|AA model:

$E_{1}$=$TotalReads\times\left( 1-\alpha\right)\times({1+f)}/2$=8764.78

$E_{2}$=$TotalReads\times\left( 1-\alpha\right)\times({1-f)}/2$=5326.51

$E_{3}$=$TotalReads\times\alpha$=70.81

$\mathrm{AIC}_{AB|AA}$=13129.87

${Adjusted AIC}_{AB|AA}$=3.800

Fitting AB|AB model:

$E_{1}$=$TotalReads\times\left( 1-\alpha\right)\times1/2$=7045.60

$E_{2}$=$TotalReads\times\left( 1-\alpha\right)\times1/2$=7045.60

$E_{3}$=$TotalReads\times\alpha$=70.81

$\mathrm{AIC}_{AB|AB}$=19279.24

${Adjusted AIC}_{AB|AB}$=5.579

Fitting AB|AC model:

$E_{1}$=${TotalReads\times1}/2$=7081.00

$E_{2}$=$TotalReads\times({1-f)}/2$=5353.24

$E_{3}$=$TotalReads\times f/2$=1727.76

$\mathrm{AIC}_{AB|AB}$=19156.21

${Adjusted AIC}_{AB|AB}$=5.544

As $\mathrm{AIC}_{AA|AA}<\mathrm{AIC}_{AA|AB}<\mathrm{AIC}_{AB|AA}<\mathrm{AIC}_{AB|AC}<\mathrm{AIC}_{AB|AB}$, the estimated genotype for ID-01 site is AA|AA.

Minimal AIC=$\mathrm{AIC}_{AA|AA}$=159.41

Minimal adjusted AIC=${Adjusted AIC}_{AA|AA}$=0.046

ΔAIC=${{AIC}_{AA|AB}-AIC}_{AA|AA}$=3469.89-159.41=3310.48

Adjusted ΔAIC=${\Delta AIC}/{f/{TotalReads}}$=${Adjusted AIC}_{AA|AB}-{Adjusted AIC}_{AA|AA}$=1.004-0.046=0.958

The AICs for all other sites are calculated similarly and listed below.

| Marker  ID | Total Reads | AIC | | | | | Estimated Genotype | Minimal AIC | Minimal Adjusted AIC |
| --- | --- | --- | --- | --- | --- | --- | --- | --- | --- |
|  |  | AA\|AA | AA\|AB | AB\|AA | AB\|AB | AB\|AC |  |  |  |
| ID-01 | 14162 | 159.41 | 3469.89 | 13129.87 | 19279.24 | 19156.21 | AA\|AA | 159.41 | 0.046 |
| ID-02 | 4695 | 2655.61 | 1.67 | 1526.69 | 2996.40 | 3189.20 | AA\|AB | 1.67 | 0.001 |
| ID-03 | 6303 | 24207.47 | 5213.19 | 369.79 | 9.62 | 1336.75 | AB\|AB | 9.62 | 0.006 |
| ID-04 | 9388 | 25240.70 | 4035.06 | 6.14 | 555.64 | 2276.37 | AB\|AA | 6.14 | 0.003 |
| ID-05 | 8046 | 27177.07 | 8863.34 | 4777.25 | 4833.02 | -3.01 | AB\|AC | -3.01 | -0.001 |

**Maternal-Fetal Genotype Estimation for Short Genetic Variations**

For each site, the maternal-fetal genotype group (AA|AA, AA|AB, AB|AA, AB|AB or AB|AC) was estimated first using its allelic read counts. Then, the wildtype sequence was compared with its different alleles (Table S7, S8) and the maternal-fetal mutational status was determined accordingly. For example, if the target site had the genotype AA|AA, and the R1 allele’s sequence was wildtype, then the maternal-fetal genotype was WW|WW, while if the R1’s sequence was mutant, then it was MM|MM, where W for wildtype and M for mutant. The wildtype/mutant status for other genotypes could be processed similarly.

**Maternal-Fetal Chromosomal/Subchromosomal Abnormality Detection**

If a polymorphic site is from a diploid mother carrying a diploid fetus (herein labeled as disomy-disomy for each chromosome), it can only be one of the following five maternal-fetal genotypes, namely AA|AA, AA|AB, AB|AA, AB|AB and AB|AC. However, if the polymorphic site is on the target chromosome of a diploid mother carrying a trisomy fetus (labeled as disomy-trisomy for the target chromosome), it can only be one of the following ten genotypes (AA|AAA, AA|AAB, AA|ABB, AA|ABC, AB|AAA, AB|AAB, AB|AAC, AB|ABC, AB|ACC and AB|ACD). In each cfDNA sample containing a possible trisomy fetal chromosome, all polymorphic sites on the target chromosome are either all disomy-disomy or all disomy-trisomy, but not both. Therefore, for each target polymorphic site, the minimal adjusted AIC for each chromosomal model was calculated first, and then the model that shows best overall fit for all polymorphic sites is selected. To detect fetal chromosomal monosomy, each polymorphic site is tested against all possible genotypes for a chromosome that is possible monosomy in fetus, and the model that shows best overall fit for all polymorphic sites is selected. Subchromosomal deletions/duplications could be detected similarly using all possible chromosomal models for the target chromosome.

For example, the following are imaginary representative allelic read counts on a target chromosome for two samples, and each has five polymorphic sites on the target chromosome (R1-R4: allelic read counts in descending order). Suppose that the target chromosome is chromosome 21, and we want to test if any of the two samples are trisomy 21. Background α is set to 0.01.

| SampleId | SiteId | Allelic Counts (Descending) | | | |
| --- | --- | --- | --- | --- | --- |
|  |  | R1 | R2 | R3 | R4 |
| S001 | Id001 | 9565 | 14 | 4 | 0 |
|  | Id002 | 5820 | 652 | 6 | 3 |
|  | Id003 | 6718 | 4465 | 12 | 5 |
|  | Id004 | 7838 | 7656 | 34 | 12 |
|  | Id005 | 9465 | 7552 | 1898 | 33 |
| S002 | Id001 | 7021 | 1574 | 7 | 3 |
|  | Id002 | 10588 | 1185 | 1164 | 23 |
|  | Id003 | 3408 | 2861 | 23 | 12 |
|  | Id004 | 9059 | 6012 | 1505 | 34 |
|  | Id005 | 9386 | 9373 | 1899 | 18 |

Then each polymorphic site is tested against all genotypes of both the disomy-disomy model and the disomy-trisomy model, and the best fit genotypes for the disomy-disomy model and the disomy-trisomy model are listed below.

| Overall goodness-of-fit test results for different chromosomal models | | | | | | | | | |
| --- | --- | --- | --- | --- | --- | --- | --- | --- | --- |
| SampleId | SiteId | Best Fit Genotype for Each Model | | | | | | | |
|  |  | Disomy-Disomy Model | | | | Disomy-Trisomy Model | | | |
|  |  | Genotype | TC | G | AIC | Genotype | TC | G | AIC |
| S001 | Id001 | AA\|AA | 9565 | 0 | 0 | AA\|AAA | 9565 | 0 | 0 |
|  | Id002 | AA\|AB | 6472 | 0.039 | -1.961 | AA\|AAB | 6472 | 7.338 | 5.338 |
|  | Id003 | AB\|AA | 11183 | 0.025 | -1.975 | AB\|AAA | 11183 | 60.564 | 58.564 |
|  | Id004 | AB\|AB | 15494 | 2.138 | 0.138 | AB\|AAB | 15494 | 97.537 | 95.537 |
|  | Id005 | AB\|AC | 18915 | 0.054 | -3.946 | AB\|AAC | 18915 | 154.291 | 150.291 |
| S002 | Id001 | AA\|AB | 8595 | 543.745 | 541.745 | AA\|ABB | 8595 | 0.099 | -1.901 |
|  | Id002 | AA\|AB | 12937 | 3131.2 | 3129.2 | AA\|ABC | 12937 | 0.193 | -3.807 |
|  | Id003 | AB\|AB | 6269 | 47.789 | 45.789 | AB\|AAB | 6269 | 0.084 | -1.916 |
|  | Id004 | AB\|AC | 16576 | 143.656 | 139.656 | AB\|AAC | 16576 | 0.077 | -3.923 |
|  | Id005 | AB\|AC | 20658 | 245.48 | 241.48 | AB\|ABC | 20658 | 0.266 | -3.734 |

For the sample S001, nearly all polymorphic sites fit the disomy-disomy model better than the disomy-trisomy model, hence chromosome 21 in the sample S001 is normal for both the mother and the fetus.

For the sample S002, all polymorphic sites fit the disomy-trisomy model better than the disomy-disomy model, hence chromosome 21 in the sample S002 is normal for the mother and trisomy for the fetus.

**Plotting Distributions of Relative Allelic Read Counts**

For the disomy-disomy model, five maternal-fetal genotypes are possible and there are at most three alleles for each polymorphic site. Hence, knowing the relative allelic counts for any two alleles is informative enough to calculate the relative counts for the third one. Therefore, for each polymorphic site, allelic read counts were calculated and labeled as R1, R2 and R3, whereas R1≥R2≥R3, and then the relative allelic counts RRC1=R1/(R1+R2+R3) and RRC2= R2/(R1+R2+R3) were calculated, followed by the plotting of RRC2 against RRC1. For the disomy-monosomy model or the subchromosomal deletion model, RRC1 was plotted against RRC2 similarly for each polymorphic site, and distinct clusters corresponding to different chromosomal genotypes were shown in the generated plot. As there were at most four alleles for each polymorphic site for the disomy-trisomy model or the subchromosomal duplication model, RRC1 was calculated as R1/(R1+R2+R3+R4), RRC2 as R2/(R1+R2+R3+R4) , RRC3 as R3/(R1+R2+R3+R4) and RRC4 as R4/(R1+R2+R3+R4) . Then, RRC2 and RRC4 were plotted against RRC1 for the disomy-trisomy model, while RRC2 and RRC3 were plotted against RRC1 for the subchromosomal duplication model to distinguish all possible genotypes graphically.

**Plotting Short Genetic Variations**

For each polymorphic site, the wildtype allele (R_w_) was counted first followed by the count of mutant alleles as R_m1_, R_m2_, R_m3_, whereas R_m1_≥R_m2_≥R_m3_. Then the relative mutant allele 1’s count (R_m1_/TotalCount) was plotted against the relative wildtype allele count (R_w_/TotalCount) and all the possible maternal-fetal wildtype-mutant genotypes could be identified on the generated graph easily.

**Fetal Fraction Estimation for cfDNA samples from surrogate mothers**

For cfDNA samples from surrogate mothers, at most 4 alleles are possible for each polymorphic site on a normal chromosome. To estimate fetal fraction using a panel of polymorphic sites (Fig. S15), an initial fetal fraction estimate (f_0_) is set, followed by iteratively updating f_0_ until converge. To update f_0_, allelic goodness-of-fit test is performed for each polymorphic site to select the best fit genotype under the current f_0_ estimate, followed by the estimation of read count derived from fetal genetic materials (FC, FetalReads) and the total read count (TC, TotalReads) , and then new fetal fraction (f) is estimated by fitting a rlm model using the FCs and TCs of all polymorphic sites. Finally f_0_ is set to f, f_0_ is updated iteratively until the change of f_0_ for each iteration is very small (|f-f_0_|<ε).

For example, the following are imaginary representative allelic read counts for nine polymorphic sites from a sample (R1-R5: allelic read counts in descending order). Background α is set to 0.01, ε=0.001.

| SiteId | Allelic Counts | | | | |
| --- | --- | --- | --- | --- | --- |
|  | 1 | 2 | 3 | 4 | 5 |
| Id001 | 35 | 14127 |  |  |  |
| Id002 | 4105 | 577 | 13 | 7 | 9 |
| Id003 | 54 | 3101 | 3148 | 23 |  |
| Id004 | 11 | 5809 | 27 | 3552 | 17 |
| Id005 | 3028 | 1011 | 4007 | 6 | 6 |
| Id006 | 36 | 936 | 3322 | 28 | 16 |
| Id007 | 5422 | 52 | 974 | 938 | 27 |
| Id008 | 1498 | 4835 | 1537 | 4711 | 38 |
| Id009 | 36 | 3412 | 2237 | 3493 | 23 |

Step 1, set f_0_=0.10 (initial estimate).

Step 2, for each polymorphic site, estimate FC and TC using allelic goodness-of-fit test. For example, for site Id006, R1 to R4 are set to 3322, 936, 36 and 28. As R2/(R1+R2)≥α and R3/(R1+R2+R3)<α, there are two informative alleles. The allelic counts R1 to R4 are tested against 9 genotype models (AA|AA, AA|AB, AB|AA, AB|AB, AB|AC, AA|BB, AA|BC, AB|CC, AB|CD) assuming fetal fraction is f_0_. As the best fit genotype for Id006 is AA|BB, FC=936 and TC=4258. FCs and TCs for all other polymorphic sites are estimated similarly.

Step 3, calculate fetal fraction f by fitting a robust linear regression model for all FC and TC pairs.

Step 4, if |f-f_0_|>ε, then set f_0_=f and execute Step 2; else the fetal fraction is estimated as f.

The iterative results for the above example are listed below.

| Iterative results for fetal fraction estimation | | | |
| --- | --- | --- | --- |
| Iterative Step | f_0_ | f | **\|**f-f_0_**\|** |
| 1 | 0.1 | 0.2385 | 0.1385 |
| 2 | 0.2385 | 0.2436 | 0.0051 |
| 3 | 0.2436 | 0.2436 | 0 |

Therefore, the estimated fetal fraction (f) for the sample is 0.2436.
